# Experiences of Indigenous peoples living with pelvic health conditions: A scoping review

**DOI:** 10.1101/2024.07.22.24309744

**Authors:** Kaeleigh Brown, Katherine Choi, Esther Kim, Sandra M Campbell, Jane Schulz, Pertice Moffitt, Susan Chatwood

**Author notes:** Corresponding author (KB) Kaeleigh Brown.

## Abstract

**Background:** Pelvic health conditions significantly impact quality of life and are prevalent in the general population. Urinary and fecal incontinence, pelvic organ prolapse, and pelvic pain are examples of pelvic health conditions. A scoping review was conducted to understand what is currently known about pelvic health conditions experienced by Indigenous populations worldwide. To date, no such review has been reported.

**Methods:** A scoping review methodology was used. In June 2023, a search was conducted, and then updated in February 2024, capturing both primary and grey literature. An iterative process of abstract and full text screening was conducted by two reviewers before proceeding to data extraction. Inclusion criteria focused on English publications and reports of pelvic health conditions experienced by Indigenous peoples. Data was collected in Google Sheets, and then underwent descriptive statistical analysis. Publications that provided qualitative data were analyzed using thematic analysis.

**Results:** A total of 203 publications were included in the analysis. Several patterns emerged regarding publication region, gender and age representation, pelvic health conditions reported, and representation of Indigenous peoples. Notable gaps were a lack of publications from China, Russia, and Nordic countries, minimal representation of gender diverse populations, few publications reporting on auto-immune and bowel conditions, and limited mention of trauma-informed and culturally safe approaches.

**Conclusions:** This study highlights gaps in the current literature around gender representation, bowel and auto-immune conditions, regional representation, and the use of safety frameworks, which may inform future research initiatives. It also summarizes the existing literature, which may inform clinical and health system-level decision making.

## Introduction

To date, there has been no published review of pelvic health conditions experienced by Indigenous populations. The purpose of this scoping review is to describe what is currently known about Indigenous peoples’ experiences with pelvic health conditions by asking the question: “What does the literature say about pelvic health conditions experienced by Indigenous populations worldwide?”. Where there is a need for exploratory work in this area, we decided a scoping review method was best suited. This scoping review will identify gaps and patterns in the existing literature and provide recommendations for future research and practice [1].

The United Nations (UN) estimates that there are at least 370 million Indigenous peoples across the globe [2]. Indigenous peoples is a broad term used to describe the original inhabitants of a region prior to the arrival of other peoples [2]. To increase understanding of the term “indigenous”, the UN outlines several criteria, rather than offering an official definition. These criteria include: self-identification, relationships with the land, association with pre-colonial/settler societies, distinct knowledges (e.g., culture, language, social systems), belonging to non-dominant groups of society, and commitment to continue as “distinctive peoples and communities” [2]. Indigenous peoples experience barriers to health service access [3,4], and poorer health outcomes than non-Indigenous counterparts [5–9]. There have been calls from national [5,10,11] and international [12,13] fronts to address these attitudes and inequities.

The pelvic floor includes interconnections between many body systems which are housed within the bony pelvis. These structures are supported by connective tissue, ligaments, and the pelvic floor muscles. When there is a change in these supporting structures, whether due to aging, pregnancy and childbirth, nervous system sensitivity, or pathology, an individual may experience symptoms of a pelvic health condition. Symptoms of pelvic health conditions in men include erectile dysfunction, in women include protrusion of pelvic organs (prolapse), and in both genders include urinary or bowel incontinence/urgency/frequency or pain (with voiding, defecation, or intercourse). Several treatment options exist for pelvic health conditions, including medication, pelvic floor rehabilitation, surgery, continence products, pessaries, and behavioural modifications.

These conditions are prevalent in the general population [14–18] and the number of individuals affected is expected to increase [19]. The existing literature demonstrates varying prevalence rates across race/ethnicity [20,21], particularly with an over-representation of White people and an under-representation of Indigenous people and people of colour in pelvic health research [22–24]. This may have impacts on clinical decision making and generalizability of research results [22–24].

## Methods

We selected a scoping review methodology to systematically search and analyze the existing literature on pelvic health conditions experienced by Indigenous peoples worldwide. This methodology was a good fit for the topic, since it has yet to be systematically explored, and because the scope of the research question was broadened beyond interventional studies [25]. Although our search strategy was developed in advance, we did not register the scoping review protocol. Several authors have explored and further defined scoping review methodology (see Arksey & O’Malley [25], Levac et al [26], and Bradbury-Jones et al [1]). Our review was informed by all three publications.

### Search Strategy

A search was executed by an expert searcher/health librarian (SMC) from the University of Alberta, Edmonton, Canada, on the following databases: PROSPERO, OVID Medline, OVID EMBASE, OVID Global Health, Wiley Cochrane Library (CDSR and Central), EBSCO CINAHL, Proquest Dissertations and Theses Global and SCOPUS using controlled vocabulary (eg: MeSH, Emtree, etc) and key words representing the concepts "pelvic health conditions", "Indigenous People". Variants of search filters were applied for "Indigenous People" [27–40]. Search terms for pelvic health conditions were informed by the International Continence Society’s Incontinence, 6th Edition [41], the first author’s (KB) clinical experience, and by a practicing gynecologist (JS). Searches were adjusted for each database. No limits were applied. Databases were searched from inception to mid-February 2024. Detailed search strategies are available in **S1 Appendix**.

Reference lists of the included full texts were also searched to identify any articles that may meet the inclusion criteria. In one case [22], the corresponding author was contacted to acquire a list of citations used in their analysis. Additionally, hand searching of key journals (BMC Women’s Health, Journal of Women’s & Pelvic Health Physical Therapy, and International Urogynecology Journal) was conducted from inception to January 31, 2024. Two organization websites (World Health Organization and International Continence Society) and an internet search engine (DuckDuckGo) were also searched to identify additional resources from the media, organizations, and conference presentations.

Once the results were uploaded to Covidence [42], two rounds of review were conducted by two reviewers (either KB and KC, or KB and EK): first screening the abstracts and titles, followed by screening the included full text resources. Screening criteria for each round are outlined in **Table 1**. If there was a discrepancy between the two reviewers, they met to discuss and come to a consensus on whether to exclude or proceed with inclusion. This was an iterative process, where reviewers modified the screening criteria based on the search results, and during data collection, where the co-authors identified literature that did not meet the inclusion criteria and were subsequently excluded.

**Table 1.**
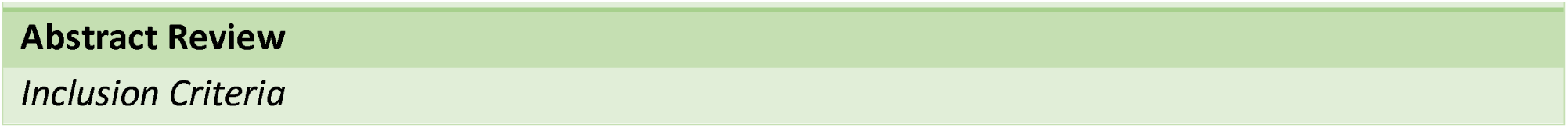
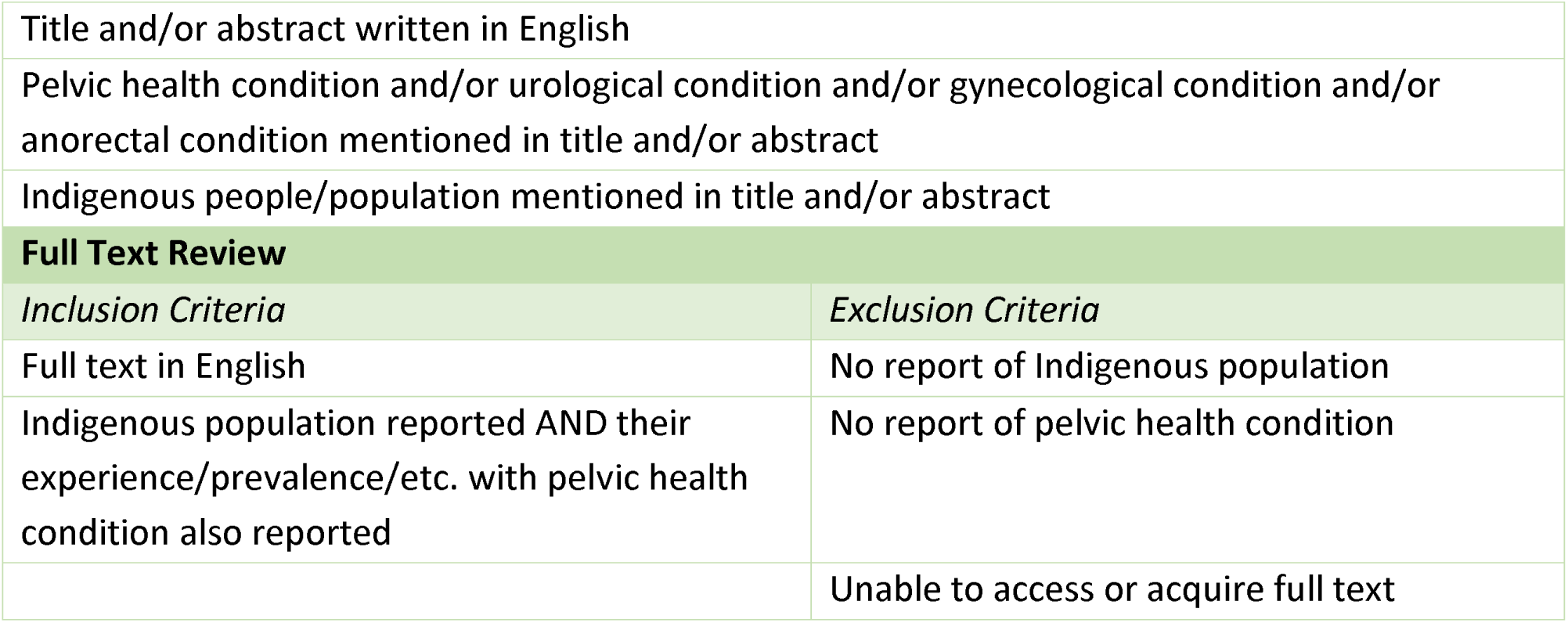
Scoping review inclusion and exclusion criteria.

|  |  |
| --- | --- |
| Title and/or abstract written in English |  |
| Pelvic health condition and/or urological condition and/or gynecological condition and/or anorectal condition mentioned in title and/or abstract |  |
| Indigenous people/population mentioned in title and/or abstract |  |
| **Full Text Review** |  |
| *Inclusion Criteria* | *Exclusion Criteria* |
| Full text in English | No report of Indigenous population |
| Indigenous population reported AND their experience/prevalence/etc. with pelvic health condition also reported | No report of pelvic health condition |
|  | Unable to access or acquire full text |

### Data Collection

Data extraction, also known as charting the data [25], is an iterative process, which evolves as the reviewers become more familiar with the literature [26]. To ensure extraction was consistent between reviewers, KB, KC and EK extracted data from up to ten articles, and then met to share their findings [26]. Any discrepancies or disagreements were discussed between reviewers. Data extraction continued, and a Google Sheets spreadsheet used to collate the results. Additional meetings between reviewers occurred ad hoc, to ensure data extraction was responsive to the literature. The extracted data included: population demographics (e.g., age, parity, medical history), population socioeconomic factors, publication characteristics, region information (e.g., country, targeted recruitment of Indigenous peoples), pelvic floor information (e.g., epidemiology, conditions reported, interventions, help-seeking behaviours, described experiences with pelvic health conditions), and safety information (cultural safety, trauma-informed approach). These headings were developed from an existing template used in previous scoping reviews by the final author (SC), and then modified to reflect the pelvic health and Indigenous health literature.

Indigenous people have experienced and continue to experience discrimination and harm in healthcare and research [6]. Culturally safe and trauma-informed practices have been proposed to improve safety in healthcare situations. We believed it was important to include headings for cultural safety and trauma-informed practices/frameworks to understand the extent of their implementation in pelvic health research with Indigenous peoples. Cultural safety is a social justice approach to healthcare based in Māori Indigenous knowledges [43,44]. It addresses power dynamics between patients and healthcare providers and has been adopted internationally [44]. The concept within healthcare has been interpreted in several ways [44], but its use within research has not been explored extensively [43]. Traumatic experiences are prevalent, such as around birth and sexual trauma [45,46], and there is an association between sexual trauma and pelvic health conditions [47]. Trauma Theory suggests that traumatic experiences can affect how the body functions, including mental and physical health [48,49]. By ‘becoming’ trauma-informed, it is assumed that adopting a trauma-informed approach will have a positive effect on service users and participants [49,50]. Trauma-informed practice is a newly evolving construct in human research, but has been used in clinical and educational practices for a number of years.

### Analysis

The PAGER framework guided analysis and reporting of the literature review findings [1]. The five domains of the framework are: Patterns, Advances, Gaps, Evidence for practice, and Research recommendations. Bradbury-Jones et al [1] recommend using a patterning chart to identify trends in methodology and themes. Due to the number of included publications, we opted out of this process, and instead looked for patterns within the charted data around publication characteristics, participant demographics, regions, pelvic health conditions, Indigenous representation, and safety. Following pattern identification, we then proceeded to examine the evolution of the research – specifically looking at theoretical and methodological advances [1]. To identify gaps in the literature, interconnections between themes and the current research and practice landscape were considered [1]. Evidence for practice and research recommendations were considered at the end of analysis to provide potential knowledge users with information, and guidance on what the next research steps may be [1,26].

## Results

Results (2911) were exported to Covidence review management software [42], where duplicates (453) were removed. Nineteen additional duplicates were removed manually. After abstract and full text screening, a total of 243 resources were included, of which 40 were merged due to duplication of publication (e.g., same content published in several conference abstracts, or conference abstract merged with original journal article). This left 203 publications for analysis. See **Fig 1** for a summary of the study selection process, as illustrated in a Preferred Report Items for Systematic Review and Meta-Analysis (PRISMA) flow chart [51].

**Fig 1.**
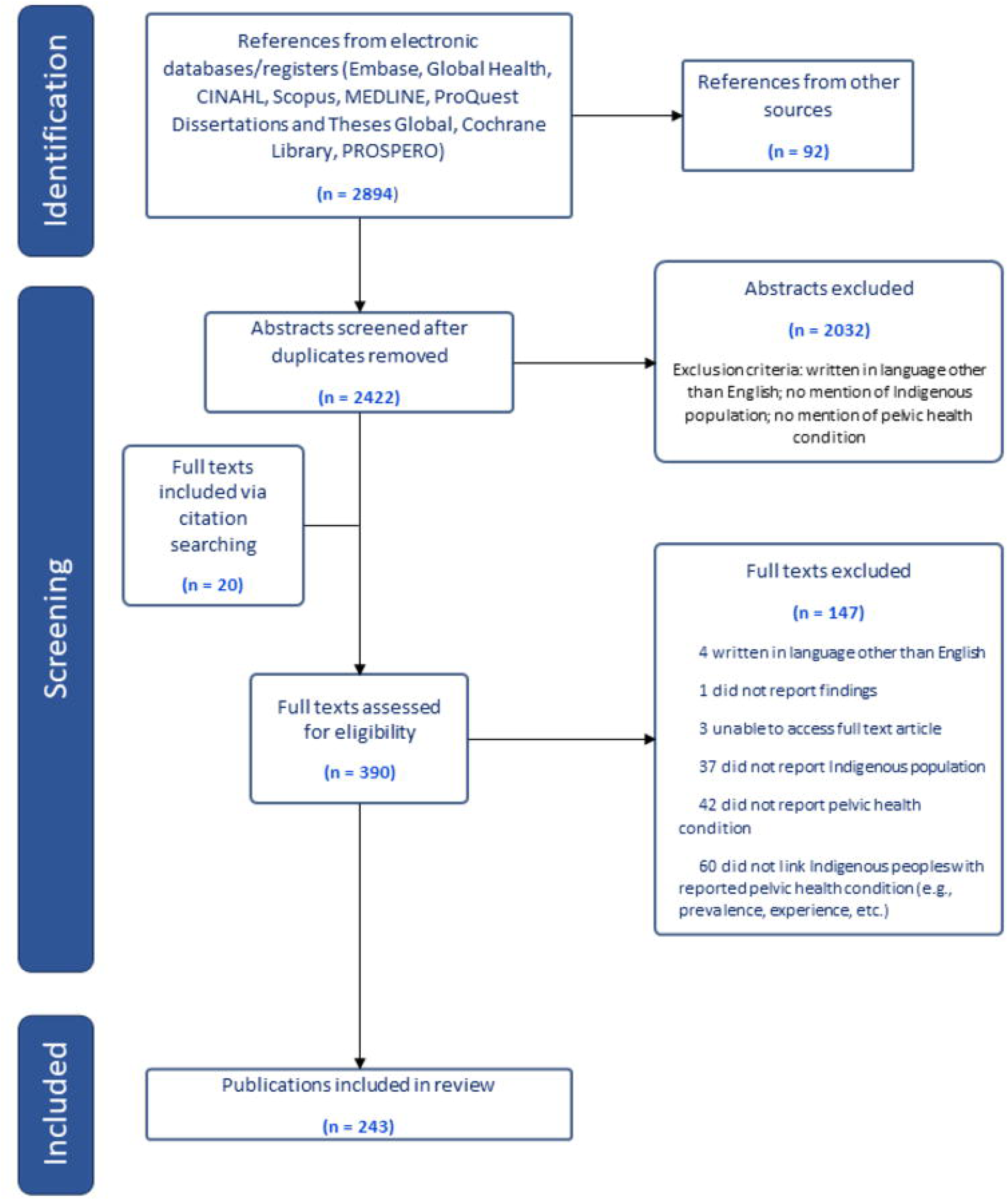
PRISMA flow chart.

### Publication Characteristics

**Fig 2** provides a summary of all included studies based on region of publication. North America was represented the most (40.9% of publications), whereas the Middle East (2%) and multinational (1.5%) publications were the least represented. Notably, there were two publications from a circumpolar region (Northwest Territories, Canada; Alaska, United States of America), which includes Nordic countries, Russia, Greenland, Alaska, and the northern territories of Canada. The majority of publications were from the primary literature (74.3%) and utilized a quantitative methodology (63.1%). The proportion of Indigenous peoples in the publications varied widely, from 0-100%. The proportion of Indigenous participants was reported 231 times (this is due to multiple treatment arms in interventional studies, or several Indigenous groups identified within a publication – where the total did not equal 100%). As shown in **Fig 3**, the distribution of representation weighed heavily towards 100% (32% of instances), and towards <5% (28.6%).

**Fig 2.**
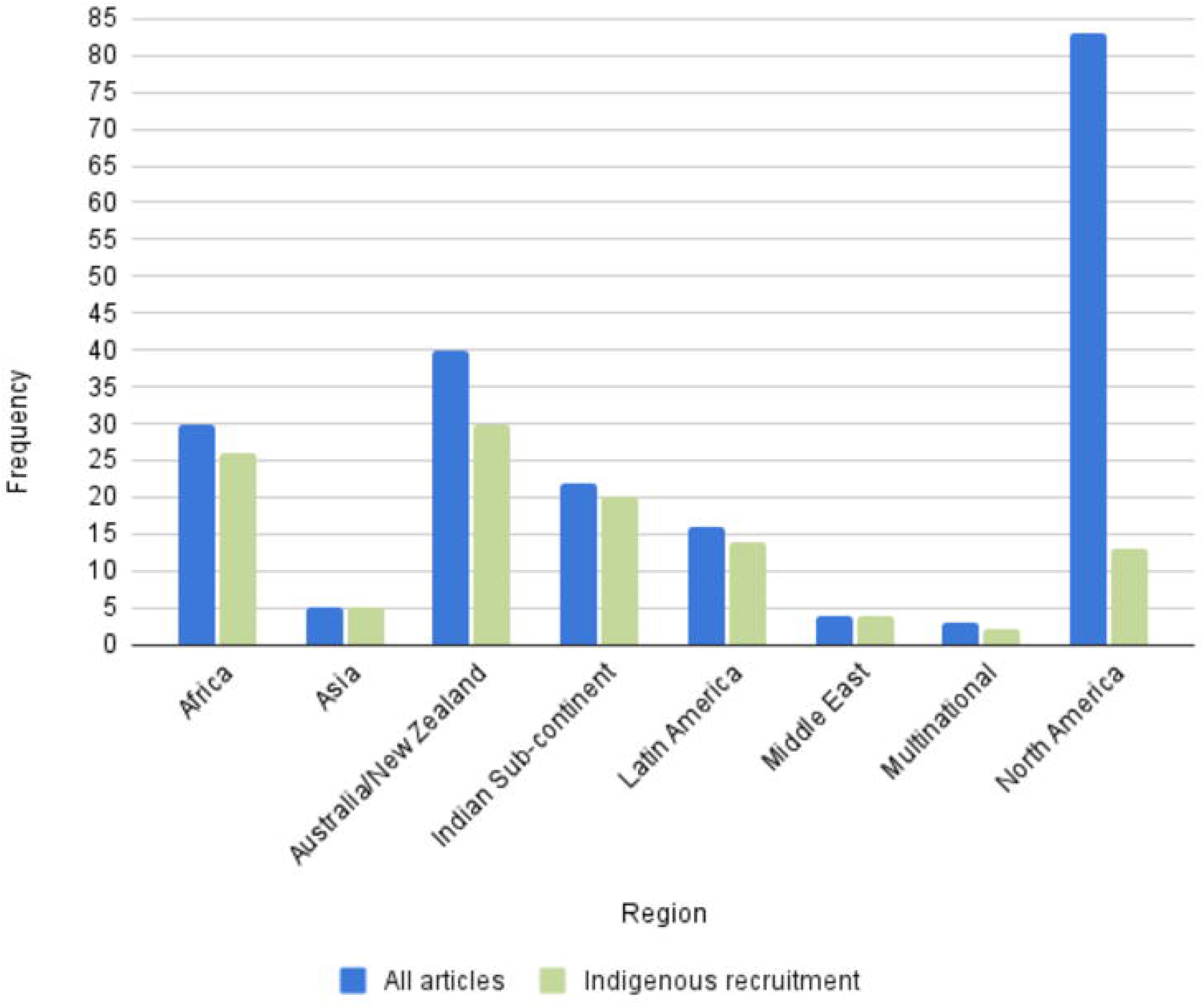
Regional representation of included publications.

**Fig 3.**
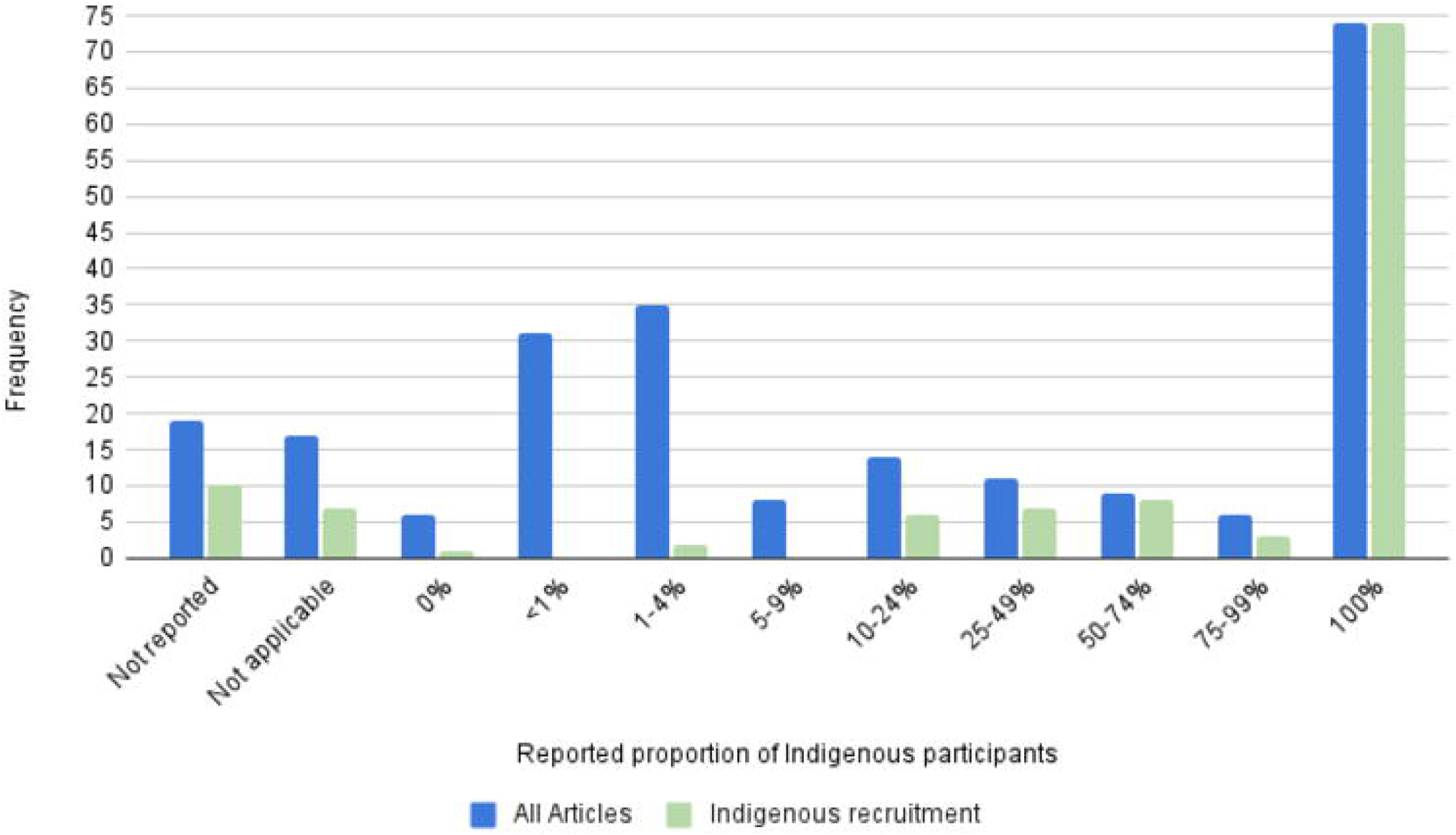
Frequency of reporting of the proportion of Indigenous participants.

Approximately 56% of references specifically recruited Indigenous peoples (n=114). These references were then isolated and underwent subsequent analysis. Per **Fig 2**, the distribution of publication region was highest in Australia/New Zealand (26.3%), followed by Africa (22.8%), and the Indian sub-continent (17.5%). As with the original sample, the majority of publications were from the primary literature (83%). The methodology of the publications was distributed as such: quantitative (48.2%), ethnobotanical/zoological (21.1%), qualitative (9.6%), reviews (systematic, scoping, narrative, etc. – 8.8%), mixed methods (4.4%), other (1.8%). The remaining publications (6.1%) either did not report the methodology, or it was not applicable (e.g., news article). In this sample, the proportion of Indigenous participants was reported 120 times. Publications that specifically recruited Indigenous populations, and attained 100% of Indigenous participants, were in the majority (62.5%). This differed from the initial sample, as illustrated in **Fig 3**. One publication did not recruit Indigenous participants, but spoke with non-Indigenous practitioners who provided services to Indigenous peoples.

### Demographics

Women/females were represented in 44.7% of publications, whereas two-spirit people were represented in <1%. Men were included in 10.5% of the publications, 28.9% included men and women, and 14.9% did not report on sex or gender or it did not apply (i.e., the publication reported on service delivery for Indigenous populations). The reported age of Indigenous participants varied, both in the method of reporting (e.g., median, mean, range) and in the represented ages (children to elders). Analysis was challenging, due to this variability in reporting. We opted to aggregate the data into categories (child, youth, adult, Elder) based on our best estimate on what was reported and extracted. Not all publications reported age, leaving 79 publications for analysis. The majority (91.1%) of those publications recruited adults (ages 19-59), which includes articles that reported age ranges extending into youth (ages 12-18) and Elder (ages 60+) categories. Only 6.3% only recruited Elders, and 2.5% only recruited children (ages 0-11) or youth.

### Safety

The use of trauma-informed approaches/frameworks and cultural safety practices was analyzed. Of the analyzed articles (n=114), 2.6% reported using a trauma-informed approach or framework and 25.4% mentioned the use of culturally safe practices.

### Pelvic health information

Of the articles that were interventional or described treatment options (n=38), the majority (65.8%) described herbal or traditional medicines, 13.2% mentioned surgical treatment, 5.3% education, 5.3% a combination of Western and traditional medicines, and the remainder mentioned medications, multiple treatment options, social support, or did not report the intervention (2.6% each).

**Fig 4** visualizes the frequency of pelvic health conditions mentioned in the literature that focused specifically on Indigenous peoples. One publication did not report a pelvic health condition but spoke to pelvic health physiotherapy service delivery to Australian Indigenous peoples. It was removed from this analysis (n=113). Since many publications reported multiple pelvic health conditions, the total frequency is higher than the number of publications. Bladder conditions were reported most frequently (47.1%), followed by pain conditions (18.4%), pelvic organ prolapse (11.5%), and erectile dysfunction (10.3%).

**Fig 4.**
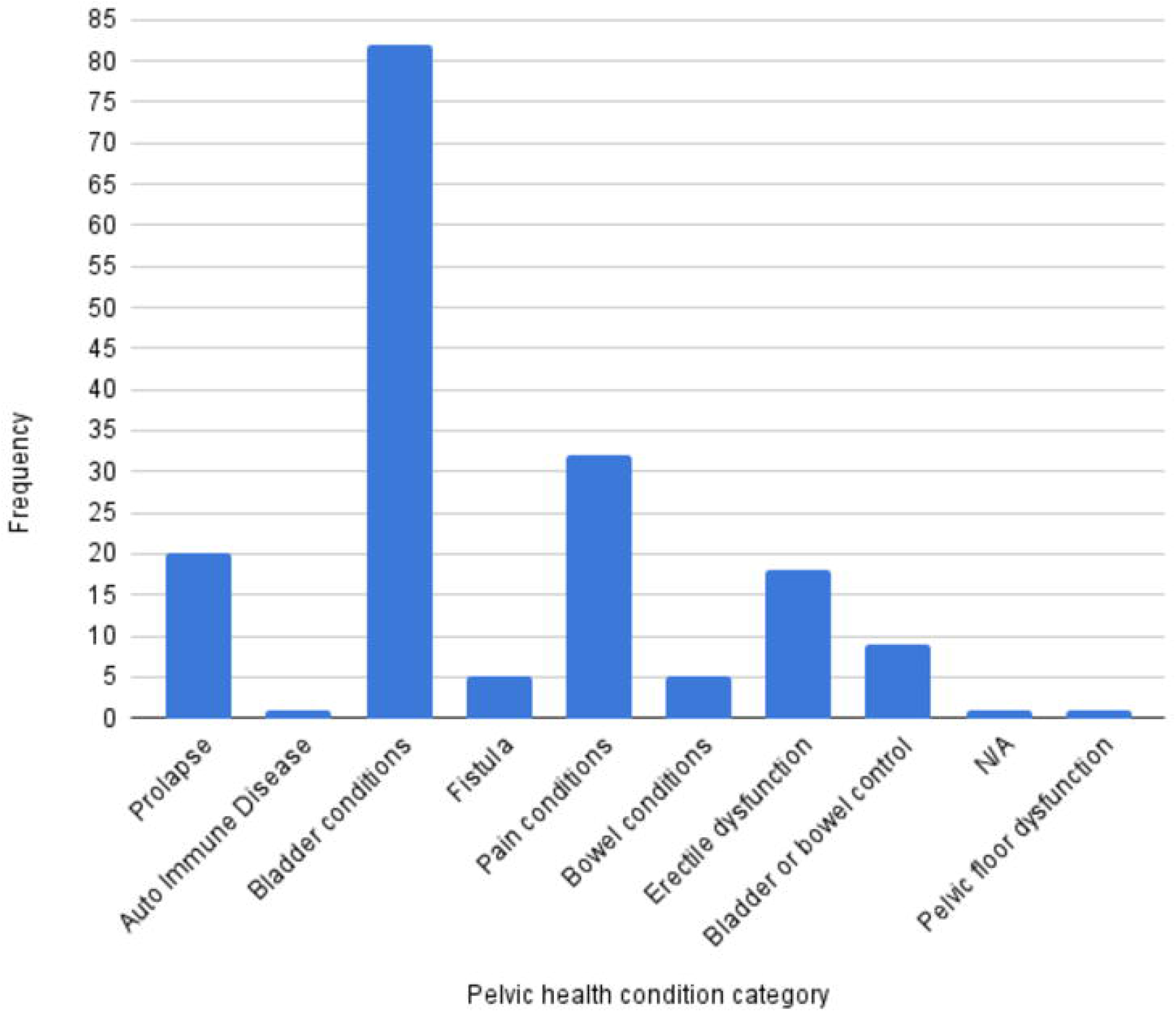
Frequency of pelvic health conditions described in the literature.

We also extracted epidemiological data, including prevalence of pelvic health conditions. Reports of urinary incontinence (including stress, urge, and mixed incontinence) ranged from 3-81%, dysuria 8.1-25.6%, bladder frequency 10.4-37.3%, pelvic organ prolapse 32-90%, dyspareunia 24.4-29%, dysmenorrhea 64.7-69.3%, and erectile dysfunction 3.6-61.5%. This variability can be explained by several factors including severity of symptoms (e.g., mild vs severe erectile dysfunction, grading of pelvic organ prolapse), life course (e.g., pregnancy vs post-partum, peri-vs post-menopausal), structure causing the symptoms (e.g., uterocele, cystocele, rectocele), and symptom presentation (e.g., urge incontinence, stress incontinence, mixed incontinence). There was one report of accidental bowel leakage (fecal incontinence) experienced by 2% of older Indigenous adults [52].

### Help-seeking and service delivery

Barriers and facilitators to help-seeking and service delivery were identified in the publications. Cultural norms and traditions in some cases were facilitators to accessing treatment, such as remedies rooted in Indigenous knowledges (9 publications), or in one publication, opportunities to seek care outside of community for Indigenous men. In other cases, cultural norms were a barrier to seeking help (5 publications). Privacy, gender roles and expectations, and societal perceptions were reported as limiting factors.

Several publications described the influence of the healthcare system and its staff, in many cases, posing a barrier to access. Service costs, location of services in relation to communities, healthcare staff attitudes and behaviour, timeliness of service, and staff competencies were reported as barriers. Two publications reported efforts to facilitate access to services, either by expanding the scope of practitioners or by addressing bureaucratic barriers experienced by practitioners.

Finally, emotions, knowledges, and beliefs of the participants in relation to help-seeking and service delivery were also described as barriers and facilitators. Shame, shyness, stigma, embarrassment, and self-perception were barriers (7 publications). Four publications described active knowledge seeking, and participants expressing interest in accessing services or learning more about pelvic health conditions as facilitators.

### Perceptions of pelvic health conditions

Similar themes regarding emotions and knowledge seeking arose around how people perceived pelvic health conditions. Unpleasant emotions such as embarrassment and shame were reported (4 publications). Knowledge seeking and awareness was identified as a need (4 publications) and in some cases, was being addressed by researchers, health care providers, and Indigenous community members (5 publications).

Participants reported several reasons for their symptoms (6 publications), with some rooted in spiritual beliefs (3 publications). Moreover, personal, interpersonal, and societal impacts of these pelvic health conditions were described, such as leading to tension within marriages (2 publications), need for family and community support (4 publications), migration away from communities to larger centres for treatment (1 publication), loss of fertility (2 publications), reduction in quality of life (1 publication), health outcomes (2 publications), and participation in work and community (2 publications).

### Commentary on healthcare systems

Functioning of the healthcare system was another theme. Experiences seeking treatment were described, many of which shared stories of system barriers and mal-treatment by healthcare providers (5 publications). Gaps in services and training were identified (5 publications). Solutions around provision of accessible and safe treatment were also described (2 publications).

## Discussion

This review systematically searched the literature to understand what is currently known about pelvic health conditions experienced by Indigenous peoples worldwide. The findings of this review will be summarized and discussed within the PAGER framework proposed by Bradbury-Jones et al [1].

### Patterns

Much of pelvic health literature originated from English-speaking regions – namely North America, Australia, and New Zealand. However, when we examined articles that specifically recruited Indigenous peoples, North American publications moved from most frequent to mid-range. To our knowledge, international trends around the country/region of publication origin has not been explored in the pelvic health literature.

Women (i.e., sex reported as female or gender reported as woman) participated in more studies than men (i.e., sex reported as male or gender reported as man) or gender diverse people. To our knowledge, there has been no study examining sex and gender representation within the pelvic health literature.

This finding is not surprising, considering that pelvic health conditions such as incontinence are more prevalent in women [14,53], female specific conditions (e.g., endometriosis, pelvic organ prolapse) were more frequent in our data, and that our journal hand search focused on publications relating to women’s health Bladder conditions were most frequently reported in the publications, whereas autoimmune diseases, bowel conditions, and fistula were least frequently reported. As in the broader literature, this review also demonstrated variability in prevalence rates of pelvic health conditions. For example, Vogel et al [15] report post-partum urinary incontinence ranging between 8-31%, Shaw et al [14] report 23.7% of Canadian adults, and Peinado-Molina et al [17] report 55.8% experienced by adult women in Spain. Our study reports urinary incontinence prevalence from 3-81%. Only one publication in our study reported the prevalence of fecal incontinence experienced by older Indigenous adults at 2% [52]. A recent systematic review and meta-analysis identified a global prevalence rate of 8.0% [54]. The prevalence of pelvic organ prolapse within the included studies (32-90%) was higher than in other publications. Palmieri et al [16] found a prevalence of 6.1% in a population of pregnant and post-partum Italian women. Other authors report a range from 3.4-10.76% in adult American women [55], 14% in adult Spanish women [17], and 22.7% in Ethiopian women [56]. Rates of dyspareunia identified in our study (24.4-29%) were higher than a sample of women from the United Kingdom at 7.5% [57], but were within the range reported by other authors who explored pregnancy and post-partum (22-44%) [58], and menopause (20-77.6%) [59].

Ethnobotanical and zoological studies were prevalent methodologies in the primary literature and were a high proportion of the interventional publications in this study. These publications described traditional medicine interventions for pelvic health conditions. By mentioning remedies for pelvic health conditions, the studies imply that Indigenous peoples in those groups experience symptoms, although the extent is not clear from the literature.

Emotional experiences such as shame, embarrassment, and stigma were reported by Indigenous peoples. These experiences were in response to having a pelvic health condition and posed a barrier to accessing support and healthcare services. There were numerous barriers to healthcare access identified, which included logistical challenges such as finances and travel requirements. Other reports of healthcare access for Indigenous peoples in Australia [4] and Canada [3] include geographic and economic barriers. We also identified pelvic health conditions’ impacts on quality of life, relationships, and participation within community. This aligns with existing literature on the negative effects of prolapse [60], pelvic pain conditions [20,61], and bladder incontinence [62] on quality of life.

### Advances

Representation of women is often lacking in biomedical research, and sex and gender is under-reported [63]. The majority of publications in our study reported on sex and gender. There was also a high percentage of women/female participants compared to men/males.

A surprising finding of this review was the number of ethnobotanical and zoological studies. These publications came from the primary literature and reported on the application of traditional knowledge within Indigenous communities to treat pelvic health conditions, by using substances derived from plants and animals. As summarized by Saini [64], Western science has been dismissive of Indigenous ways of knowing, which includes traditional medicines. Additionally, access to Western medicine can challenging, and Indigenous peoples may turn to traditional remedies, which are more accessible and align with their cultural values [65–68]. Overall, the publications in this review explored the depth of Indigenous knowledges and did not report on the efficacy of the substances.

### Gaps

Among the regions represented in the literature, there were noticeable gaps from regions with known high proportions of Indigenous peoples. Alaska, USA and northern Canada were represented with a small number of publications. The Canadian territories of the Northwest Territories, Nunavut, and Yukon report the highest proportions of Indigenous peoples in the country, at approximately 50% [69], 86% [70], and 22% [71], respectively. Alaska is reported to have the highest proportion of American Indians and Alaska Natives in the United States of America (22%) [72]. There were also no publications from Iceland, Greenland, Norway, Sweden, Finland, Russia, several of which are known to be home to Indigenous peoples (e.g., Kalaallit from Greenland, and Sámi peoples from Norway, Sweden, Russia, and Finland). Finally, China was missing from the review, which does not recognize Indigenous populations [73].

Upon analysis of the reported age and gender, gaps were also revealed in terms of participant demographics. Although women were represented more in this scoping review, there was a paucity of publications relating to gender diverse Indigenous people. One publication mentioned two-spirit gender identity. Additionally, a gap exists in research concerning Indigenous children (0-12 year olds), youth (13-18 year olds), and elders (60+). Pelvic health conditions increase in prevalence as the population ages [17,54] and children and youth can experience different conditions from adults and elders (such as bedwetting and encopresis).

Reporting of certain pelvic health conditions, such as auto-immune diseases (like lichens sclerosis) and bowel conditions was lacking. Perhaps this is due to known underreporting of bowel conditions [74], or due to low prevalence of auto-immune conditions [75].

No publications investigated the economic impacts of pelvic health conditions experienced by Indigenous populations. Other publications have reported substantial costs to the general population and the systems that serve it. Self-management including medications and products can range $1400 – 2100 CAD annually for a senior living with incontinence, per The Canadian Continence Foundation [76]. Costs incurred by insurers and healthcare systems include medications, medical devices, personnel, and surgery [77]. Urinary incontinence alone is estimated to cost the Canadian healthcare system $3.84 billion annually [76].

Finally, few publications mentioned the use of cultural safety and/or trauma-informed approaches. Whether this is due to under-reporting, or under-utilization remains unclear. Limitations in the existing literature may have contributed to these results. Although used widely in healthcare settings, cultural safety within research has not been explored extensively in the existing literature [43]. Gaps also exist in the literature relating to gold standard approaches to trauma-informed research [78].

### Evidence for Practice

Pelvic health conditions are prevalent in the general population. As several authors [22–24] have proposed, homogeneity and under-representation of Indigenous people and people of colour in the pelvic health literature may have consequences on clinical decision making. We argue that this may also extend to policy decisions and thereby impact service delivery to Indigenous populations. The current literature demonstrates that pelvic health conditions are experienced by Indigenous peoples at comparable prevalence rates to the general population. Recognizing that Indigenous people may live in remote regions [3,4], where delivery of health services can be challenging and costly, decision makers need to consider novel approaches to increase equity of access.

Discussing pelvic health conditions may be perceived as a private issue, and there may be stigma or cultural norms that health care practitioners should be aware of when working with Indigenous peoples. Creating culturally safe environments within the healthcare system may be one way in which practitioners and the systems they work in can improve quality of and access to care.

### Recommendations for Research

Jefferson et al [78] identified existing gaps with research best practices while working with trauma-exposed populations, and noted the under-representation of marginalized populations. The authors recommend that gold-standard research guidelines be developed, implemented, and tested, and that trauma-exposed individuals and communities participate in guideline development. We agree that additional research is needed in partnership with Indigenous peoples to develop and implement research practices that promote participant safety and are rooted in Indigenous epistemologies. This may be achieved by using culturally safe and trauma-informed approaches.

There were disparities in the literature in sex and gender, age, and type of pelvic health conditions reported. Since this review has demonstrated that women and females were more represented, future research in pelvic health conditions should emphasize including men and gender diverse participants. Additional research is also needed to explore the experiences of Indigenous children, youth, and Elders. Furthermore, under-studied pelvic health conditions included bowel conditions, prolapse, auto-immune disorders, and erectile dysfunction. In regions where there is an abundance of pelvic health research relating to urinary incontinence, for example, we recommend shifting focus towards these under-studied conditions.

### Limitations

Mental wellness and pelvic health conditions are interrelated. Depression and/or anxiety are associated with both urinary and fecal incontinence [62,79,80], interstitial cystitis [61], and pelvic organ prolapse [60]. Women reporting dyspareunia (painful intercourse) can also experience depression, and dissatisfaction or distress with their sex life [57]. We did not actively search for reports or descriptions of mental health diagnoses in our analysis, and as such can make no conclusions about Indigenous peoples’ experiences with these often co-occurring experiences.

There are challenges developing a search string representative of Indigenous peoples globally, such as a lack of a universal definition, which leaves researchers to their own devices to develop search terms, resulting in bias and gaps in knowledge [81]. As such, we used the United Nations’ description of Indigenous peoples to inform search development and screening criteria. Also, replication of previously used, but inadequate, search terms can perpetuate bias [81]. To address these challenges, and in an attempt to be as comprehensive as possible, a university librarian (SMC) was recruited to develop and run the online database searches. There were other limitations identified during development of the search and sorting of the publications. Firstly, some countries (such as China), do not acknowledge Indigenous peoples’ existence within their borders [73] and as such, there is no reporting on their experiences. Secondly, we only included publications in English, which may have resulted in under-reporting from regions that primarily publish in other languages (e.g., Russia, Latin America, etc.). Finally, some regions (such as Sweden) do not collect data on race or ethnicity [82], which poses barriers to researchers describing and reporting about Indigenous peoples’ experiences.

Several authors [25,26,83] either propose or stress the importance of the final stage of a scoping review: consultation. We agree that this stage is important to contextualize the findings and increase the rigor of the literature review [26]. However, we believe consultation should include the people we are talking about. Consultation with Indigenous peoples requires intentional engagement, focused on building trust and developing relationships [84,85]. This is a resource-intensive endeavor, and the timeline for engagement and consultation falls outside of typical scoping review publication timelines. As such, we plan to incorporate the findings of this review into an entire study dedicated to understanding Indigenous women’s experiences with pelvic health conditions in northern Canada using community-based participatory principles and a trauma-informed framework. The findings of this work will be shared in future publications.

## Supporting information

S1 Appendix. Primary literature database search strategies.

S2 Checklist. PRISMA-S checklist.

S3 Dataset. Dataset extracted from included full texts.

## Data Availability

All relevant data are within the manuscript and its Supporting Information files.

## Authors’ contributions

KB conceptualized the study. SMC developed the search strategies for both pelvic health conditions and Indigenous populations, ran the online database searches, and imported the results into Covidence. KC and EK screened the results and participated in data extraction. SC, PM, and JS reviewed and contributed to the pelvic health conditions list used to develop the search strategies. SC supervised the study. All authors contributed to reviewing and editing the submitted manuscript.

## Supporting Information

**S1 Appendix. Primary literature database search strategies.**

**S2 Checklist. PRISMA-S checklist.**

**S3 Dataset. Dataset extracted from included full texts.**

