## Supplementary material for "Experiences of Indigenous peoples living with pelvic health conditions: A scoping review": S1 Appendix. Primary literature database search strategies.

**Ovid MEDLINE(R) ALL <1946 to February 13, 2024>**

| **1** | **urinary incontinence/ or urinary incontinence, stress/ or urinary incontinence, urge/** | **36731** |
| --- | --- | --- |
| **2** | **exp Fecal Incontinence/** | **11031** |
| **3** | **pelvic organ prolapse/ or cystocele/ or rectal prolapse/ or uterine prolapse/ or visceral prolapse/** | **14524** |
| **4** | **Urinary Bladder, Overactive/** | **6085** |
| **5** | **exp Urinary Retention/** | **5292** |
| **6** | **Constipation/** | **16416** |
| **7** | **Encopresis/** | **672** |
| **8** | **Pelvic Floor Disorders/** | **1320** |
| **9** | **Pelvic Girdle Pain/** | **228** |
| **10** | **exp Dyspareunia/** | **2537** |
| **11** | **exp Vulvodynia/** | **586** |
| **12** | **exp Vaginismus/** | **179** |
| **13** | **exp Cystitis, Interstitial/** | **2673** |
| **14** | **Adenomyosis/** | **1343** |
| **15** | **Pudendal Neuralgia/** | **147** |
| **16** | **exp Gynecologic Surgical Procedures/** | **92595** |
| **17** | **exp Colectomy/** | **23829** |
| **18** | **exp Rectal Neoplasms/** | **55324** |
| **19** | **exp Endometrial Neoplasms/ or exp Uterine Neoplasms/ or exp Genital Neoplasms, Female/ or exp Ovarian Neoplasms/** | **261190** |
| **20** | **exp Prostatic Neoplasms/** | **151820** |
| **21** | **exp Prostatectomy/ or exp Prostatic Neoplasms/** | **166910** |
| **22** | **((gyn?eologic* or rectal or rectum or genital or prostat* or unterine or uterus or ovarian or endometrial) adj3 (surgery or surgical)).mp. [mp=title, book title, abstract, original title, name of substance word, subject heading word, floating sub-heading word, keyword heading word, organism supplementary concept word, protocol supplementary concept word, rare disease supplementary concept word, unique identifier, synonyms, population supplementary concept word, anatomy supplementary concept word]** | **20824** |
| **23** | **94 or 95 or 96 or 97 or 98 or 99 or 100** | **572807** |
| **24** | **exp Postoperative Complications/ or (postoperative or "post operative" or "post surg*").mp.** | **1152646** |
| **25** | **101 and 102** | **60603** |
| **26** | **exp Urinary Bladder, Neurogenic/** | **8005** |
| **27** | **sexual dysfunction, physiological/ or erectile dysfunction/ or impotence, vasculogenic/** | **31125** |
| **28** | **exp Nocturnal Enuresis/** | **1450** |
| **29** | **Encopresis/** | **672** |
| **30** | **exp Hirschsprung Disease/** | **5046** |
| **31** | **Sex Reassignment Surgery/** | **935** |
| **32** | **Lichen Sclerosus et Atrophicus/** | **1412** |
| **33** | **exp Nocturia/** | **1192** |
| **34** | **urinary fistula/ or urinary bladder fistula/ or vaginal fistula/ or rectovaginal fistula/ or vesicovaginal fistula/** | **10151** |
| **35** | **Pelvic Floor/** | **6974** |
| **36** | **lower urinary tract symptoms/ or dysuria/ or nocturia/ or prostatism/ or urinary bladder, overactive/ or urinary bladder, underactive/ or urinary incontinence/ or urinary incontinence, stress/ or urinary incontinence, urge/** | **46934** |
| **37** | **79 or 80 or 81 or 82 or 83 or 84 or 85 or 86 or 87 or 88 or 89 or 90 or 91 or 92 or 93 or 103 or 104 or 105 or 106 or 107 or 108 or 109 or 110 or 111 or 112 or 113 or 114** | **199274** |
| **38** | **(prolaps* adj3 ("pelvic organ" or uterine or uterus or bladder or rectal or rectum or urethra* or apical)).mp.** | **18200** |
| **39** | [**incontinence.mp**](http://incontinence.mp/)**.** | **67979** |
| **40** | **(uterocele or cystocele or rectocele or enterocele or urethrocele or sigmoidocele).mp. [mp=title, book title, abstract, original title, name of substance word, subject heading word, floating sub-heading word, keyword heading word, organism supplementary concept word, protocol supplementary concept word, rare disease supplementary concept word, unique identifier, synonyms, population supplementary concept word, anatomy supplementary concept word]** | **3070** |
| **41** | **(bladder* adj3 (overactive or urgency or urgent or frequency or heistency or retention)).mp. [mp=title, book title, abstract, original title, name of substance word, subject heading word, floating sub-heading word, keyword heading word, organism supplementary concept word, protocol supplementary concept word, rare disease supplementary concept word, unique identifier, synonyms, population supplementary concept word, anatomy supplementary concept word]** | **11036** |
| **42** | **(bowel adj3 (frequency or urgency)).mp. [mp=title, book title, abstract, original title, name of substance word, subject heading word, floating sub-heading word, keyword heading word, organism supplementary concept word, protocol supplementary concept word, rare disease supplementary concept word, unique identifier, synonyms, population supplementary concept word, anatomy supplementary concept word]** | **1476** |
| **43** | [**constipation.mp**](http://constipation.mp/)**.** | **35221** |
| **44** | [**encopresis.mp**](http://encopresis.mp/)**.** | **908** |
| **45** | **(dyssynergia adj3 (bowel* or bladder*)).mp.** | **149** |
| **46** | **"vaginal wind".mp.** | **10** |
| **47** | **"pelvic girdle pain".mp.** | **465** |
| **48** | **(dyspareunia or vulvodynia or vaginismus or vestibulodynia or "interstitial cystitis" or "painful bladder syndrome" or "proctalgia fugax" or prostatitis or endometriosis or adenomyosis or "pudendal neuralgia" or anismus).mp. [mp=title, book title, abstract, original title, name of substance word, subject heading word, floating sub-heading word, keyword heading word, organism supplementary concept word, protocol supplementary concept word, rare disease supplementary concept word, unique identifier, synonyms, population supplementary concept word, anatomy supplementary concept word]** | **54523** |
| **49** | **Neurogenic** [**bladder.mp**](http://bladder.mp/)**.** | **5130** |
| **50** | **("sexual dysfunction" or "erectile dysfunction" or "persistent genital arousal" or impostence).mp. [mp=title, book title, abstract, original title, name of substance word, subject heading word, floating sub-heading word, keyword heading word, organism supplementary concept word, protocol supplementary concept word, rare disease supplementary concept word, unique identifier, synonyms, population supplementary concept word, anatomy supplementary concept word]** | **44459** |
| **51** | **("bed wett*" or bedwett* or encopresis or "hirshprung* disease").mp. [mp=title, book title, abstract, original title, name of substance word, subject heading word, floating sub-heading word, keyword heading word, organism supplementary concept word, protocol supplementary concept word, rare disease supplementary concept word, unique identifier, synonyms, population supplementary concept word, anatomy supplementary concept word]** | **1550** |
| **52** | **("gender confirmation surg*" or "sex reassignment surg*").mp. [mp=title, book title, abstract, original title, name of substance word, subject heading word, floating sub-heading word, keyword heading word, organism supplementary concept word, protocol supplementary concept word, rare disease supplementary concept word, unique identifier, synonyms, population supplementary concept word, anatomy supplementary concept word]** | **1289** |
| **53** | **"lichen sclerosis".mp.** | **181** |
| **54** | [**nocturia.mp**](http://nocturia.mp/)**.** | **4304** |
| **55** | **((fistula* or fistulae) adj3 (bladder* or urinary or vagina* or rectovaginal* or vesicovaginal*)).mp. [mp=title, book title, abstract, original title, name of substance word, subject heading word, floating sub-heading word, keyword heading word, organism supplementary concept word, protocol supplementary concept word, rare disease supplementary concept word, unique identifier, synonyms, population supplementary concept word, anatomy supplementary concept word]** | **12891** |
| **56** | **"levator ani avulsion".mp.** | **57** |
| **57** | **("Lower urinary tract symptom*" or dysuria or "LUTS").mp. [mp=title, book title, abstract, original title, name of substance word, subject heading word, floating sub-heading word, keyword heading word, organism supplementary concept word, protocol supplementary concept word, rare disease supplementary concept word, unique identifier, synonyms, population supplementary concept word, anatomy supplementary concept word]** | **17977** |
| **58** | **"voiding dysfunction".mp.** | **2679** |
| **59** | **or/115-136** | **305875** |
| **60** | **((exp Indians, North American/ and Canad*.mp.) or Indigenous Canadians/ or exp Inuits/ or exp Health Services, Indigenous/ or exp Ethnopharmacology/ or (Athapaskan or Saulteaux or Wakashan or Cree or Dene or Inuit or Inuk or Inuvialuit* or Haida or Ktunaxa or Tsimshian or Gitxsan or Gitksan or "Nisga'a" or Haisla or Heiltsuk or Oweenkeno or "Kwakwaka'wakw" or "Nuu chah nulth" or "Tsilhqot'in" or Dakelh or "Wet'suwet'en" or Sekani or Dunne-za or Dene or Tahltan or Kaska or Tagish or Tutchone or Nuxalk or Salish or St'at'imc or Stl'atl'imx or Stl'atl'imc or Nlaka'pamux or Okanagan or "Sec wepmc" or Secwepemc or Tlingit or Anishinaabe or Blackfoot or Nakoda or Tasttine or "Tsuu T'ina" or "Tsuut'ina" or "Gwich'in" or (Han not (China or Chinese)) or Algonquin or Nipissing or Ojibwa or Potawatomi or Innu or Maliseet or "Mi'kmaq" or Micmac or Passamaquoddy or Haudenosaunee or Cayuga or Mohawk or Oneida or Onondaga or Seneca or Tuscarora or Wyandot or Aboriginal* or Indigenous* or Metis or red road or "on reserve" or off-reserve or First Nation or First Nations or Amerindian).mp. or (urban adj3 (Indian* or Native* or Aboriginal*)).mp. or** [**ethnomedicine.mp**](http://ethnomedicine.mp/)**. or country food*.mp. or residential school*.mp. or ((exp Medicine, Traditional/ or traditional medicine*.mp.) not Chinese.mp.) or exp Shamanism/ or shaman*.mp. or traditional heal*.mp. or traditional food*.mp. or medicine** [**man.mp**](http://man.mp/)**. or medicine** [**woman.mp**](http://woman.mp/)**. or autochtone*.mp. or (Native* adj1 (man or men or women or woman or boy* or girl* or adolescent* or youth or youths or person* or adult or people* or Indian* or Nation or tribe* or tribal or band or bands)).mp.) and (exp Canada/ or (Canad* or British Columbia or Colombie Britannique or Alberta or Saskatchewan or Manitoba or Ontario or Quebec or Nova Scotia or New Brunswick or Newfoundland or Labrador or Prince Edward Island or Yukon Territory or NWT or Northwest Territories or Nunavut or Nunavik or Nunatsiavut or NunatuKavut).mp.) [Canada]** | **8926** |
| **61** | **((exp australia/ or (australia* or northern territory or tasmania or new south wales or Victoria or queensland).ti,ab.) and ("Native Hawaiian or Other Pacific Islander"/ or aborigin*.ti,ab. or indigenous.ti,ab.)) or "torres strait islander*".ti,ab. [Australia and Torres Strait]** | **12869** |
| **62** | **(Maori or "tangata whenua").mp. or ((exp New Zealand/ or (New Zealand or Aukland).mp.) and ("Native Hawaiian or Other Pacific Islander"/ or (Aborig* or Indig*).ti,ab.)) [Maori]** | **5839** |
| **63** | **(Saami or Sampi or (Sami not Ulus) or Samis or Southernsami* or Umesami* or Pitesami* or Lulesami* or Northernsami* or Enaresami* or Kolasami* or Lapp or Lapps or Lappish or Lappland or (Lapland* not longspur) or Lappalainen* or Saamelainen* or reindeer herd* or reindeer culture* or reindeer pastoral* or Lappbys or Samebys or reinbeitesdistrikt or paliskunta or siida).mp. or (((Fennoscandia or Finnmark or Scandinavia or Nordic or Sweden or Norway or Finland or Swedish or Finnish or Norwegian or Norge or Svensk* or Suomi or Barents Region or (Kola not (garcinia or gotu)) or Arctic Europe* or Polar Europe* or North* Europ*).mp. or Finland/ or Norway/ or Sweden/) and ((traditional adj3 (food* or heal* or medicine* or shaman*)) or (Indigenous* adj3 (people* or person* or mother* or father* or parent* or grandparent* or grandmother* or grandfather* or elder or elders or child* or boy or boys or girl* or youth* or healer* or patient or patients or famil* or herder* or village* or communit*))).mp.) [Sami]** | **1683** |
| **64** | **((exp Indigenous People/ or "indigenous people* ".mp.) and (exp Africa/ or ((Africa* not "African American*") or Algeria or Angola or Benin or Botswana or "Burkina Faso" or Burundi or Cameroon or "Cape Verde" or "Cabo Verde" or "Central African Republic" or Chad or Comoros or Congo or Djibouti or Egypt or "Equitorial Guinea" or Eritrea or Eswatini or Ethiopia or Gabon or Gambia or Ghana* or Guinea or "Guinea Bissau" or "Ivory Coast" or Kenya or Lesotho or Liberia or Libya or Madagascar or Malawi or Mali or Mauritania or Mauritius or Mayotte or Morocco or Mozambique or Namibia or Niger or Nigeria or Reunion or Rwanda or "Saint Helena" or Ascension or "Tristan de Cunha" or "Sao Tome" or Principe or Senegal or Seychelles or "Sierra Leone" or Somalia or "South Africa" or Sudan or Tanzania or Togo or Tunisia or Uganda or "Western Sahara" or Zambia or Zimbabwe).mp.)) or (Abakuria or Abaluhya or Abagusii or Abakuria or Aembu or Agikuyu or Akamba or Anuak or Anywaa or Amazigh or Ambala or Ambeere or Ambundu or Ambuun or Amharan or Angba or Baaka or Baamba or Babindi or Babini or Baboma or Bachokwe or Bacwa or Bafumbira or Baganda or Bagyele or Bagwere or Bagyeli or Bakiga or Bakola or Baholo or Bakalanga or Bakiga or Bakolo or Bakongo or Bakonjo or Baluba or Balunda or Balovale or Bamasaba or Bambuti or Bangala or Bangoli or Bangungu or Bantu or Banyankole or Banyarwanda or Banyole or Banyoro or Bapende or Bapedi or Barabaig or Barombi or Barundi or Baruuli or Basamia or Basoga or Batswana or Batooro or Batsamba or Batswana or Batwa or Bayaka or Bedzan or Bazombe or Bebayaka or Bedzan or Bhaca or Biaka or Borana or Chewa or Copts or Cormorian or Cushitic or Dahalo or Datooga or Dikidiki or Dogon or Ewondo or Fulani or Fuliru or Ganguela or Gciriku or Gyele or Hadza or Hadzabe or Haillom or Haratin or Herero or Himba or Hlubi or Iriryen or Iqvayliyen or Kabyle* or Kalenjin or Kanioka or Kanioka or Kaonde or Karamojong or Kavango or Kereuyu or Khoikhoi or KhoiSan or Kikuyu Kwangali or Lokele or Lowme or Lotuko or Lwalwa or Maasai or Makonde or Makua or Mande or Masalit or Matumbi or Mayeuyi or Mayeyi or Mbenga or Mbukushu or Mbochi or Mboro or Mbuti or Medzan or Mijikenda or Mozabite* or Nafusa or Ndebele or Ngombe or Namaqua or Nyanga or Nyamwezi or Ogiek or Ovambo or Ovimbundu or Phuthi or Pokomo or Rendille or Riffian or Riffians or Sakuma or Samburu or Sandawe or Sangha or Sango or Sengwer or Serer or Sesotho or Shangaan or Shawiya or Shenwa or Shi or Shilluk or Sukua or Sukus or Swahili or Tabwa or Tambuka or Taveta or Thembu or Tigrayan or Topoke or Tsonga or Toubou or Tuareg or Tumbuka or Ugana or Wochua or Xhosa or Xindonga or Yoruba or Zenati or Zuwara or ((Indigenous or Afar or Afars or "Aka People" or Akie or Ameru or Asua or Ateker or Atwot or Awjila or Bafia or Baka or Bakongo or Bakwe or Balunda or Balovale or Bango or Bassa or Beja or Bekpak or Bemba or Bembe or Benet or Berber or Berbers or Bira or Bowe or Bubi or Budja or Bulu or Bunrun or Chaga or Chopi or Damara or Dinka or Djerba or Duala or Dzing or Efe or Elmolo or Fang or Foora or Fula or Fur or Ghomara or Ghadames or Gllana or Glu or Gogo or Gongo or Haya or Havu or Hema or Hima or Hunde or Hutu or Huva or Iboko or Igbo or Ijo or Jieng or Kadu or Kande or Kango or Katla or Kgaga or Khoe or Kola or Komo or Kota or Kua or Kuba or Kwango or Kx'z or Kxoe or Lala or Lozi or Luo or Luba or Lupu or Masmuda or Matmata or Mbala or Mbam or "Mbo People" or Mbolo or Mbuza or Mongo or Mpondo or Myene or Naadh or Nama or Nande or Naro or Ngoni or Ndau or Ndebele or Ngoli or Ngondi or Ngoni or Nguni or Nkoya or Nkumu or Nuba or Nubian or Nuer or Nzebi or Ogoni or Omoro or Oroko or Pygmy or Popoi or Poto or Puru or Rashad or (San not ("San Francisco" or "San Diego" or "San Antonio")) or Sango or Sanhaja or Sena or Shilha or Shira or Shona or Shua or Sokna or Somali* or Sotho or Sua or Subu or Swazi or Taitaa or Tchokwe or Teke or Tembo or Tetela or (Tonga and Africa*) or Tshwa or Tsoa or Twa or Turkana or Tuu or Venda or Vira or Watta or Wakuti or Yaaku or Yaka or Yakoma or Yanzi or Yao or Yeke or Yela or Yeyi or Zulu) adj3 (population* or people* or person* or elder* or man or men or woman or women or child* or youth* or clan or clans or tribe or tribes or tribal or family or families or parent* or grandparent* or elder or elders or grandmother* or grandfather* or baby or babies or infant or infants or patient or patients or speakers or speaking or village* or communit*))).mp. [Africa]** | **32246** |
| **65** | **(((Acatec or Aguacateco or Amuzgo or Bokota or Boruca or Bribri or "Bri Bri" or Buglere or Cabecar or Cakchiquel or Changuena or Chatino or Chiapanec or Chicomuceltec or Chinantee or Chocho or Cholti or "Ch'olti'" or "Ch'olti'anor Chontal" or Chorotega or Chorti or Chuj or Chumbia or Corobici or (Cueva not Spain) or Cuicatec or Cuitlatee or Cuytec or Dorasque or Embera or Garifuna or Guatuso or Guaymi or Guaymis or Guetar or Huastec or Huave or Huetar or Itzaj or Ixil or Jacalteco or Jonaz or Kanjobal or Kekchi or Kuna or Maleku or Mangue or Matambu or Matlatzinca or Mazahua or Motozintlec or Mayan or Mayangna or Miskito or Mixtec or Mopan or Nahua or Nahuatl or Ngabe or Otomi or Pantec or Paya or Popoloca or Popoloc or Poqomam or Poqomchi or "Q'eqchi'" or Quiche or Quitirrisi or Sacapulteco or Sipacapense or Subtiaba or Tacaneco or Tarasco or Tamaulipec or Tamazultec or Tecoxquin or Tectiteco or Tecual or Tecuexe or Tepehura or Tepuztecor or Teribe or Terraba or Totonac or Trique or Tzeltal or Tzotzil or Tzutujil or Ulwa or Uspantec* or Uspanteko or Voto or Xinca or Waunana or Wounaan or Yucatec or Zapotec or Zoque or (("Costa Rica*" or Hondura* or Nicaragua* or Panama* or Guatemala* or Achi or Belize or Belizean* or Maya* or Mixe or Pame or Pipil or Pech or Chol or "Ch'olan" or Cora or Cuna or Mam or Rama) adj5 (Indian or Indians or Amerindian* or Amerindio* or Aboriginal* or Indigenous or Indigena* or Aborigen* or Mestizo or tribe or tribes or tribal or "traditional medicine*" or shaman*))).tw. or (Belize or "Costa Rica" or "El Salvador" or Guatemala or Honduras or Nicaragua or Panama).mp.) and (exp Indigenous Peoples/ or (indigenous adj3 (population* or people* or person* or elder* or man or men or woman or women or child* or youth* or clan or clans or tribe or tribes or tribal or family or families or parent* or grandparent* or elder or elders or grandmother* or grandfather* or baby or babies or infant or infants or patient or patients or speakers or speaking or village* or communit*)).mp.)) not ((Mexico or Mexican).mp. or exp Mexico/) [Central America]** | **736** |
| **66** | **Indigenous People/ or American Indian/ or Canadian Aboriginal/ or Eskimo/ or Inuit/ or Indigenous Health Services/ or ("A' ani" or Absaroka or Haaninin or Atsina or "Gros Ventre" or Acopsel or Tlacopsel or Lacopsel or Ahtna or Ahtena or Akenitsi or Occaneechi or Akokisa or Horcoquisa or Orcoquizas or Aleut or Unangax or Unangan or Alibamu or "Alabama Alsea" or Alutiiq or Sugpiag or Amahami or Awaxawi or Androscoggin or Arosaguntacook or Ameriscoggin or Anishinaabeg or Chippewa or Anihsinape or Saulteaux or Apalachee or Aranama or "Texan Coahuilteca" or Tamique or Arikara or Sahnish or Arickaree or Adakadaho or Assiniboine or Hohe or Nakota or Nakoda or Nakona or "Atsa' Kudok-wa" or Awatixa or Bannock or "Snake Indian*" or Bidai or Quasmigdo or Biloxi or Blackfoot or Niitsitapi or Sikasikaitsitapi or Cahto or Kaipomo or Cahuilla or Ivilyuqaletem or Ivilyuat or Catawba or Inna or Iswa or Chemehuevi or Chickasaw or "Chilula Chimakum" or Aqokulo or Chimariko or Chiricahua or Tsokanende or Chitimacha or Chetimachan or Sitimacha or Chowanoke or Roanoke or Chumash or Ciboney or "Taino Ciwat" or Clatsop or Coos or Coosa or Uchis or Chiaha or Coste or Talisi or Coquille or Kokwell or Coso or Cowlitz or Taitnapam or "Crow Nation" or "Cui Ui Ticutta" or Cupeno or Kuupangaxwichem or Cupa or "Cup' ig" or Nunivak or "Dakota Oyate" or Lakota or Nakota or Santee or Teton or Sioux or Deadose or "Deg Xina" or "Deg Xit' an" or Kaiyuhkhotana or "Deg Hit' an" or "Dena' ina" or Tanaina or "Dichinanek' Hwt' ana" or "Upper Kuskokwim Athabascan*" or Kolchan or Goltsan or "Tundra Kolosh" or "Do lkabya" or Duwamish or Esselen or Eyak or "Gidi' tikadi" or Guwevkabaya or "Gwich' in" or Kutchin or Haida or Xaadas or Xaat or Halchidhoma or Havasupai or "Green Water People" or Hiratsa or Hiraaca or "Ho-chaaqa" or Winnebago or Holikachuk or Innoko or "Tlegon-khotana" or Hopi or "Houma-Louisiana" or Huaco or Waco or Hualapai or Hupa or Natinixwe or "Natinook-wa" or "Hwech' in" or Hankutchin or "Iroquois Confederacy" or "Hodinoso ni" or "Illinois Confedera*" or Ilinoweg or Illini or Inupiat or Inuit or Ioway or Baxoje or Jicarilla or Juaneno or Acjachemen or Jumano or Kalapuya or Clackama or Kalispel or "Pend d' Oreilles" or Qlispe or Karuk or Karok or "Chum-ne" or Katkoc or Kansa or Kanza or Kawaiisu or Nuwa or Kennebec or "Kinipekw Kittitas" or Klickitat or "Qwu' lh-hwai-pum" or "Awi-adshi" or Mahane or Wahnookt or "Koa' aga' itoka" or Keresan or Kichai or Kitsai or Keechi or "K' itaish" or Kiowa or Gaigwu or Cauigu or Kutjau or "Kwu-da" or "Tep-da" or Kitanemuk or Kittitas or Klickitat or "Qwu' lh-hwai-pum" or "Awi-adshi" or Mahane or Wahnookt or "Koa' aga' itoka" or Konkow or "Koop Ticutta" or Koyukon or Ktunaxa or Kootenai or Flathead or Kucadikadi or "Kotsa' va" or Kumeyaay or "Tipai-Ipai" or Kamia or Diegueno or Kwapa or Cocopah or Cucapa or "Xawitt kwnchawaay" or Lassik or Lenape or "Leni-Lenape" or Lipan or Luiseno or Payomkawichum or Madqwadabaya or "Desert Yavapai" or Mahican or Mohicans or Makah or Makuhadokado or Maliseet or Wolistoqiag or Manahoac or Mahock or Meipontsky or Mandan or Mattole or "Bear River" or "Tul' bush" or "Ni' ekeni" or Meherrin or Menominee or Mackinac or Mescalero or Myaamiaki or Kickapoo or Twigtwee or Missouria or Miwok or Miwuk or Moadokado or Modoc or Mohave or "Aha Makhav" or Mohawk or "Kaneng' hega" or Molala or Molale or Molele or Nyyhmy or Moosonee or "Moose Cree" or Monsonis or Multnomah or Chinook or Nabedache or Nabaydacu or Wawadishe or Nabiltse or Dakubetede or "Nacho Nyak Dun" or Tutchone or Nacono or "Na' isha" or Nanticoke or Navajo or Ndee or Nial or Niimiipu or "Nez Perce" or Watapala or Watapahlute or Nisenan or Nisqually or Nomlaki or Noamlakee or "Central Wintun" or Nongatl or Nottoway or Cheroenhaka or "Northern Cheyenne" or Ohlone or Costanoan or Omaha or "O' odham" or Pima or Papago or Osage or Otoe or Otse or "Ozav Dika" or Palus or Passamaquoddy or Pestomuhkati or Patiri or Petaros or Pastia or Patwin or "Southern Wintun" or Panis or Skidi or Pedee or Penobscot or "Petun Piipaash" or "Kokmalik' op" or Piscatawa or Doeg or Conoy or "Pit River" or Pomo or Kashaya or Ponca or Ponka or Pottawatomi or Bodewadmik or Powhatan or Puyallup or Spuyalepabs or Quapaw or Ugahxpa or Quechan or Yuma or Kwtsaan or Quileute or Salinan or Saponi or Monacan or Sapon or "Eastern Blackfoot" or Christanna or Sawawatodo or Serrano or Taaqtam or "Maarenga' yam" or Yuhaviatam or Shasta or Chasta or Sasti or Shoshone or Siletz or Sinkine or Sinkyone or "Siuslaw Umpqua" or Skitswish or "Schitsu' umash" or Snohomish or Snuqualmi or Sokoki or Missiquoi or Stillaguamish or Stoluckwamish or Suquamish or Sutaio or Swinomish or Skagit or Syilx or Okanagan or Sotaae or "Taga Ticutta" or Takelma or Dagelma or Taltushtuntede or Galice or "Tanan Gwich' in" or Taos or Taovaya or Tataviam or Alliklik or Tawakoni or Tahuacano or Tenino or Thawikila or Hathawekela or "Fort Ancient" or Tigua or Tillamook or Nehalem or Timbisha or Panamint or Timpanogos or Tlingit or "Toi Ticutta" or Tolowa or "Talawa Dini' " or Tongva or Gabrieleno or Fernandeno or Tobikhar or Tonkawa or Ticanwatic or Tsikip or Appalousa or Opelousa or Tsitsistas or Tubatulabal or Tukabatchee or Tuscarora or Tomahittan or Kuskarawock or Tutelo or Tutero or Totteroy or Tutera or Yusan or Tututni or Umatilla or Umpqua or Waccamaw or Waxmaw or Wadatika or "Harney Valley Paiute" or Wailiki or Waluulapam or "Walla Walla" or Walpapi or Huipui or Wampanoag or Massasoit or Wanapum or Wappo or Washoe or Wichita or Willapa or Kwalhioqua or "Wi pukba" or "Verde Valley Yavapai" or Wintu or "Northern Wintun" or Wiyot or "Wee' at" or Weyet or Yakama or "Yamosopo Tuviwarai" or Yaqui or Yoeme or Yatasi or Yattasih or "Yavbe' " or "Yavapai" or "Ysleta del Sur" or Yojuane or Yokuts or Mariposa or Yuki or Yupighyt or "Yup'ik" or Yupik or Yurok or "Olekwo'l" or Zuni).mp. or ((Applegate or Delaware or Iowa or Ishak or Kaw or Kato or Spokane or Miami or Arkansas or Tali or Tunica or (Han not (China or Chinese)) or Pawnee or "Coeur D' Alene" or Piscataway or Ree or Tula) adj3 (reservation* or nation or people or peoples or population or man or men or woman or women or child* or youth* or elder or elders or communit* or tribe or tribes or tribal or Indian*)).mp. [United States]** | **28200** |
| **67** | **((Mexico/ or ((Mexico not "New Mexico") or Mexican or Aguascalientes or "Baja California" or Campeche or Chiapas or (Chihuahua not (dog or dogs or pet or pets)) or Coahuila or Colima or Durango or Guanajuato or Guerrero or Hidalgo or Jalisco or Michoacan or Morelos or Nayarit or "Nuevo Leon" or Oaxaca or Puebla or Queretaro or "Quintana Roo" or "San Luis Potosi" or Sinaloa or Sonora or (Tabasco not (sauce* or flavo*)) or Tamaulipas or Tlaxcala or Veracruz or Yucatan or Zatecas).mp.) and (exp Indigenous Peoples/ or (Mesoamerindian* or Indigen* or aborig* or "first people*" or "indos mexicano" or "original people*" or "pueblos indigenas").mp.)) or (Aguacatec or Akwa'ala or Abxubal or Ayuukja'ay or "Batzil k'op" or Binizaa or "Chichimeca Jonaz" or Chinantec or Chocho or "Ch ol" or Chontal or "Chuj" or Cochimi or Comcaac or Hamasipini or Harijio or "Ha shuta enima" or "Hach t'an" or Huastecor Hnahnu or Hnatho or Ixcatec or Ixil or Jacaltec or K'akchikel or K'anjobal or Kanjobal or Kaqchikel or Kechi or K'iche or Kikapooa or Kikapu or Kiliwa or "Ko'lew" or "K'op o winik atel" or Kumiai or Lacandon or Laymon or Makurawe or Maya or Maya'wiinik or Mazahua or Mazatec or Me'phaa or Mexicanero or Mexikatlajtolli or Mixe or Mixtec or Motocintleco or mti'pa or Nahuas or Ocuiltec or Otomi or (Oaxaca not Oaxaca-Blinder) or "Pame" or Papago or Tlahuica or Paipai or "Pima Bajo" or Purepecha or P'urhepecha or Qatok or (Quiche not Guatemala) or Q'iche or Raramuri or "Runixa ngiigua" or ("Seri" not "Seri 82") or "Slijuala sihanuk" or Tacuate or Tarahumara or Teenek or Tepehua or Ti'pai or Tlapanec or "Tohono O'odham" or Totonac or Tachiwin or "Tsa jujmi" or Tzotzil or "Tu'un savi" or Tzeltal or "Uza" or Winik or Xigue or Yucatec or Zapotec).ti,ab,kw. [Mexico]** | **4844** |
| **68** | **((exp India/ or Bangladesn/ or Bhutan/ or Nepal/ or Pakistan/ or Sri Lanka/ or (India or Bangladesh* or (Bhutan* not bhutanensis) or (Nepal* not nepalensis) or Pakistan* or "Sri Lanka*").mp.) and (Indigenous Peoples/ or ((Indigenous* or tribe or tribal or tribes or Rai) adj3 (population* or people* or person* or elder* or man or men or woman or women or child* or youth* or clan or clans or tribe or tribes or tribal or family or families or parent* or grandparent* or elder or elders or grandmother* or grandfather* or baby or babies or infant or infants or patient or patients or speakers or speaking or village* or communit*)).mp.)) or ("Adivasis" or "Adnamanese" or "Andaman" or "Baluch" or "Baluchis" or "Bodo" or "Boro" or "Boros" or "Bote" or "Brahuis" or "Chakmas" or "Chepang" or "Chhantyal" or "Damai" or "Dewan" or "Ghale" or "Gurkha" or "Gurung" or "Hayu" or "Hyolmo" or "Jarawa" or "Jirel" or "Jumma" or "Kalash" or "Khas" or "Kirati" or "Koinch" or "Kulung" or "Kusunda" or "Limbu" or "Lohorung" or "Magar" or "Makrani" or "Mangar" or "Marma" or "Miji" or "Mongar" or "Mro" or "Naga" or "Nepami" or "Newar" or "Nicobar" or "Onge" or "Rang" or "Raute" or "Sajolang" or "Santhal" or "Sentinelese" or "Sindhis" or "Sulemani" or "Sunuwar" or "Tamang" or "Thakali" or "Thangmi" or "Tharu" or "Tripura" or "Tumbahangphe" or "Wanniyala-Aetto" or "Yakkha" or "Yolmopa").mp. [Indian Sub-Continent]** | **7250** |
| **69** | **Indians, South American/ or (Abipon or Achuar or Achuagua or Akawaio or Amarizana or Andoque or Akawaio or Akuriyo or Anauya or Araona or Arawak or Ayamn or Aguaruna or Amahuaca or Amarakaeri or Andoa or Arabela or Arawak or Arhuaco or Ashaninca or Asheninca or Atsahuaca or Aymara or Ayoreo or Bakairi or "Baniva" or Barasana or Baniwa or "Baure" or Bororo or Cabiyari or Cacataibo or Caquinte or Cacua or Cahuarano or "Caiua" or "Camara Indians" or Camaracoto or Camsa or Canamari or Candoshi or Canela or Canichana or Capanahua or Carapana or Cariay or "Carib" or Carijona or Carutana or Cashibo or Cashinahua or Cawishana or Cavinena or Caxuiana or Cayuvava or Chontaquiro or Cocama or "Cubeo" or Curipaco or Chacobo or Chaima or (Chana not striatus) or Chapacura or Charrua or Chimila or Chitonahua or Chorote or Chipaya or Chiquitano or Chulupi or Carare or Coconuco or Cofan or Coreguaje or Coyaima or Chamacoco or Chamicuro or Chayahuita or Cocama or "Culina" or Culino or Cubeo or Cuiba or "Cuiva" or Cumanagoto or Curripaco or "Deni" or Desano or Embera or Guarani or Guajajara or "Guana" or Guanano or Guarayo or Guarayu or Guahibo or Guajiro or Guambiano or Guanano or Guayabero or Guarequena or Guinao or "Guana" or Gayon or Guahibo or Hixkaryana or Huachipairi or Huambisa or Huarayo or Lauanaua or Ikpeng or Ingariko or Irantxe or Itonama or Inapari or Iquito or Isconahua or Jumana or Japreria or Jirajara or Juruti or Jaqaru or Jebero or Kadiweu or Kaingang or Kamayura or Karaja or Karipuna or "Kariri" or Katukina or Kaxarari or Kayabi or Kayapo or "Kuikuro alapalo" or Kulina or "Kaiwa" or Kallawaya or "Kogui" or "Kuna" or Kaweskar or "Lule" or Macuna or Maipure or Mapuche or Mataco or Mocovi or Machinere or Machinerev or Machiguenga or Macushi or Macuna or "Madi" or Malayo or Mamainde or Manao or Mandauaca or Mandawaka or Mapidian or Mapuche or Mapidian or Maquiritare or Maquiritari or Maragua or Marawan or Mariate or Marubo or Mastanahua or Mataco or Matipuhy or "Matis" or "Matses" or Mawakua or Mawakwa or Maxakali or Mehinaku or Miranha or Moronawa or Munduruku or Movima or Muellama or Muinane or Mapoyo or "Mashco Piro" or Muniche or Nambikwara or Nocaman or Nuquini or Nomatsiguenga or Nanti or Ocaina or Omagua or Orejon or "Opon" or Pacahuara or "Paez" or Paicone or Palicur or Panare or "Pano" or "Paresi" or Paumari or "Pemon" or Pilaga or Puelche or Pauna or Pauserna or Piapoco or Piraha or Piratapuyo or Pisabo or Piaroa or "Pijao" or Piratapuyo or Paraujano or Pemon or Pemono or Piapoco or Puinave or Patamona or Poyanawa or Puinave or Puquina or Quechua or Quichua or Retuara or Resigaro or Reyesano or Sabanes or "Saliba" or Saluma or Sarave or Secoya or Selknam or "Sensi" or Shaninawa or Shapra or Sharanahua or Shebayo or Shiwiar or Shikiana or Sikiana or Siriono or Sinsiga or "Siona" or Suruwaha or Tacano or Tamanaco or Tiahuanaco or Tariano or Tehuelche or Tariano or Tatuyo or "Tembe" or "Terena" or Telembi or Ticuna or Ticuna or Tiriyo or Tiwanaku or Tiwanaku or "Torom" or "Totoro" or Tsimane or Tuberao or "Tucano" or Tunebo or Tuxinawa or Tuyuca or Uainuma or Urarina or Vilela or Waimaha or Waiampi or Waiwai or Wapishana or Waraiku or Warekena or Waura or Wayampi or Wayana or Wirina or Waimaha or Waunana or "Wiwa" or "Warao" or Wayuu or Witoto or Xavante or Xipaya or Xiriana or Xokleng or Yabaana or Yaminawa or Yaminahua or Yaruma or Yawalapiti or Yuracare or Yabarana or Yavitero or "Yine" or Yamana or Yaghan or Yucuna or Yurumangui or Yukpa or Yanesha or Yoranahua or Yagua or Yaminahua or Zaparo or Zamuco or "Trio Indians" or "More Indians" or "Bare Indians" or (("Inga" or "Maca" or "Leco" or "Mojo" or "Uro" or "Maco" or Lengua or "Toba" or "Zoe" or "Ona" or "Catio" or "Passe" or "Bari" or "Awa" or "Bora" or "Bara" or "Remo" or "Pano" or "Sape") adj3 (Indians or Indian or Indigenous or Amerindian* or Aborigin* or people or peoples or elder or elders or grandmother* or grandfather* or parent* or women or men or woman or man or child* or youth or youths or baby or babies or tribe or tribes or tribal or shaman* or native or patient or patients))).mp. or ((Indian* or Amerindian, or Aboriginal* or Indigenas or Indigenous or tribe or tribes or tribal).mp. and (exp South America/ or "South America*".mp. or Argentin*.mp. or Bolivia*.mp. or Brazil*.mp. or Chile*.mp. or Colombia*.mp. or Ecuador*.mp. or "French Guiana".mp. or Guyana*.mp. or Paraguay*.mp. or Peru.mp. or Peruvian.mp. or Suriname.mp. or Uruguay*.mp. or Venezuela*.mp. or "Amazon Region".mp. or Amazonia.mp. or Andes.mp. or Andean.mp.)) [South America]** | **12538** |
| **70** | **((Ainus or Ainu or Aleuts or Alyutors or Chukchis or Chuvans or Dolgans or Enets or Entsy or Yupik or Yup'ik or Yuit or Yupigyt or Chaplino or Naukan or Itelmens or Kamchadals or Kereks or "Komi" or Koryaks or Nenets or Nentsy or Nganasans or Tavgi or Sami or Veps or Yukaghirs or Chulyms or Evenks or Tungus or Evens or "Kets" or Khantys or Mansi or Vguls or Selkups or Teleuts or Nanais or Nanaitsy or Negidal or Nivikh or Oroch or orok or Taz or udege or ulch or Kumadins or Chelkans or Shorians or Soyots or Telengits or Tofalars or Tugalars or "Tufans Todzhins" or Laks or Tabasarans or Turuls or Aguls or Tsakhurs or Kumyks or Nogais or "Andis" or Akhvakh or Archins or Bagvalals or Bezhta or Botlikhs or Chamalals or Godoberi or Hinukh or Hunzibs or Khwarshi or Karata or Tindis or Tsez or Abazin or Besermyan or Izhorians or Karelians or Nagaybaks or Setos or Shapsugs or Quratay).mp. or (exp Indigenous people/ or (Indigenous adj3 (population* or people* or person* or elder* or man or men or woman or women or child* or youth* or clan or clans or tribe or tribes or tribal or family or families or parent* or grandparent* or elder or elders or grandmother* or grandfather* or baby or babies or infant or infants or patient or patients or speakers or speaking or village* or communit*)).mp.)) and (Russia*.mp. or exp Russia/)** | **1056** |
| **71** | **((Greenland/ or Greenland*.mp. or "Kalaallit Nunaat".mp. or Nuuk.mp. or Sisimiut.mp. or Ilulissat.mp. or Qaqortoq Aasiaat.mp. or Maniitsoq.mp. or Tasiilaq.mp. or Uummannaq.mp. or Narsaq.mp. or Paamiut.mp. or Nanortalik.mp. or Upernavik.mp. or Qasigiannguit.mp.) and (Indigenous Peoples/ or Inuit.mp. or Inuk.mp. or ((Indigen* or Aborigin*) adj3 (population* or people or peoples or person or persons or elder or elders or man or men or woman or women or child* or youth* or clan or clans or tribe or tribes or tribal or family or families or parent* or grandparent* or elder or elders or grandmother* or grandfather* or baby or babies or infant or infants or patient or patients or speakers or speaking or village* or communit*)).mp.)) or (Greenlandic or Kalaallit or Kalaallisut or Tunumiit or Inughuit or Avanersuarmiut).mp. [Greenland ]** | **1276** |
| **72** | **((exp Asia, Southeastern/ or ("Bali" or "Borneo" or "Brunei" or Cambodia or "Flores Island" or "Indonesia" or Java or Komodo or "Laos" or "Lombok" or "Malasia" or "Malay Peninsula" or "Myanmar" or "Mekong Valley" or "New Guinea" or "Philippeans" or "Sabah" or "Singapore" or "Sumatra" or "Sumba" or "Sumbawa" or "Thailand" or "Timor" or "Vietnam").mp.) and (Indigenous Peoples/ or ((Indigen* or aborig*) adj3 (population* or people or peoples or person or persons or elder or elders or man or men or woman or women or child* or youth* or clan or clans or tribe or tribes or tribal or family or families or parent* or grandparent* or elder or elders or grandmother* or grandfather* or baby or babies or infant or infants or patient or patients or speakers or speaking or village* or communit*)).mp.)) or ("Abui" or "Aeta" or "Alfur" or "Ati People" or "Bahau" or "Bali Aga" or "Bali Mula" or "Baliaga" or "Balinese" or "Basap" or "Bataq" or "Batek" or "Bateq" or "Batin" or "Blaan" or "Brao" or "Bru" or "Bru-Van Kieu" or "Bugkalot" or "Bumiputera" or "Bunong" or "Chams" or "Champa" or "Cheq Wong" or "Dasun" or "Dayak" or "Deli Malay" or "Filipina*" or "Filiopino*" or "Gaddang" or "Ibaloi" or "Iban" or "Idan" or "Igorot" or "Jahai" or "Jehai" or "Jakun" or "Javanese Kshatriya" or "Kangeanese" or " Kagayanen" or "Kanayatn" or "Katu" or "Kayan" or "Kavet" or "Kenyah" or "Khmer Krom" or "Kimaragang" or "Klemantan" or "Kreung" or "Kuijau" or "Kuy" or "Kwijau" or "Lampung*" or "Lanoh" or "Lawangan" or "Luangan" or "Lumad" or "Dayak" or "Loloan Malay*" or "Mah Meri" or "Malay Singaporean*" or "Malbog" or "Maluku" or "Mangka'ak" or "Mangyan" or "Maniq" or "Maragang" or "Ma'anyan" or "Minokok" or "Moluccas" or Montagnard or "Murut" or "Naga" or "Negrito" or "New Guinea" or "Ngaju" or "Orang Hulu" or "Orang Kanaq" or "Orang Kuala" or "Orang Seletar" or "Orang Ulu" or "Ot Danum" or "Palawano" or "Palembang*" or " Panay-Bukidnon" or "Pan-ayanon" or "Papua" or "Phnong" or "Punong" or "Pear People" or "Por People" or "Punan" or "Rumanau" or "Sasak" or "Semai" or "Semang" or "Semaq Beri" or "Semelai" or "Senoi" or "Simeulue" or "Suludnon" or "Sunda Island*" or "Tabanwa" or "Takua" or "Tambanuo" or " Tombonuo" or "Taron" or "Temiar*" or "Temoq" or "Temuan" or "Tidung" or "Trone" or "Taaw't Bato").mp. [Southeast Asia]** | **11954** |
| **73** | **(Indigenous Peoples/ or ((Indigen* or aborig*) adj3 (population* or people or peoples or person or persons or elder or elders or man or men or woman or women or child* or youth* or clan or clans or tribe or tribes or tribal or family or families or parent* or grandparent* or elder or elders or grandmother* or grandfather* or baby or babies or infant or infants or patient or patients or speakers or speaking or village* or communit*)).mp.) and (pacific islands/ or exp melanesia/ or exp micronesia/ or exp polynesia/ or (Micronesia or "Caroline Island" or Guam or Kiribati or "Mariana Island*" or Nauru or Palau or Melanesia or "East Nusa Tenggara" or Fiji or Fijian or "Loyalty Island*" or "Maluku Islands" or Moluccas or "New Caledonia*" or "New Guinea" or Papua or Rotuma or "Solomon Island*" or Vanuatu or Polynesia or "Austral Island*" or "Cook Island*" or "Chatham Islands" or "Easter Island*" or Hawaii* or "Marquesas Island" or "Norfolk Island" or Samoa or Tahiti or Tokelau or Tonga or Tuvalu or Tuamotu or Kermadec or Banabans or "Native Hawaiian*" or Guamanian or Chamarro* or Chuukese or I-Kiribati or Kosraean* or Maohi or Marshallese or Moriori or Mortlockese or Mwokilese or Namonuito or Nauruans or Ngatikese or Niuean or Paafang or Paluans or Pingelapese or Pohnpeian* or Pollapese or Puluwat or Rapanui or Rotuman or Satwalese or Samoan*or Sonsorolese or Tahitian* or Tobian or Tongan or "Torres Strait Islander*" or Tuvaluan or Ulithian or Woleian or Yapese or Polynesian* or Micronesian* or Melanesian*).mp.) [Pacific Islands]** | **6104** |
| **74** | **((exp Indigenous Peoples/ or ((Indige* or aborig* or nomadic or seminomad* or "semi nomad*") adj3 ("people" or "peoples" or "elder" or "elders" or "grandmother*" or "grandfather*" or "parent*" or "women" or "men" or "woman" or "man" or "child*" or "youth" or "youths" or "baby" or "babies" or "tribe" or "tribes" or "tribal" or "shaman*" or "native" or "patient*")).mp.) and (("Middle East" or "Afghanistan" or "Bahrain" or "Iran" or "Iraq" or "Israel" or ("Jordan" not "Jordan's Principle") or "Kuwait" or "Lebanon" or "Oman" or "Palestine" or "Qatar" or "Saudi Arabia").mp. or middle east/ or afghanistan/ or bahrain/ or iran/ or iraq/ or israel/ or jordan/ or kuwait/ or lebanon/ or oman/ or qatar/ or saudi arabia/ or syria/ or turkey/ or united arab emirates/ or yemen/)) or ("Bedouin*" or "Jahalin" or "al-Kaabneh" or "al-Azazmeh" or "al-Ramadin" or "al-Rshaida").mp. [Middle East]** | **1363** |
| **78** | **or/138-152** | **106434** |
| **79** | **137 and 153** | **352** |
| **75** | **(2023062* or 202307*or 202308* or 202309* or 20231* or 2024*).dt,ez,da.** | **900350** |
| **76** | **154 and 155** | **15** |

**Embase <1974 to 2024 February 13>**

| # | Search Statement | Results |
| --- | --- | --- |
| 1 | urinary incontinence/ or urinary incontinence, stress/ or urinary incontinence, urge/ | 51655 |
| 2 | exp Fecal Incontinence/ | 24906 |
| 3 | pelvic organ prolapse/ or cystocele/ or rectal prolapse/ or uterine prolapse/ or visceral prolapse/ | 26541 |
| 4 | Urinary Bladder, Overactive/ | 8227 |
| 5 | exp Urinary Retention/ | 36018 |
| 6 | Constipation/ | 113440 |
| 7 | Encopresis/ | 19523 |
| 8 | Pelvic Floor Disorders/ | 2743 |
| 9 | Pelvic Girdle Pain/ | 557 |
| 10 | exp Dyspareunia/ | 13098 |
| 11 | exp Vulvodynia/ | 1991 |
| 12 | exp Vaginismus/ | 1262 |
| 13 | exp Cystitis, Interstitial/ | 6636 |
| 14 | Adenomyosis/ | 7022 |
| 15 | Pudendal Neuralgia/ | 358 |
| 16 | exp Gynecologic Surgical Procedures/ | 188739 |
| 17 | exp Colectomy/ | 21010 |
| 18 | exp Rectal Neoplasms/ | 79108 |
| 19 | exp Endometrial Neoplasms/ or exp Uterine Neoplasms/ or exp Genital Neoplasms, Female/ or exp Ovarian Neoplasms/ | 429133 |
| 20 | exp Prostatic Neoplasms/ | 302973 |
| 21 | exp Prostatectomy/ or exp Prostatic Neoplasms/ | 325647 |
| 22 | ((gyn?eologic* or rectal or rectum or genital or prostat* or unterine or uterus or ovarian or endometrial) adj3 (surgery or surgical)).mp. [mp=title, abstract, heading word, drug trade name, original title, device manufacturer, drug manufacturer, device trade name, keyword heading word, floating subheading word, candidate term word] | 43728 |
| 23 | 16 or 17 or 18 or 19 or 20 or 21 or 22 | 964544 |
| 24 | exp Postoperative Complications/ or (postoperative or "post operative" or "post surg*").mp. | 1620455 |
| 25 | 23 and 24 | 111079 |
| 26 | exp Urinary Bladder, Neurogenic/ | 11916 |
| 27 | sexual dysfunction, physiological/ or erectile dysfunction/ or impotence, vasculogenic/ | 53899 |
| 28 | exp Nocturnal Enuresis/ | 4595 |
| 29 | Encopresis/ | 19523 |
| 30 | exp Hirschsprung Disease/ | 8557 |
| 31 | Sex Reassignment Surgery/ | 210 |
| 32 | Lichen Sclerosus et Atrophicus/ | 5229 |
| 33 | exp Nocturia/ | 9884 |
| 34 | urinary fistula/ or urinary bladder fistula/ or vaginal fistula/ or rectovaginal fistula/ or vesicovaginal fistula/ | 13175 |
| 35 | Pelvic Floor/ | 508 |
| 36 | lower urinary tract symptoms/ or dysuria/ or nocturia/ or prostatism/ or urinary bladder, overactive/ or urinary bladder, underactive/ or urinary incontinence/ or urinary incontinence, stress/ or urinary incontinence, urge/ | 89168 |
| 37 | 1 or 2 or 3 or 4 or 5 or 6 or 7 or 8 or 9 or 10 or 11 or 12 or 13 or 14 or 15 or 25 or 26 or 27 or 28 or 29 or 30 or 31 or 32 or 33 or 34 or 35 or 36 | 451460 |
| 38 | (prolaps* adj3 ("pelvic organ" or uterine or uterus or bladder or rectal or rectum or urethra* or apical)).mp. | 32353 |
| 39 | [incontinence.mp](http://incontinence.mp/). | 130543 |
| 40 | (uterocele or cystocele or rectocele or enterocele or urethrocele or sigmoidocele).mp. [mp=title, abstract, heading word, drug trade name, original title, device manufacturer, drug manufacturer, device trade name, keyword heading word, floating subheading word, candidate term word] | 7150 |
| 41 | (bladder* adj3 (overactive or urgency or urgent or frequency or heistency or retention)).mp. [mp=title, abstract, heading word, drug trade name, original title, device manufacturer, drug manufacturer, device trade name, keyword heading word, floating subheading word, candidate term word] | 24165 |
| 42 | (bowel adj3 (frequency or urgency)).mp. [mp=title, abstract, heading word, drug trade name, original title, device manufacturer, drug manufacturer, device trade name, keyword heading word, floating subheading word, candidate term word] | 2697 |
| 43 | [constipation.mp](http://constipation.mp/). | 125364 |
| 44 | [encopresis.mp](http://encopresis.mp/). | 991 |
| 45 | (dyssynergia adj3 (bowel* or bladder*)).mp. | 197 |
| 46 | "vaginal wind".mp. | 16 |
| 47 | "pelvic girdle pain".mp. | 754 |
| 48 | (dyspareunia or vulvodynia or vaginismus or vestibulodynia or "interstitial cystitis" or "painful bladder syndrome" or "proctalgia fugax" or prostatitis or endometriosis or adenomyosis or "pudendal neuralgia" or anismus).mp. [mp=title, abstract, heading word, drug trade name, original title, device manufacturer, drug manufacturer, device trade name, keyword heading word, floating subheading word, candidate term word] | 91815 |
| 49 | Neurogenic [bladder.mp](http://bladder.mp/). | 13630 |
| 50 | ("sexual dysfunction" or "erectile dysfunction" or "persistent genital arousal" or impostence).mp. [mp=title, abstract, heading word, drug trade name, original title, device manufacturer, drug manufacturer, device trade name, keyword heading word, floating subheading word, candidate term word] | 80429 |
| 51 | ("bed wett*" or bedwett* or encopresis or "hirshprung* disease").mp. [mp=title, abstract, heading word, drug trade name, original title, device manufacturer, drug manufacturer, device trade name, keyword heading word, floating subheading word, candidate term word] | 1916 |
| 52 | ("gender confirmation surg*" or "sex reassignment surg*").mp. [mp=title, abstract, heading word, drug trade name, original title, device manufacturer, drug manufacturer, device trade name, keyword heading word, floating subheading word, candidate term word] | 818 |
| 53 | "lichen sclerosis".mp. | 440 |
| 54 | [nocturia.mp](http://nocturia.mp/). | 11294 |
| 55 | ((fistula* or fistulae) adj3 (bladder* or urinary or vagina* or rectovaginal* or vesicovaginal*)).mp. [mp=title, abstract, heading word, drug trade name, original title, device manufacturer, drug manufacturer, device trade name, keyword heading word, floating subheading word, candidate term word] | 13006 |
| 56 | "levator ani avulsion".mp. | 110 |
| 57 | ("Lower urinary tract symptom*" or dysuria or "LUTS").mp. [mp=title, abstract, heading word, drug trade name, original title, device manufacturer, drug manufacturer, device trade name, keyword heading word, floating subheading word, candidate term word] | 45304 |
| 58 | "voiding dysfunction".mp. | 5078 |
| 59 | or/37-58 | 597140 |
| 60 | ((exp Indians, North American/ and Canad*.mp.) or Indigenous Canadians/ or exp Inuits/ or exp Health Services, Indigenous/ or exp Ethnopharmacology/ or (Athapaskan or Saulteaux or Wakashan or Cree or Dene or Inuit or Inuk or Inuvialuit* or Haida or Ktunaxa or Tsimshian or Gitxsan or Gitksan or "Nisga'a" or Haisla or Heiltsuk or Oweenkeno or "Kwakwaka'wakw" or "Nuu chah nulth" or "Tsilhqot'in" or Dakelh or "Wet'suwet'en" or Sekani or Dunne-za or Dene or Tahltan or Kaska or Tagish or Tutchone or Nuxalk or Salish or St'at'imc or Stl'atl'imx or Stl'atl'imc or Nlaka'pamux or Okanagan or "Sec wepmc" or Secwepemc or Tlingit or Anishinaabe or Blackfoot or Nakoda or Tasttine or "Tsuu T'ina" or "Tsuut'ina" or "Gwich'in" or (Han not (China or Chinese)) or Algonquin or Nipissing or Ojibwa or Potawatomi or Innu or Maliseet or "Mi'kmaq" or Micmac or Passamaquoddy or Haudenosaunee or Cayuga or Mohawk or Oneida or Onondaga or Seneca or Tuscarora or Wyandot or Aboriginal* or Indigenous* or Metis or red road or "on reserve" or off-reserve or First Nation or First Nations or Amerindian).mp. or (urban adj3 (Indian* or Native* or Aboriginal*)).mp. or [ethnomedicine.mp](http://ethnomedicine.mp/). or country food*.mp. or residential school*.mp. or ((exp Medicine, Traditional/ or traditional medicine*.mp.) not Chinese.mp.) or exp Shamanism/ or shaman*.mp. or traditional heal*.mp. or traditional food*.mp. or medicine [man.mp](http://man.mp/). or medicine [woman.mp](http://woman.mp/). or autochtone*.mp. or (Native* adj1 (man or men or women or woman or boy* or girl* or adolescent* or youth or youths or person* or adult or people* or Indian* or Nation or tribe* or tribal or band or bands)).mp.) and (exp Canada/ or (Canad* or British Columbia or Colombie Britannique or Alberta or Saskatchewan or Manitoba or Ontario or Quebec or Nova Scotia or New Brunswick or Newfoundland or Labrador or Prince Edward Island or Yukon Territory or NWT or Northwest Territories or Nunavut or Nunavik or Nunatsiavut or NunatuKavut).mp.) [Canada] | 10538 |
| 61 | ((exp australia/ or (australia* or northern territory or tasmania or new south wales or Victoria or queensland).ti,ab.) and ("Native Hawaiian or Other Pacific Islander"/ or aborigin*.ti,ab. or indigenous.ti,ab.)) or "torres strait islander*".ti,ab. [Australia and Torres Strait] | 14442 |
| 62 | (Maori or "tangata whenua").mp. or ((exp New Zealand/ or (New Zealand or Aukland).mp.) and ("Native Hawaiian or Other Pacific Islander"/ or (Aborig* or Indig*).ti,ab.)) [Maori] | 7485 |
| 63 | (Saami or Sampi or (Sami not Ulus) or Samis or Southernsami* or Umesami* or Pitesami* or Lulesami* or Northernsami* or Enaresami* or Kolasami* or Lapp or Lapps or Lappish or Lappland or (Lapland* not longspur) or Lappalainen* or Saamelainen* or reindeer herd* or reindeer culture* or reindeer pastoral* or Lappbys or Samebys or reinbeitesdistrikt or paliskunta or siida).mp. or (((Fennoscandia or Finnmark or Scandinavia or Nordic or Sweden or Norway or Finland or Swedish or Finnish or Norwegian or Norge or Svensk* or Suomi or Barents Region or (Kola not (garcinia or gotu)) or Arctic Europe* or Polar Europe* or North* Europ*).mp. or Finland/ or Norway/ or Sweden/) and ((traditional adj3 (food* or heal* or medicine* or shaman*)) or (Indigenous* adj3 (people* or person* or mother* or father* or parent* or grandparent* or grandmother* or grandfather* or elder or elders or child* or boy or boys or girl* or youth* or healer* or patient or patients or famil* or herder* or village* or communit*))).ti,ab.) | 2053 |
| 64 | ((exp indigenous people/ or "indigenous people* ".mp.) and (exp Africa/ or ((Africa* not "African American*") or Algeria or Angola or Benin or Botswana or "Burkina Faso" or Burundi or Cameroon or "Cape Verde" or "Cabo Verde" or "Central African Republic" or Chad or Comoros or Congo or Djibouti or Egypt or "Equitorial Guinea" or Eritrea or Eswatini or Ethiopia or Gabon or Gambia or Ghana* or Guinea or "Guinea Bissau" or "Ivory Coast" or Kenya or Lesotho or Liberia or Libya or Madagascar or Malawi or Mali or Mauritania or Mauritius or Mayotte or Morocco or Mozambique or Namibia or Niger or Nigeria or Reunion or Rwanda or "Saint Helena" or Ascension or "Tristan de Cunha" or "Sao Tome" or Principe or Senegal or Seychelles or "Sierra Leone" or Somalia or "South Africa" or Sudan or Tanzania or Togo or Tunisia or Uganda or "Western Sahara" or Zambia or Zimbabwe).mp.)) or (Abakuria or Abaluhya or Abagusii or Abakuria or Aembu or Agikuyu or Akamba or Anuak or Anywaa or Amazigh or Ambala or Ambeere or Ambundu or Ambuun or Amharan or Angba or Baaka or Baamba or Babindi or Babini or Baboma or Bachokwe or Bacwa or Bafumbira or Baganda or Bagyele or Bagwere or Bagyeli or Bakiga or Bakola or Baholo or Bakalanga or Bakiga or Bakolo or Bakongo or Bakonjo or Baluba or Balunda or Balovale or Bamasaba or Bambuti or Bangala or Bangoli or Bangungu or Bantu or Banyankole or Banyarwanda or Banyole or Banyoro or Bapende or Bapedi or Barabaig or Barombi or Barundi or Baruuli or Basamia or Basoga or Batswana or Batooro or Batsamba or Batswana or Batwa or Bayaka or Bedzan or Bazombe or Bebayaka or Bedzan or Bhaca or Biaka or Borana or Chewa or Copts or Cormorian or Cushitic or Dahalo or Datooga or Dikidiki or Dogon or Ewondo or Fulani or Fuliru or Ganguela or Gciriku or Gyele or Hadza or Hadzabe or Haillom or Haratin or Herero or Himba or Hlubi or Iriryen or Iqvayliyen or Kabyle* or Kalenjin or Kanioka or Kanioka or Kaonde or Karamojong or Kavango or Kereuyu or Khoikhoi or KhoiSan or Kikuyu Kwangali or Lokele or Lowme or Lotuko or Lwalwa or Maasai or Makonde or Makua or Mande or Masalit or Matumbi or Mayeuyi or Mayeyi or Mbenga or Mbukushu or Mbochi or Mboro or Mbuti or Medzan or Mijikenda or Mozabite* or Nafusa or Ndebele or Ngombe or Namaqua or Nyanga or Nyamwezi or Ogiek or Ovambo or Ovimbundu or Phuthi or Pokomo or Rendille or Riffian or Riffians or Sakuma or Samburu or Sandawe or Sangha or Sango or Sengwer or Serer or Sesotho or Shangaan or Shawiya or Shenwa or Shi or Shilluk or Sukua or Sukus or Swahili or Tabwa or Tambuka or Taveta or Thembu or Tigrayan or Topoke or Tsonga or Toubou or Tuareg or Tumbuka or Ugana or Wochua or Xhosa or Xindonga or Yoruba or Zenati or Zuwara or ((Indigenous or Afar or Afars or "Aka People" or Akie or Ameru or Asua or Ateker or Atwot or Awjila or Bafia or Baka or Bakongo or Bakwe or Balunda or Balovale or Bango or Bassa or Beja or Bekpak or Bemba or Bembe or Benet or Berber or Berbers or Bira or Bowe or Bubi or Budja or Bulu or Bunrun or Chaga or Chopi or Damara or Dinka or Djerba or Duala or Dzing or Efe or Elmolo or Fang or Foora or Fula or Fur or Ghomara or Ghadames or Gllana or Glu or Gogo or Gongo or Haya or Havu or Hema or Hima or Hunde or Hutu or Huva or Iboko or Igbo or Ijo or Jieng or Kadu or Kande or Kango or Katla or Kgaga or Khoe or Kola or Komo or Kota or Kua or Kuba or Kwango or Kx'z or Kxoe or Lala or Lozi or Luo or Luba or Lupu or Masmuda or Matmata or Mbala or Mbam or "Mbo People" or Mbolo or Mbuza or Mongo or Mpondo or Myene or Naadh or Nama or Nande or Naro or Ngoni or Ndau or Ndebele or Ngoli or Ngondi or Ngoni or Nguni or Nkoya or Nkumu or Nuba or Nubian or Nuer or Nzebi or Ogoni or Omoro or Oroko or Pygmy or Popoi or Poto or Puru or Rashad or (San not ("San Francisco" or "San Diego" or "San Antonio")) or Sango or Sanhaja or Sena or Shilha or Shira or Shona or Shua or Sokna or Somali* or Sotho or Sua or Subu or Swazi or Taitaa or Tchokwe or Teke or Tembo or Tetela or (Tonga and Africa*) or Tshwa or Tsoa or Twa or Turkana or Tuu or Venda or Vira or Watta or Wakuti or Yaaku or Yaka or Yakoma or Yanzi or Yao or Yeke or Yela or Yeyi or Zulu) adj3 (population* or people* or person* or elder* or man or men or woman or women or child* or youth* or clan or clans or tribe or tribes or tribal or family or families or parent* or grandparent* or elder or elders or grandmother* or grandfather* or baby or babies or infant or infants or patient or patients or speakers or speaking or village* or communit*))).ti,ab. [Africa] | 38363 |
| 65 | (((Acatec or Aguacateco or Amuzgo or Bokota or Boruca or Bribri or "Bri Bri" or Buglere or Cabecar or Cakchiquel or Changuena or Chatino or Chiapanec or Chicomuceltec or Chinantee or Chocho or Cholti or "Ch'olti'" or "Ch'olti'anor Chontal" or Chorotega or Chorti or Chuj or Chumbia or Corobici or (Cueva not Spain) or Cuicatec or Cuitlatee or Cuytec or Dorasque or Embera or Garifuna or Guatuso or Guaymi or Guaymis or Guetar or Huastec or Huave or Huetar or Itzaj or Ixil or Jacalteco or Jonaz or Kanjobal or Kekchi or Kuna or Maleku or Mangue or Matambu or Matlatzinca or Mazahua or Motozintlec or Mayan or Mayangna or Miskito or Mixtec or Mopan or Nahua or Nahuatl or Ngabe or Otomi or Pantec or Paya or Popoloca or Popoloc or Poqomam or Poqomchi or "Q'eqchi'" or Quiche or Quitirrisi or Sacapulteco or Sipacapense or Subtiaba or Tacaneco or Tarasco or Tamaulipec or Tamazultec or Tecoxquin or Tectiteco or Tecual or Tecuexe or Tepehura or Tepuztecor or Teribe or Terraba or Totonac or Trique or Tzeltal or Tzotzil or Tzutujil or Ulwa or Uspantec* or Uspanteko or Voto or Xinca or Waunana or Wounaan or Yucatec or Zapotec or Zoque or (("Costa Rica*" or Hondura* or Nicaragua* or Panama* or Guatemala* or Achi or Belize or Belizean* or Maya* or Mixe or Pame or Pipil or Pech or Chol or "Ch'olan" or Cora or Cuna or Mam or Rama) adj5 (Indian or Indians or Amerindian* or Amerindio* or Aboriginal* or Indigenous or Indigena* or Aborigen* or Mestizo or tribe or tribes or tribal or "traditional medicine*" or shaman*))).tw. or (Belize or "Costa Rica" or "El Salvador" or Guatemala or Honduras or Nicaragua or Panama).ti,ab.) and (exp indigenous people/ or (indigenous adj3 (population* or people* or person* or elder* or man or men or woman or women or child* or youth* or clan or clans or tribe or tribes or tribal or family or families or parent* or grandparent* or elder or elders or grandmother* or grandfather* or baby or babies or infant or infants or patient or patients or speakers or speaking or village* or communit*)).mp.)) not ((Mexico or Mexican).ti,ab. or exp Mexico/) [Central America] | 619 |
| 66 | Indigenous People/ or American Indian/ or Canadian Aboriginal/ or Eskimo/ or Inuit/ or Indigenous Health Services/ or ("A' ani" or Absaroka or Haaninin or Atsina or "Gros Ventre" or Acopsel or Tlacopsel or Lacopsel or Ahtna or Ahtena or Akenitsi or Occaneechi or Akokisa or Horcoquisa or Orcoquizas or Aleut or Unangax or Unangan or Alibamu or "Alabama Alsea" or Alutiiq or Sugpiag or Amahami or Awaxawi or Androscoggin or Arosaguntacook or Ameriscoggin or Anishinaabeg or Chippewa or Anihsinape or Saulteaux or Apalachee or Aranama or "Texan Coahuilteca" or Tamique or Arikara or Sahnish or Arickaree or Adakadaho or Assiniboine or Hohe or Nakota or Nakoda or Nakona or "Atsa' Kudok-wa" or Awatixa or Bannock or "Snake Indian*" or Bidai or Quasmigdo or Biloxi or Blackfoot or Niitsitapi or Sikasikaitsitapi or Cahto or Kaipomo or Cahuilla or Ivilyuqaletem or Ivilyuat or Catawba or Inna or Iswa or Chemehuevi or Chickasaw or "Chilula Chimakum" or Aqokulo or Chimariko or Chiricahua or Tsokanende or Chitimacha or Chetimachan or Sitimacha or Chowanoke or Roanoke or Chumash or Ciboney or "Taino Ciwat" or Clatsop or Coos or Coosa or Uchis or Chiaha or Coste or Talisi or Coquille or Kokwell or Coso or Cowlitz or Taitnapam or "Crow Nation" or "Cui Ui Ticutta" or Cupeno or Kuupangaxwichem or Cupa or "Cup' ig" or Nunivak or "Dakota Oyate" or Lakota or Nakota or Santee or Teton or Sioux or Deadose or "Deg Xina" or "Deg Xit' an" or Kaiyuhkhotana or "Deg Hit' an" or "Dena' ina" or Tanaina or "Dichinanek' Hwt' ana" or "Upper Kuskokwim Athabascan*" or Kolchan or Goltsan or "Tundra Kolosh" or "Do lkabya" or Duwamish or Esselen or Eyak or "Gidi' tikadi" or Guwevkabaya or "Gwich' in" or Kutchin or Haida or Xaadas or Xaat or Halchidhoma or Havasupai or "Green Water People" or Hiratsa or Hiraaca or "Ho-chaaqa" or Winnebago or Holikachuk or Innoko or "Tlegon-khotana" or Hopi or "Houma-Louisiana" or Huaco or Waco or Hualapai or Hupa or Natinixwe or "Natinook-wa" or "Hwech' in" or Hankutchin or "Iroquois Confederacy" or "Hodinoso ni" or "Illinois Confedera*" or Ilinoweg or Illini or Inupiat or Inuit or Ioway or Baxoje or Jicarilla or Juaneno or Acjachemen or Jumano or Kalapuya or Clackama or Kalispel or "Pend d' Oreilles" or Qlispe or Karuk or Karok or "Chum-ne" or Katkoc or Kansa or Kanza or Kawaiisu or Nuwa or Kennebec or "Kinipekw Kittitas" or Klickitat or "Qwu' lh-hwai-pum" or "Awi-adshi" or Mahane or Wahnookt or "Koa' aga' itoka" or Keresan or Kichai or Kitsai or Keechi or "K' itaish" or Kiowa or Gaigwu or Cauigu or Kutjau or "Kwu-da" or "Tep-da" or Kitanemuk or Kittitas or Klickitat or "Qwu' lh-hwai-pum" or "Awi-adshi" or Mahane or Wahnookt or "Koa' aga' itoka" or Konkow or "Koop Ticutta" or Koyukon or Ktunaxa or Kootenai or Flathead or Kucadikadi or "Kotsa' va" or Kumeyaay or "Tipai-Ipai" or Kamia or Diegueno or Kwapa or Cocopah or Cucapa or "Xawitt kwnchawaay" or Lassik or Lenape or "Leni-Lenape" or Lipan or Luiseno or Payomkawichum or Madqwadabaya or "Desert Yavapai" or Mahican or Mohicans or Makah or Makuhadokado or Maliseet or Wolistoqiag or Manahoac or Mahock or Meipontsky or Mandan or Mattole or "Bear River" or "Tul' bush" or "Ni' ekeni" or Meherrin or Menominee or Mackinac or Mescalero or Myaamiaki or Kickapoo or Twigtwee or Missouria or Miwok or Miwuk or Moadokado or Modoc or Mohave or "Aha Makhav" or Mohawk or "Kaneng' hega" or Molala or Molale or Molele or Nyyhmy or Moosonee or "Moose Cree" or Monsonis or Multnomah or Chinook or Nabedache or Nabaydacu or Wawadishe or Nabiltse or Dakubetede or "Nacho Nyak Dun" or Tutchone or Nacono or "Na' isha" or Nanticoke or Navajo or Ndee or Nial or Niimiipu or "Nez Perce" or Watapala or Watapahlute or Nisenan or Nisqually or Nomlaki or Noamlakee or "Central Wintun" or Nongatl or Nottoway or Cheroenhaka or "Northern Cheyenne" or Ohlone or Costanoan or Omaha or "O' odham" or Pima or Papago or Osage or Otoe or Otse or "Ozav Dika" or Palus or Passamaquoddy or Pestomuhkati or Patiri or Petaros or Pastia or Patwin or "Southern Wintun" or Panis or Skidi or Pedee or Penobscot or "Petun Piipaash" or "Kokmalik' op" or Piscatawa or Doeg or Conoy or "Pit River" or Pomo or Kashaya or Ponca or Ponka or Pottawatomi or Bodewadmik or Powhatan or Puyallup or Spuyalepabs or Quapaw or Ugahxpa or Quechan or Yuma or Kwtsaan or Quileute or Salinan or Saponi or Monacan or Sapon or "Eastern Blackfoot" or Christanna or Sawawatodo or Serrano or Taaqtam or "Maarenga' yam" or Yuhaviatam or Shasta or Chasta or Sasti or Shoshone or Siletz or Sinkine or Sinkyone or "Siuslaw Umpqua" or Skitswish or "Schitsu' umash" or Snohomish or Snuqualmi or Sokoki or Missiquoi or Stillaguamish or Stoluckwamish or Suquamish or Sutaio or Swinomish or Skagit or Syilx or Okanagan or Sotaae or "Taga Ticutta" or Takelma or Dagelma or Taltushtuntede or Galice or "Tanan Gwich' in" or Taos or Taovaya or Tataviam or Alliklik or Tawakoni or Tahuacano or Tenino or Thawikila or Hathawekela or "Fort Ancient" or Tigua or Tillamook or Nehalem or Timbisha or Panamint or Timpanogos or Tlingit or "Toi Ticutta" or Tolowa or "Talawa Dini' " or Tongva or Gabrieleno or Fernandeno or Tobikhar or Tonkawa or Ticanwatic or Tsikip or Appalousa or Opelousa or Tsitsistas or Tubatulabal or Tukabatchee or Tuscarora or Tomahittan or Kuskarawock or Tutelo or Tutero or Totteroy or Tutera or Yusan or Tututni or Umatilla or Umpqua or Waccamaw or Waxmaw or Wadatika or "Harney Valley Paiute" or Wailiki or Waluulapam or "Walla Walla" or Walpapi or Huipui or Wampanoag or Massasoit or Wanapum or Wappo or Washoe or Wichita or Willapa or Kwalhioqua or "Wi pukba" or "Verde Valley Yavapai" or Wintu or "Northern Wintun" or Wiyot or "Wee' at" or Weyet or Yakama or "Yamosopo Tuviwarai" or Yaqui or Yoeme or Yatasi or Yattasih or "Yavbe' " or "Yavapai" or "Ysleta del Sur" or Yojuane or Yokuts or Mariposa or Yuki or Yupighyt or "Yup'ik" or Yupik or Yurok or "Olekwo'l" or Zuni).mp. or ((Applegate or Delaware or Iowa or Ishak or Kaw or Kato or Spokane or Miami or Arkansas or Tali or Tunica or (Han not (China or Chinese)) or Pawnee or "Coeur D' Alene" or Piscataway or Ree or Tula) adj3 (reservation* or nation or people or peoples or population or man or men or woman or women or child* or youth* or elder or elders or communit* or tribe or tribes or tribal or Indian*)).mp. [United States] | 55417 |
| 67 | ((Mexico/ or ((Mexico not "New Mexico") or Mexican or Aguascalientes or "Baja California" or Campeche or Chiapas or (Chihuahua not (dog or dogs or pet or pets)) or Coahuila or Colima or Durango or Guanajuato or Guerrero or Hidalgo or Jalisco or Michoacan or Morelos or Nayarit or "Nuevo Leon" or Oaxaca or Puebla or Queretaro or "Quintana Roo" or "San Luis Potosi" or Sinaloa or Sonora or (Tabasco not (sauce* or flavo*)) or Tamaulipas or Tlaxcala or Veracruz or Yucatan or Zatecas).mp.) and (exp indigenous people/ or (Mesoamerindian* or Indigen* or aborig* or "first people*" or "indos mexicano" or "original people*" or "pueblos indigenas").mp.)) or (Aguacatec or Akwa'ala or Abxubal or Ayuukja'ay or "Batzil k'op" or Binizaa or "Chichimeca Jonaz" or Chinantec or Chocho or "Ch ol" or Chontal or "Chuj" or Cochimi or Comcaac or Hamasipini or Harijio or "Ha shuta enima" or "Hach t'an" or Huastecor Hnahnu or Hnatho or Ixcatec or Ixil or Jacaltec or K'akchikel or K'anjobal or Kanjobal or Kaqchikel or Kechi or K'iche or Kikapooa or Kikapu or Kiliwa or "Ko'lew" or "K'op o winik atel" or Kumiai or Lacandon or Laymon or Makurawe or Maya or Maya'wiinik or Mazahua or Mazatec or Me'phaa or Mexicanero or Mexikatlajtolli or Mixe or Mixtec or Motocintleco or mti'pa or Nahuas or Ocuiltec or Otomi or (Oaxaca not Oaxaca-Blinder) or "Pame" or Papago or Tlahuica or Paipai or "Pima Bajo" or Purepecha or P'urhepecha or Qatok or (Quiche not Guatemala) or Q'iche or Raramuri or "Runixa ngiigua" or ("Seri" not "Seri 82") or "Slijuala sihanuk" or Tacuate or Tarahumara or Teenek or Tepehua or Ti'pai or Tlapanec or "Tohono O'odham" or Totonac or Tachiwin or "Tsa jujmi" or Tzotzil or "Tu'un savi" or Tzeltal or "Uza" or Winik or Xigue or Yucatec or Zapotec).ti,ab,kw. [Mexico] | 5476 |
| 68 | ((exp India/ or Bangladesn/ or Bhutan/ or Nepal/ or Pakistan/ or Sri Lanka/ or (India or Bangladesh* or (Bhutan* not bhutanensis) or (Nepal* not nepalensis) or Pakistan* or "Sri Lanka*").mp.) and (indigenous people/ or ((Indigenous* or tribe or tribal or tribes or Rai) adj3 (population* or people* or person* or elder* or man or men or woman or women or child* or youth* or clan or clans or tribe or tribes or tribal or family or families or parent* or grandparent* or elder or elders or grandmother* or grandfather* or baby or babies or infant or infants or patient or patients or speakers or speaking or village* or communit*)).mp.)) or ("Adivasis" or "Adnamanese" or "Andaman" or "Baluch" or "Baluchis" or "Bodo" or "Boro" or "Boros" or "Bote" or "Brahuis" or "Chakmas" or "Chepang" or "Chhantyal" or "Damai" or "Dewan" or "Ghale" or "Gurkha" or "Gurung" or "Hayu" or "Hyolmo" or "Jarawa" or "Jirel" or "Jumma" or "Kalash" or "Khas" or "Kirati" or "Koinch" or "Kulung" or "Kusunda" or "Limbu" or "Lohorung" or "Magar" or "Makrani" or "Mangar" or "Marma" or "Miji" or "Mongar" or "Mro" or "Naga" or "Nepami" or "Newar" or "Nicobar" or "Onge" or "Rang" or "Raute" or "Sajolang" or "Santhal" or "Sentinelese" or "Sindhis" or "Sulemani" or "Sunuwar" or "Tamang" or "Thakali" or "Thangmi" or "Tharu" or "Tripura" or "Tumbahangphe" or "Wanniyala-Aetto" or "Yakkha" or "Yolmopa").ti,ab. [Indian Sub-Continent] | 9741 |
| 69 | Indians, South American/ or (Abipon or Achuar or Achuagua or Akawaio or Amarizana or Andoque or Akawaio or Akuriyo or Anauya or Araona or Arawak or Ayamn or Aguaruna or Amahuaca or Amarakaeri or Andoa or Arabela or Arawak or Arhuaco or Ashaninca or Asheninca or Atsahuaca or Aymara or Ayoreo or Bakairi or "Baniva" or Barasana or Baniwa or "Baure" or Bororo or Cabiyari or Cacataibo or Caquinte or Cacua or Cahuarano or "Caiua" or "Camara Indians" or Camaracoto or Camsa or Canamari or Candoshi or Canela or Canichana or Capanahua or Carapana or Cariay or "Carib" or Carijona or Carutana or Cashibo or Cashinahua or Cawishana or Cavinena or Caxuiana or Cayuvava or Chontaquiro or Cocama or "Cubeo" or Curipaco or Chacobo or Chaima or (Chana not striatus) or Chapacura or Charrua or Chimila or Chitonahua or Chorote or Chipaya or Chiquitano or Chulupi or Carare or Coconuco or Cofan or Coreguaje or Coyaima or Chamacoco or Chamicuro or Chayahuita or Cocama or "Culina" or Culino or Cubeo or Cuiba or "Cuiva" or Cumanagoto or Curripaco or "Deni" or Desano or Embera or Guarani or Guajajara or "Guana" or Guanano or Guarayo or Guarayu or Guahibo or Guajiro or Guambiano or Guanano or Guayabero or Guarequena or Guinao or "Guana" or Gayon or Guahibo or Hixkaryana or Huachipairi or Huambisa or Huarayo or Lauanaua or Ikpeng or Ingariko or Irantxe or Itonama or Inapari or Iquito or Isconahua or Jumana or Japreria or Jirajara or Juruti or Jaqaru or Jebero or Kadiweu or Kaingang or Kamayura or Karaja or Karipuna or "Kariri" or Katukina or Kaxarari or Kayabi or Kayapo or "Kuikuro alapalo" or Kulina or "Kaiwa" or Kallawaya or "Kogui" or "Kuna" or Kaweskar or "Lule" or Macuna or Maipure or Mapuche or Mataco or Mocovi or Machinere or Machinerev or Machiguenga or Macushi or Macuna or "Madi" or Malayo or Mamainde or Manao or Mandauaca or Mandawaka or Mapidian or Mapuche or Mapidian or Maquiritare or Maquiritari or Maragua or Marawan or Mariate or Marubo or Mastanahua or Mataco or Matipuhy or "Matis" or "Matses" or Mawakua or Mawakwa or Maxakali or Mehinaku or Miranha or Moronawa or Munduruku or Movima or Muellama or Muinane or Mapoyo or "Mashco Piro" or Muniche or Nambikwara or Nocaman or Nuquini or Nomatsiguenga or Nanti or Ocaina or Omagua or Orejon or "Opon" or Pacahuara or "Paez" or Paicone or Palicur or Panare or "Pano" or "Paresi" or Paumari or "Pemon" or Pilaga or Puelche or Pauna or Pauserna or Piapoco or Piraha or Piratapuyo or Pisabo or Piaroa or "Pijao" or Piratapuyo or Paraujano or Pemon or Pemono or Piapoco or Puinave or Patamona or Poyanawa or Puinave or Puquina or Quechua or Quichua or Retuara or Resigaro or Reyesano or Sabanes or "Saliba" or Saluma or Sarave or Secoya or Selknam or "Sensi" or Shaninawa or Shapra or Sharanahua or Shebayo or Shiwiar or Shikiana or Sikiana or Siriono or Sinsiga or "Siona" or Suruwaha or Tacano or Tamanaco or Tiahuanaco or Tariano or Tehuelche or Tariano or Tatuyo or "Tembe" or "Terena" or Telembi or Ticuna or Ticuna or Tiriyo or Tiwanaku or Tiwanaku or "Torom" or "Totoro" or Tsimane or Tuberao or "Tucano" or Tunebo or Tuxinawa or Tuyuca or Uainuma or Urarina or Vilela or Waimaha or Waiampi or Waiwai or Wapishana or Waraiku or Warekena or Waura or Wayampi or Wayana or Wirina or Waimaha or Waunana or "Wiwa" or "Warao" or Wayuu or Witoto or Xavante or Xipaya or Xiriana or Xokleng or Yabaana or Yaminawa or Yaminahua or Yaruma or Yawalapiti or Yuracare or Yabarana or Yavitero or "Yine" or Yamana or Yaghan or Yucuna or Yurumangui or Yukpa or Yanesha or Yoranahua or Yagua or Yaminahua or Zaparo or Zamuco or "Trio Indians" or "More Indians" or "Bare Indians" or (("Inga" or "Maca" or "Leco" or "Mojo" or "Uro" or "Maco" or Lengua or "Toba" or "Zoe" or "Ona" or "Catio" or "Passe" or "Bari" or "Awa" or "Bora" or "Bara" or "Remo" or "Pano" or "Sape") adj3 (Indians or Indian or Indigenous or Amerindian* or Aborigin* or people or peoples or elder or elders or grandmother* or grandfather* or parent* or women or men or woman or man or child* or youth or youths or baby or babies or tribe or tribes or tribal or shaman* or native or patient or patients))).ti,ab. or ((Indian* or Amerindian, or Aboriginal* or Indigenas or Indigenous or tribe or tribes or tribal).ti,ab. and (exp South America/ or "South America*".mp. or Argentin*.mp. or Bolivia*.mp. or Brazil*.mp. or Chile*.mp. or Colombia*.mp. or Ecuador*.mp. or "French Guiana".mp. or Guyana*.mp. or Paraguay*.mp. or Peru.mp. or Peruvian.mp. or Suriname.mp. or Uruguay*.mp. or Venezuela*.mp. or "Amazon Region".mp. or Amazonia.mp. or Andes.mp. or Andean.mp.)) [South America] | 34121 |
| 70 | ((Ainus or Ainu or Aleuts or Alyutors or Chukchis or Chuvans or Dolgans or Enets or Entsy or Yupik or Yup'ik or Yuit or Yupigyt or Chaplino or Naukan or Itelmens or Kamchadals or Kereks or "Komi" or Koryaks or Nenets or Nentsy or Nganasans or Tavgi or Sami or Veps or Yukaghirs or Chulyms or Evenks or Tungus or Evens or "Kets" or Khantys or Mansi or Vguls or Selkups or Teleuts or Nanais or Nanaitsy or Negidal or Nivikh or Oroch or orok or Taz or udege or ulch or Kumadins or Chelkans or Shorians or Soyots or Telengits or Tofalars or Tugalars or "Tufans Todzhins" or Laks or Tabasarans or Turuls or Aguls or Tsakhurs or Kumyks or Nogais or "Andis" or Akhvakh or Archins or Bagvalals or Bezhta or Botlikhs or Chamalals or Godoberi or Hinukh or Hunzibs or Khwarshi or Karata or Tindis or Tsez or Abazin or Besermyan or Izhorians or Karelians or Nagaybaks or Setos or Shapsugs or Quratay).mp. or (exp Indigenous people/ or (Indigenous adj3 (population* or people* or person* or elder* or man or men or woman or women or child* or youth* or clan or clans or tribe or tribes or tribal or family or families or parent* or grandparent* or elder or elders or grandmother* or grandfather* or baby or babies or infant or infants or patient or patients or speakers or speaking or village* or communit*)).ti,ab.)) and (Russia*.mp. or exp Russia/) | 1158 |
| 71 | ((Greenland/ or Greenland*.mp. or "Kalaallit Nunaat".mp. or Nuuk.mp. or Sisimiut.mp. or Ilulissat.mp. or Qaqortoq Aasiaat.mp. or Maniitsoq.mp. or Tasiilaq.mp. or Uummannaq.mp. or Narsaq.mp. or Paamiut.mp. or Nanortalik.mp. or Upernavik.mp. or Qasigiannguit.mp.) and (exp indigenous people/ or Inuit.mp. or Inuk.mp. or ((Indigen* or Aborigin*) adj3 (population* or people or peoples or person or persons or elder or elders or man or men or woman or women or child* or youth* or clan or clans or tribe or tribes or tribal or family or families or parent* or grandparent* or elder or elders or grandmother* or grandfather* or baby or babies or infant or infants or patient or patients or speakers or speaking or village* or communit*)).mp.)) or (Greenlandic or Kalaallit or Kalaallisut or Tunumiit or Inughuit or Avanersuarmiut).mp. [mp=title, abstract, heading word, drug trade name, original title, device manufacturer, drug manufacturer, device trade name, keyword heading word, floating subheading word, candidate term word] | 1082 |
| 72 | ((exp Asia, Southeastern/ or ("Bali" or "Borneo" or "Brunei" or Cambodia or "Flores Island" or "Indonesia" or Java or Komodo or "Laos" or "Lombok" or "Malasia" or "Malay Peninsula" or "Myanmar" or "Mekong Valley" or "New Guinea" or "Philippeans" or "Sabah" or "Singapore" or "Sumatra" or "Sumba" or "Sumbawa" or "Thailand" or "Timor" or "Vietnam").mp.) and (indigenous people/ or ((Indigen* or aborig*) adj3 (population* or people or peoples or person or persons or elder or elders or man or men or woman or women or child* or youth* or clan or clans or tribe or tribes or tribal or family or families or parent* or grandparent* or elder or elders or grandmother* or grandfather* or baby or babies or infant or infants or patient or patients or speakers or speaking or village* or communit*)).mp.)) or ("Abui" or "Aeta" or "Alfur" or "Ati People" or "Bahau" or "Bali Aga" or "Bali Mula" or "Baliaga" or "Balinese" or "Basap" or "Bataq" or "Batek" or "Bateq" or "Batin" or "Blaan" or "Brao" or "Bru" or "Bru-Van Kieu" or "Bugkalot" or "Bumiputera" or "Bunong" or "Chams" or "Champa" or "Cheq Wong" or "Dasun" or "Dayak" or "Deli Malay" or "Filipina*" or "Filiopino*" or "Gaddang" or "Ibaloi" or "Iban" or "Idan" or "Igorot" or "Jahai" or "Jehai" or "Jakun" or "Javanese Kshatriya" or "Kangeanese" or " Kagayanen" or "Kanayatn" or "Katu" or "Kayan" or "Kavet" or "Kenyah" or "Khmer Krom" or "Kimaragang" or "Klemantan" or "Kreung" or "Kuijau" or "Kuy" or "Kwijau" or "Lampung*" or "Lanoh" or "Lawangan" or "Luangan" or "Lumad" or "Dayak" or "Loloan Malay*" or "Mah Meri" or "Malay Singaporean*" or "Malbog" or "Maluku" or "Mangka'ak" or "Mangyan" or "Maniq" or "Maragang" or "Ma'anyan" or "Minokok" or "Moluccas" or Montagnard or "Murut" or "Naga" or "Negrito" or "New Guinea" or "Ngaju" or "Orang Hulu" or "Orang Kanaq" or "Orang Kuala" or "Orang Seletar" or "Orang Ulu" or "Ot Danum" or "Palawano" or "Palembang*" or " Panay-Bukidnon" or "Pan-ayanon" or "Papua" or "Phnong" or "Punong" or "Pear People" or "Por People" or "Punan" or "Rumanau" or "Sasak" or "Semai" or "Semang" or "Semaq Beri" or "Semelai" or "Senoi" or "Simeulue" or "Suludnon" or "Sunda Island*" or "Tabanwa" or "Takua" or "Tambanuo" or " Tombonuo" or "Taron" or "Temiar*" or "Temoq" or "Temuan" or "Tidung" or "Trone" or "Taaw't Bato").ti,ab. | 11642 |
| 73 | (exp indigenous people/ or ((Indigen* or aborig*) adj3 (population* or people or peoples or person or persons or elder or elders or man or men or woman or women or child* or youth* or clan or clans or tribe or tribes or tribal or family or families or parent* or grandparent* or elder or elders or grandmother* or grandfather* or baby or babies or infant or infants or patient or patients or speakers or speaking or village* or communit*)).mp.) and (pacific islands/ or exp melanesia/ or exp micronesia/ or exp polynesia/ or (Micronesia or "Caroline Island" or Guam or Kiribati or "Mariana Island*" or Nauru or Palau or Melanesia or "East Nusa Tenggara" or Fiji or Fijian or "Loyalty Island*" or "Maluku Islands" or Moluccas or "New Caledonia*" or "New Guinea" or Papua or Rotuma or "Solomon Island*" or Vanuatu or Polynesia or "Austral Island*" or "Cook Island*" or "Chatham Islands" or "Easter Island*" or Hawaii* or "Marquesas Island" or "Norfolk Island" or Samoa or Tahiti or Tokelau or Tonga or Tuvalu or Tuamotu or Kermadec or Banabans or "Native Hawaiian*" or Guamanian or Chamarro* or Chuukese or I-Kiribati or Kosraean* or Maohi or Marshallese or Moriori or Mortlockese or Mwokilese or Namonuito or Nauruans or Ngatikese or Niuean or Paafang or Paluans or Pingelapese or Pohnpeian* or Pollapese or Puluwat or Rapanui or Rotuman or Satwalese or Samoan*or Sonsorolese or Tahitian* or Tobian or Tongan or "Torres Strait Islander*" or Tuvaluan or Ulithian or Woleian or Yapese or Polynesian* or Micronesian* or Melanesian*).ti,ab.) | 3911 |
| 74 | ((exp indigenous people/ or ((Indige* or aborig* or nomadic or seminomad* or "semi nomad*") adj3 ("people" or "peoples" or "elder" or "elders" or "grandmother*" or "grandfather*" or "parent*" or "women" or "men" or "woman" or "man" or "child*" or "youth" or "youths" or "baby" or "babies" or "tribe" or "tribes" or "tribal" or "shaman*" or "native" or "patient*")).mp.) and (("Middle East" or "Afghanistan" or "Bahrain" or "Iran" or "Iraq" or "Israel" or ("Jordan" not "Jordan's Principle") or "Kuwait" or "Lebanon" or "Oman" or "Palestine" or "Qatar" or "Saudi Arabia" or syria or syrian).mp. or middle east/ or bahrain/ or cyprus/ or iran/ or exp iraq/ or israel/ or jordan/ or kuwait/ or lebanon/ or oman/ or palestine/ or qatar/ or saudi arabia/ or syrian arab republic/ or "turkey (republic)"/ or exp united arab emirates/ or yemen/)) or ("Bedouin*" or "Jahalin" or "al-Kaabneh" or "al-Azazmeh" or "al-Ramadin" or "al-Rshaida").ti,ab. | 1711 |
| 75 | or/60-74 | 135733 |
| 76 | 59 and 75 | 1155 |
| 77 | animal experiment/ | 3111405 |
| 78 | 76 not 77 | 1137 |
| 79 | limit 78 to dc=20230701-20240214 | 81 |

**Global Health <1910 to 2024 Week 05>**

| **#** | **Search Statement** | **Results** |
| --- | --- | --- |
| **1** | **urinary incontinence/ or urinary incontinence, stress/ or urinary incontinence, urge/** | **1119** |
| **2** | **rectal prolapse/ or uterine prolapse/ or vaginal prolapse/** | **254** |
| **3** | **constipation/** | **3962** |
| **4** | **intestinal cancer/ or anal cancer/ or colorectal cancer/ or rectal cancer/** | **22362** |
| **5** | **cervical cancer/ or endometrial cancer/ or uterine cancer/ or uretheral cancer/** | **22178** |
| **6** | **((gyn?eologic* or rectal or rectum or genital or prostat* or unterine or uterus or ovarian or endometrial) adj3 (surgery or surgical)).mp.** | **617** |
| **7** | **4 or 5 or 6** | **43649** |
| **8** | **exp postoperative complications/ or (postoperative or "post operative" or "post surg*").mp.** | **36151** |
| **9** | **7 and 8** | **1129** |
| **10** | **urinary tract diseases/ or pelvic inflammatory disease/ or urinary tract infections/ or urination disorders/ or polyuria/** | **20074** |
| **11** | **(prolaps* adj3 ("pelvic organ" or uterine or uterus or bladder or rectal or rectum or urethra* or apical)).mp.** | **691** |
| **12** | [**incontinence.mp**](http://incontinence.mp/)**.** | **2635** |
| **13** | **(uterocele or cystocele or rectocele or enterocele or urethrocele or sigmoidocele).mp.** | **65** |
| **14** | **(bladder* adj3 (overactive or urgency or urgent or frequency or heistency or retention)).mp.** | **367** |
| **15** | **(bowel adj3 (frequency or urgency)).mp.** | **298** |
| **16** | [**constipation.mp**](http://constipation.mp/)**.** | **6849** |
| **17** | [**encopresis.mp**](http://encopresis.mp/)**.** | **35** |
| **18** | **(dyssynergia adj3 (bowel* or bladder*)).mp.** | **3** |
| **19** | **"vaginal wind".mp.** | **1** |
| **20** | **"pelvic girdle pain".mp.** | **29** |
| **21** | **(dyspareunia or vulvodynia or vaginismus or vestibulodynia or "interstitial cystitis" or "painful bladder syndrome" or "proctalgia fugax" or prostatitis or endometriosis or adenomyosis or "pudendal neuralgia" or anismus).mp.** | **3675** |
| **22** | **Neurogenic** [**bladder.mp**](http://bladder.mp/)**.** | **186** |
| **23** | **("sexual dysfunction" or "erectile dysfunction" or "persistent genital arousal" or impostence).mp.** | **2316** |
| **24** | **("bed wett*" or bedwett* or encopresis or "hirshprung* disease").mp.** | **122** |
| **25** | **("gender confirmation surg*" or "sex reassignment surg*").mp.** | **25** |
| **26** | **"lichen sclerosis".mp.** | **13** |
| **27** | [**nocturia.mp**](http://nocturia.mp/)**.** | **331** |
| **28** | **((fistula* or fistulae) adj3 (bladder* or urinary or vagina* or rectovaginal* or vesicovaginal*)).mp.** | **353** |
| **29** | **"levator ani avulsion".mp.** | **1** |
| **30** | **("Lower urinary tract symptom*" or dysuria or "LUTS").mp.** | **1744** |
| **31** | **"voiding dysfunction".mp.** | **86** |
| **32** | **1 or 2 or 3 or 9 or 10 or 11 or 12 or 13 or 14 or 15 or 16 or 17 or 18 or 19 or 20 or 21 or 22 or 23 or 24 or 25 or 26 or 27 or 28 or 29 or 30 or 31** | **37661** |
| **33** | **((exp Indians, North American/ and Canad*.mp.) or exp inuit/ or exp Health Services, Indigenous/ or exp Ethnopharmacology/ or (Athapaskan or Saulteaux or Wakashan or Cree or Dene or Inuit or Inuk or Inuvialuit* or Haida or Ktunaxa or Tsimshian or Gitxsan or Gitksan or "Nisga'a" or Haisla or Heiltsuk or Oweenkeno or "Kwakwaka'wakw" or "Nuu chah nulth" or "Tsilhqot'in" or Dakelh or "Wet'suwet'en" or Sekani or Dunne-za or Dene or Tahltan or Kaska or Tagish or Tutchone or Nuxalk or Salish or St'at'imc or Stl'atl'imx or Stl'atl'imc or Nlaka'pamux or Okanagan or "Sec wepmc" or Secwepemc or Tlingit or Anishinaabe or Blackfoot or Nakoda or Tasttine or "Tsuu T'ina" or "Tsuut'ina" or "Gwich'in" or (Han not (China or Chinese)) or Algonquin or Nipissing or Ojibwa or Potawatomi or Innu or Maliseet or "Mi'kmaq" or Micmac or Passamaquoddy or Haudenosaunee or Cayuga or Mohawk or Oneida or Onondaga or Seneca or Tuscarora or Wyandot or Aboriginal* or Indigenous* or Metis or red road or "on reserve" or off-reserve or First Nation or First Nations or Amerindian).mp. or (urban adj3 (Indian* or Native* or Aboriginal*)).mp. or** [**ethnomedicine.mp**](http://ethnomedicine.mp/)**. or country food*.mp. or residential school*.mp. or ((exp Medicine, Traditional/ or traditional medicine*.mp.) not Chinese.mp.) or exp Shamanism/ or shaman*.mp. or traditional heal*.mp. or traditional food*.mp. or medicine** [**man.mp**](http://man.mp/)**. or medicine** [**woman.mp**](http://woman.mp/)**. or autochtone*.mp. or (Native* adj1 (man or men or women or woman or boy* or girl* or adolescent* or youth or youths or person* or adult or people* or Indian* or Nation or tribe* or tribal or band or bands)).mp.) and (exp Canada/ or (Canad* or British Columbia or Colombie Britannique or Alberta or Saskatchewan or Manitoba or Ontario or Quebec or Nova Scotia or New Brunswick or Newfoundland or Labrador or Prince Edward Island or Yukon Territory or NWT or Northwest Territories or Nunavut or Nunavik or Nunatsiavut or NunatuKavut).mp.) [Canada]** | **4416** |
| **34** | **((exp australia/ or (australia* or northern territory or tasmania or new south wales or Victoria or queensland).ti,ab.) and (aborigines/ or aborigin*.ti,ab. or indigenous.ti,ab.)) or "torres strait islander*".ti,ab. [Australia and Torres Strait]** | **6460** |
| **35** | **(Maori or "tangata whenua").mp. or ((exp New Zealand/ or (New Zealand or Aukland).mp.) and ("Native Hawaiian or Other Pacific Islander"/ or (Aborig* or Indig*).ti,ab.)) [Maori]** | **2270** |
| **36** | **(Saami or Sampi or (Sami not Ulus) or Samis or Southernsami* or Umesami* or Pitesami* or Lulesami* or Northernsami* or Enaresami* or Kolasami* or Lapp or Lapps or Lappish or Lappland or (Lapland* not longspur) or Lappalainen* or Saamelainen* or reindeer herd* or reindeer culture* or reindeer pastoral* or Lappbys or Samebys or reinbeitesdistrikt or paliskunta or siida).mp. or (((Fennoscandia or Finnmark or Scandinavia or Nordic or Sweden or Norway or Finland or Swedish or Finnish or Norwegian or Norge or Svensk* or Suomi or Barents Region or (Kola not (garcinia or gotu)) or Arctic Europe* or Polar Europe* or North* Europ*).mp. or Finland/ or Norway/ or Sweden/) and ((traditional adj3 (food* or heal* or medicine* or shaman*)) or (Indigenous* adj3 (people* or person* or mother* or father* or parent* or grandparent* or grandmother* or grandfather* or elder or elders or child* or boy or boys or girl* or youth* or healer* or patient or patients or famil* or herder* or village* or communit*))).mp.)** | **1069** |
| **37** | **((exp indigenous people/ or "indigenous people* ".mp.) and (exp Africa/ or ((Africa* not "African American*") or Algeria or Angola or Benin or Botswana or "Burkina Faso" or Burundi or Cameroon or "Cape Verde" or "Cabo Verde" or "Central African Republic" or Chad or Comoros or Congo or Djibouti or Egypt or "Equitorial Guinea" or Eritrea or Eswatini or Ethiopia or Gabon or Gambia or Ghana* or Guinea or "Guinea Bissau" or "Ivory Coast" or Kenya or Lesotho or Liberia or Libya or Madagascar or Malawi or Mali or Mauritania or Mauritius or Mayotte or Morocco or Mozambique or Namibia or Niger or Nigeria or Reunion or Rwanda or "Saint Helena" or Ascension or "Tristan de Cunha" or "Sao Tome" or Principe or Senegal or Seychelles or "Sierra Leone" or Somalia or "South Africa" or Sudan or Tanzania or Togo or Tunisia or Uganda or "Western Sahara" or Zambia or Zimbabwe).mp.)) or (Abakuria or Abaluhya or Abagusii or Abakuria or Aembu or Agikuyu or Akamba or Anuak or Anywaa or Amazigh or Ambala or Ambeere or Ambundu or Ambuun or Amharan or Angba or Baaka or Baamba or Babindi or Babini or Baboma or Bachokwe or Bacwa or Bafumbira or Baganda or Bagyele or Bagwere or Bagyeli or Bakiga or Bakola or Baholo or Bakalanga or Bakiga or Bakolo or Bakongo or Bakonjo or Baluba or Balunda or Balovale or Bamasaba or Bambuti or Bangala or Bangoli or Bangungu or Bantu or Banyankole or Banyarwanda or Banyole or Banyoro or Bapende or Bapedi or Barabaig or Barombi or Barundi or Baruuli or Basamia or Basoga or Batswana or Batooro or Batsamba or Batswana or Batwa or Bayaka or Bedzan or Bazombe or Bebayaka or Bedzan or Bhaca or Biaka or Borana or Chewa or Copts or Cormorian or Cushitic or Dahalo or Datooga or Dikidiki or Dogon or Ewondo or Fulani or Fuliru or Ganguela or Gciriku or Gyele or Hadza or Hadzabe or Haillom or Haratin or Herero or Himba or Hlubi or Iriryen or Iqvayliyen or Kabyle* or Kalenjin or Kanioka or Kanioka or Kaonde or Karamojong or Kavango or Kereuyu or Khoikhoi or KhoiSan or Kikuyu Kwangali or Lokele or Lowme or Lotuko or Lwalwa or Maasai or Makonde or Makua or Mande or Masalit or Matumbi or Mayeuyi or Mayeyi or Mbenga or Mbukushu or Mbochi or Mboro or Mbuti or Medzan or Mijikenda or Mozabite* or Nafusa or Ndebele or Ngombe or Namaqua or Nyanga or Nyamwezi or Ogiek or Ovambo or Ovimbundu or Phuthi or Pokomo or Rendille or Riffian or Riffians or Sakuma or Samburu or Sandawe or Sangha or Sango or Sengwer or Serer or Sesotho or Shangaan or Shawiya or Shenwa or Shi or Shilluk or Sukua or Sukus or Swahili or Tabwa or Tambuka or Taveta or Thembu or Tigrayan or Topoke or Tsonga or Toubou or Tuareg or Tumbuka or Ugana or Wochua or Xhosa or Xindonga or Yoruba or Zenati or Zuwara or ((Indigenous or Afar or Afars or "Aka People" or Akie or Ameru or Asua or Ateker or Atwot or Awjila or Bafia or Baka or Bakongo or Bakwe or Balunda or Balovale or Bango or Bassa or Beja or Bekpak or Bemba or Bembe or Benet or Berber or Berbers or Bira or Bowe or Bubi or Budja or Bulu or Bunrun or Chaga or Chopi or Damara or Dinka or Djerba or Duala or Dzing or Efe or Elmolo or Fang or Foora or Fula or Fur or Ghomara or Ghadames or Gllana or Glu or Gogo or Gongo or Haya or Havu or Hema or Hima or Hunde or Hutu or Huva or Iboko or Igbo or Ijo or Jieng or Kadu or Kande or Kango or Katla or Kgaga or Khoe or Kola or Komo or Kota or Kua or Kuba or Kwango or Kx'z or Kxoe or Lala or Lozi or Luo or Luba or Lupu or Masmuda or Matmata or Mbala or Mbam or "Mbo People" or Mbolo or Mbuza or Mongo or Mpondo or Myene or Naadh or Nama or Nande or Naro or Ngoni or Ndau or Ndebele or Ngoli or Ngondi or Ngoni or Nguni or Nkoya or Nkumu or Nuba or Nubian or Nuer or Nzebi or Ogoni or Omoro or Oroko or Pygmy or Popoi or Poto or Puru or Rashad or (San not ("San Francisco" or "San Diego" or "San Antonio")) or Sango or Sanhaja or Sena or Shilha or Shira or Shona or Shua or Sokna or Somali* or Sotho or Sua or Subu or Swazi or Taitaa or Tchokwe or Teke or Tembo or Tetela or (Tonga and Africa*) or Tshwa or Tsoa or Twa or Turkana or Tuu or Venda or Vira or Watta or Wakuti or Yaaku or Yaka or Yakoma or Yanzi or Yao or Yeke or Yela or Yeyi or Zulu) adj3 (population* or people* or person* or elder* or man or men or woman or women or child* or youth* or clan or clans or tribe or tribes or tribal or family or families or parent* or grandparent* or elder or elders or grandmother* or grandfather* or baby or babies or infant or infants or patient or patients or speakers or speaking or village* or communit*))).mp. [Africa]** | **30464** |
| **38** | **(((Acatec or Aguacateco or Amuzgo or Bokota or Boruca or Bribri or "Bri Bri" or Buglere or Cabecar or Cakchiquel or Changuena or Chatino or Chiapanec or Chicomuceltec or Chinantee or Chocho or Cholti or "Ch'olti'" or "Ch'olti'anor Chontal" or Chorotega or Chorti or Chuj or Chumbia or Corobici or (Cueva not Spain) or Cuicatec or Cuitlatee or Cuytec or Dorasque or Embera or Garifuna or Guatuso or Guaymi or Guaymis or Guetar or Huastec or Huave or Huetar or Itzaj or Ixil or Jacalteco or Jonaz or Kanjobal or Kekchi or Kuna or Maleku or Mangue or Matambu or Matlatzinca or Mazahua or Motozintlec or Mayan or Mayangna or Miskito or Mixtec or Mopan or Nahua or Nahuatl or Ngabe or Otomi or Pantec or Paya or Popoloca or Popoloc or Poqomam or Poqomchi or "Q'eqchi'" or Quiche or Quitirrisi or Sacapulteco or Sipacapense or Subtiaba or Tacaneco or Tarasco or Tamaulipec or Tamazultec or Tecoxquin or Tectiteco or Tecual or Tecuexe or Tepehura or Tepuztecor or Teribe or Terraba or Totonac or Trique or Tzeltal or Tzotzil or Tzutujil or Ulwa or Uspantec* or Uspanteko or Voto or Xinca or Waunana or Wounaan or Yucatec or Zapotec or Zoque or (("Costa Rica*" or Hondura* or Nicaragua* or Panama* or Guatemala* or Achi or Belize or Belizean* or Maya* or Mixe or Pame or Pipil or Pech or Chol or "Ch'olan" or Cora or Cuna or Mam or Rama) adj5 (Indian or Indians or Amerindian* or Amerindio* or Aboriginal* or Indigenous or Indigena* or Aborigen* or Mestizo or tribe or tribes or tribal or "traditional medicine*" or shaman*))).tw. or (Belize or "Costa Rica" or "El Salvador" or Guatemala or Honduras or Nicaragua or Panama).mp.) and (exp indigenous people/ or (indigenous adj3 (population* or people* or person* or elder* or man or men or woman or women or child* or youth* or clan or clans or tribe or tribes or tribal or family or families or parent* or grandparent* or elder or elders or grandmother* or grandfather* or baby or babies or infant or infants or patient or patients or speakers or speaking or village* or communit*)).mp.)) not ((Mexico or Mexican).mp. or exp Mexico/) [Central America]** | **336** |
| **39** | **Indigenous People/ or American Indian/ or Canadian Aboriginal/ or Eskimo/ or Inuit/ or Indigenous Health Services/ or ("A' ani" or Absaroka or Haaninin or Atsina or "Gros Ventre" or Acopsel or Tlacopsel or Lacopsel or Ahtna or Ahtena or Akenitsi or Occaneechi or Akokisa or Horcoquisa or Orcoquizas or Aleut or Unangax or Unangan or Alibamu or "Alabama Alsea" or Alutiiq or Sugpiag or Amahami or Awaxawi or Androscoggin or Arosaguntacook or Ameriscoggin or Anishinaabeg or Chippewa or Anihsinape or Saulteaux or Apalachee or Aranama or "Texan Coahuilteca" or Tamique or Arikara or Sahnish or Arickaree or Adakadaho or Assiniboine or Hohe or Nakota or Nakoda or Nakona or "Atsa' Kudok-wa" or Awatixa or Bannock or "Snake Indian*" or Bidai or Quasmigdo or Biloxi or Blackfoot or Niitsitapi or Sikasikaitsitapi or Cahto or Kaipomo or Cahuilla or Ivilyuqaletem or Ivilyuat or Catawba or Inna or Iswa or Chemehuevi or Chickasaw or "Chilula Chimakum" or Aqokulo or Chimariko or Chiricahua or Tsokanende or Chitimacha or Chetimachan or Sitimacha or Chowanoke or Roanoke or Chumash or Ciboney or "Taino Ciwat" or Clatsop or Coos or Coosa or Uchis or Chiaha or Coste or Talisi or Coquille or Kokwell or Coso or Cowlitz or Taitnapam or "Crow Nation" or "Cui Ui Ticutta" or Cupeno or Kuupangaxwichem or Cupa or "Cup' ig" or Nunivak or "Dakota Oyate" or Lakota or Nakota or Santee or Teton or Sioux or Deadose or "Deg Xina" or "Deg Xit' an" or Kaiyuhkhotana or "Deg Hit' an" or "Dena' ina" or Tanaina or "Dichinanek' Hwt' ana" or "Upper Kuskokwim Athabascan*" or Kolchan or Goltsan or "Tundra Kolosh" or "Do lkabya" or Duwamish or Esselen or Eyak or "Gidi' tikadi" or Guwevkabaya or "Gwich' in" or Kutchin or Haida or Xaadas or Xaat or Halchidhoma or Havasupai or "Green Water People" or Hiratsa or Hiraaca or "Ho-chaaqa" or Winnebago or Holikachuk or Innoko or "Tlegon-khotana" or Hopi or "Houma-Louisiana" or Huaco or Waco or Hualapai or Hupa or Natinixwe or "Natinook-wa" or "Hwech' in" or Hankutchin or "Iroquois Confederacy" or "Hodinoso ni" or "Illinois Confedera*" or Ilinoweg or Illini or Inupiat or Inuit or Ioway or Baxoje or Jicarilla or Juaneno or Acjachemen or Jumano or Kalapuya or Clackama or Kalispel or "Pend d' Oreilles" or Qlispe or Karuk or Karok or "Chum-ne" or Katkoc or Kansa or Kanza or Kawaiisu or Nuwa or Kennebec or "Kinipekw Kittitas" or Klickitat or "Qwu' lh-hwai-pum" or "Awi-adshi" or Mahane or Wahnookt or "Koa' aga' itoka" or Keresan or Kichai or Kitsai or Keechi or "K' itaish" or Kiowa or Gaigwu or Cauigu or Kutjau or "Kwu-da" or "Tep-da" or Kitanemuk or Kittitas or Klickitat or "Qwu' lh-hwai-pum" or "Awi-adshi" or Mahane or Wahnookt or "Koa' aga' itoka" or Konkow or "Koop Ticutta" or Koyukon or Ktunaxa or Kootenai or Flathead or Kucadikadi or "Kotsa' va" or Kumeyaay or "Tipai-Ipai" or Kamia or Diegueno or Kwapa or Cocopah or Cucapa or "Xawitt kwnchawaay" or Lassik or Lenape or "Leni-Lenape" or Lipan or Luiseno or Payomkawichum or Madqwadabaya or "Desert Yavapai" or Mahican or Mohicans or Makah or Makuhadokado or Maliseet or Wolistoqiag or Manahoac or Mahock or Meipontsky or Mandan or Mattole or "Bear River" or "Tul' bush" or "Ni' ekeni" or Meherrin or Menominee or Mackinac or Mescalero or Myaamiaki or Kickapoo or Twigtwee or Missouria or Miwok or Miwuk or Moadokado or Modoc or Mohave or "Aha Makhav" or Mohawk or "Kaneng' hega" or Molala or Molale or Molele or Nyyhmy or Moosonee or "Moose Cree" or Monsonis or Multnomah or Chinook or Nabedache or Nabaydacu or Wawadishe or Nabiltse or Dakubetede or "Nacho Nyak Dun" or Tutchone or Nacono or "Na' isha" or Nanticoke or Navajo or Ndee or Nial or Niimiipu or "Nez Perce" or Watapala or Watapahlute or Nisenan or Nisqually or Nomlaki or Noamlakee or "Central Wintun" or Nongatl or Nottoway or Cheroenhaka or "Northern Cheyenne" or Ohlone or Costanoan or Omaha or "O' odham" or Pima or Papago or Osage or Otoe or Otse or "Ozav Dika" or Palus or Passamaquoddy or Pestomuhkati or Patiri or Petaros or Pastia or Patwin or "Southern Wintun" or Panis or Skidi or Pedee or Penobscot or "Petun Piipaash" or "Kokmalik' op" or Piscatawa or Doeg or Conoy or "Pit River" or Pomo or Kashaya or Ponca or Ponka or Pottawatomi or Bodewadmik or Powhatan or Puyallup or Spuyalepabs or Quapaw or Ugahxpa or Quechan or Yuma or Kwtsaan or Quileute or Salinan or Saponi or Monacan or Sapon or "Eastern Blackfoot" or Christanna or Sawawatodo or Serrano or Taaqtam or "Maarenga' yam" or Yuhaviatam or Shasta or Chasta or Sasti or Shoshone or Siletz or Sinkine or Sinkyone or "Siuslaw Umpqua" or Skitswish or "Schitsu' umash" or Snohomish or Snuqualmi or Sokoki or Missiquoi or Stillaguamish or Stoluckwamish or Suquamish or Sutaio or Swinomish or Skagit or Syilx or Okanagan or Sotaae or "Taga Ticutta" or Takelma or Dagelma or Taltushtuntede or Galice or "Tanan Gwich' in" or Taos or Taovaya or Tataviam or Alliklik or Tawakoni or Tahuacano or Tenino or Thawikila or Hathawekela or "Fort Ancient" or Tigua or Tillamook or Nehalem or Timbisha or Panamint or Timpanogos or Tlingit or "Toi Ticutta" or Tolowa or "Talawa Dini' " or Tongva or Gabrieleno or Fernandeno or Tobikhar or Tonkawa or Ticanwatic or Tsikip or Appalousa or Opelousa or Tsitsistas or Tubatulabal or Tukabatchee or Tuscarora or Tomahittan or Kuskarawock or Tutelo or Tutero or Totteroy or Tutera or Yusan or Tututni or Umatilla or Umpqua or Waccamaw or Waxmaw or Wadatika or "Harney Valley Paiute" or Wailiki or Waluulapam or "Walla Walla" or Walpapi or Huipui or Wampanoag or Massasoit or Wanapum or Wappo or Washoe or Wichita or Willapa or Kwalhioqua or "Wi pukba" or "Verde Valley Yavapai" or Wintu or "Northern Wintun" or Wiyot or "Wee' at" or Weyet or Yakama or "Yamosopo Tuviwarai" or Yaqui or Yoeme or Yatasi or Yattasih or "Yavbe' " or "Yavapai" or "Ysleta del Sur" or Yojuane or Yokuts or Mariposa or Yuki or Yupighyt or "Yup'ik" or Yupik or Yurok or "Olekwo'l" or Zuni).mp. or ((Applegate or Delaware or Iowa or Ishak or Kaw or Kato or Spokane or Miami or Arkansas or Tali or Tunica or (Han not (China or Chinese)) or Pawnee or "Coeur D' Alene" or Piscataway or Ree or Tula) adj3 (reservation* or nation or people or peoples or population or man or men or woman or women or child* or youth* or elder or elders or communit* or tribe or tribes or tribal or Indian*)).mp. [United States]** | **20783** |
| **40** | **((south asia/ or exp India/ or Bangladesn/ or Bhutan/ or Nepal/ or Pakistan/ or Sri Lanka/ or (India or Bangladesh* or (Bhutan* not bhutanensis) or (Nepal* not nepalensis) or Pakistan* or "Sri Lanka*").mp.) and (indigenous people/ or ((Indigenous* or tribe or tribal or tribes or Rai) adj3 (population* or people* or person* or elder* or man or men or woman or women or child* or youth* or clan or clans or tribe or tribes or tribal or family or families or parent* or grandparent* or elder or elders or grandmother* or grandfather* or baby or babies or infant or infants or patient or patients or speakers or speaking or village* or communit*)).mp.)) or ("Adivasis" or "Adnamanese" or "Andaman" or "Baluch" or "Baluchis" or "Bodo" or "Boro" or "Boros" or "Bote" or "Brahuis" or "Chakmas" or "Chepang" or "Chhantyal" or "Damai" or "Dewan" or "Ghale" or "Gurkha" or "Gurung" or "Hayu" or "Hyolmo" or "Jarawa" or "Jirel" or "Jumma" or "Kalash" or "Khas" or "Kirati" or "Koinch" or "Kulung" or "Kusunda" or "Limbu" or "Lohorung" or "Magar" or "Makrani" or "Mangar" or "Marma" or "Miji" or "Mongar" or "Mro" or "Naga" or "Nepami" or "Newar" or "Nicobar" or "Onge" or "Rang" or "Raute" or "Sajolang" or "Santhal" or "Sentinelese" or "Sindhis" or "Sulemani" or "Sunuwar" or "Tamang" or "Thakali" or "Thangmi" or "Tharu" or "Tripura" or "Tumbahangphe" or "Wanniyala-Aetto" or "Yakkha" or "Yolmopa").mp. [Indian Sub-Continent]** | **8095** |
| **41** | **Indians, South American/ or (Abipon or Achuar or Achuagua or Akawaio or Amarizana or Andoque or Akawaio or Akuriyo or Anauya or Araona or Arawak or Ayamn or Aguaruna or Amahuaca or Amarakaeri or Andoa or Arabela or Arawak or Arhuaco or Ashaninca or Asheninca or Atsahuaca or Aymara or Ayoreo or Bakairi or "Baniva" or Barasana or Baniwa or "Baure" or Bororo or Cabiyari or Cacataibo or Caquinte or Cacua or Cahuarano or "Caiua" or "Camara Indians" or Camaracoto or Camsa or Canamari or Candoshi or Canela or Canichana or Capanahua or Carapana or Cariay or "Carib" or Carijona or Carutana or Cashibo or Cashinahua or Cawishana or Cavinena or Caxuiana or Cayuvava or Chontaquiro or Cocama or "Cubeo" or Curipaco or Chacobo or Chaima or (Chana not striatus) or Chapacura or Charrua or Chimila or Chitonahua or Chorote or Chipaya or Chiquitano or Chulupi or Carare or Coconuco or Cofan or Coreguaje or Coyaima or Chamacoco or Chamicuro or Chayahuita or Cocama or "Culina" or Culino or Cubeo or Cuiba or "Cuiva" or Cumanagoto or Curripaco or "Deni" or Desano or Embera or Guarani or Guajajara or "Guana" or Guanano or Guarayo or Guarayu or Guahibo or Guajiro or Guambiano or Guanano or Guayabero or Guarequena or Guinao or "Guana" or Gayon or Guahibo or Hixkaryana or Huachipairi or Huambisa or Huarayo or Lauanaua or Ikpeng or Ingariko or Irantxe or Itonama or Inapari or Iquito or Isconahua or Jumana or Japreria or Jirajara or Juruti or Jaqaru or Jebero or Kadiweu or Kaingang or Kamayura or Karaja or Karipuna or "Kariri" or Katukina or Kaxarari or Kayabi or Kayapo or "Kuikuro alapalo" or Kulina or "Kaiwa" or Kallawaya or "Kogui" or "Kuna" or Kaweskar or "Lule" or Macuna or Maipure or Mapuche or Mataco or Mocovi or Machinere or Machinerev or Machiguenga or Macushi or Macuna or "Madi" or Malayo or Mamainde or Manao or Mandauaca or Mandawaka or Mapidian or Mapuche or Mapidian or Maquiritare or Maquiritari or Maragua or Marawan or Mariate or Marubo or Mastanahua or Mataco or Matipuhy or "Matis" or "Matses" or Mawakua or Mawakwa or Maxakali or Mehinaku or Miranha or Moronawa or Munduruku or Movima or Muellama or Muinane or Mapoyo or "Mashco Piro" or Muniche or Nambikwara or Nocaman or Nuquini or Nomatsiguenga or Nanti or Ocaina or Omagua or Orejon or "Opon" or Pacahuara or "Paez" or Paicone or Palicur or Panare or "Pano" or "Paresi" or Paumari or "Pemon" or Pilaga or Puelche or Pauna or Pauserna or Piapoco or Piraha or Piratapuyo or Pisabo or Piaroa or "Pijao" or Piratapuyo or Paraujano or Pemon or Pemono or Piapoco or Puinave or Patamona or Poyanawa or Puinave or Puquina or Quechua or Quichua or Retuara or Resigaro or Reyesano or Sabanes or "Saliba" or Saluma or Sarave or Secoya or Selknam or "Sensi" or Shaninawa or Shapra or Sharanahua or Shebayo or Shiwiar or Shikiana or Sikiana or Siriono or Sinsiga or "Siona" or Suruwaha or Tacano or Tamanaco or Tiahuanaco or Tariano or Tehuelche or Tariano or Tatuyo or "Tembe" or "Terena" or Telembi or Ticuna or Ticuna or Tiriyo or Tiwanaku or Tiwanaku or "Torom" or "Totoro" or Tsimane or Tuberao or "Tucano" or Tunebo or Tuxinawa or Tuyuca or Uainuma or Urarina or Vilela or Waimaha or Waiampi or Waiwai or Wapishana or Waraiku or Warekena or Waura or Wayampi or Wayana or Wirina or Waimaha or Waunana or "Wiwa" or "Warao" or Wayuu or Witoto or Xavante or Xipaya or Xiriana or Xokleng or Yabaana or Yaminawa or Yaminahua or Yaruma or Yawalapiti or Yuracare or Yabarana or Yavitero or "Yine" or Yamana or Yaghan or Yucuna or Yurumangui or Yukpa or Yanesha or Yoranahua or Yagua or Yaminahua or Zaparo or Zamuco or "Trio Indians" or "More Indians" or "Bare Indians" or (("Inga" or "Maca" or "Leco" or "Mojo" or "Uro" or "Maco" or Lengua or "Toba" or "Zoe" or "Ona" or "Catio" or "Passe" or "Bari" or "Awa" or "Bora" or "Bara" or "Remo" or "Pano" or "Sape") adj3 (Indians or Indian or Indigenous or Amerindian* or Aborigin* or people or peoples or elder or elders or grandmother* or grandfather* or parent* or women or men or woman or man or child* or youth or youths or baby or babies or tribe or tribes or tribal or shaman* or native or patient or patients))).mp. or ((Indian* or Amerindian, or Aboriginal* or Indigenas or Indigenous or tribe or tribes or tribal).mp. and (exp South America/ or "South America*".mp. or Argentin*.mp. or Bolivia*.mp. or Brazil*.mp. or Chile*.mp. or Colombia*.mp. or Ecuador*.mp. or "French Guiana".mp. or Guyana*.mp. or Paraguay*.mp. or Peru.mp. or Peruvian.mp. or Suriname.mp. or Uruguay*.mp. or Venezuela*.mp. or "Amazon Region".mp. or Amazonia.mp. or Andes.mp. or Andean.mp.)) [South America]** | **9437** |
| **42** | **((Ainus or Ainu or Aleuts or Alyutors or Chukchis or Chuvans or Dolgans or Enets or Entsy or Yupik or Yup'ik or Yuit or Yupigyt or Chaplino or Naukan or Itelmens or Kamchadals or Kereks or "Komi" or Koryaks or Nenets or Nentsy or Nganasans or Tavgi or Sami or Veps or Yukaghirs or Chulyms or Evenks or Tungus or Evens or "Kets" or Khantys or Mansi or Vguls or Selkups or Teleuts or Nanais or Nanaitsy or Negidal or Nivikh or Oroch or orok or Taz or udege or ulch or Kumadins or Chelkans or Shorians or Soyots or Telengits or Tofalars or Tugalars or "Tufans Todzhins" or Laks or Tabasarans or Turuls or Aguls or Tsakhurs or Kumyks or Nogais or "Andis" or Akhvakh or Archins or Bagvalals or Bezhta or Botlikhs or Chamalals or Godoberi or Hinukh or Hunzibs or Khwarshi or Karata or Tindis or Tsez or Abazin or Besermyan or Izhorians or Karelians or Nagaybaks or Setos or Shapsugs or Quratay).mp. or (exp indigenous people/ or (Indigenous adj3 (population* or people* or person* or elder* or man or men or woman or women or child* or youth* or clan or clans or tribe or tribes or tribal or family or families or parent* or grandparent* or elder or elders or grandmother* or grandfather* or baby or babies or infant or infants or patient or patients or speakers or speaking or village* or communit*)).mp.)) and (Russia*.mp. or exp Russia/)** | **502** |
| **43** | **((greenland/ or Greenland*.mp. or "Kalaallit Nunaat".mp. or Nuuk.mp. or Sisimiut.mp. or Ilulissat.mp. or Qaqortoq Aasiaat.mp. or Maniitsoq.mp. or Tasiilaq.mp. or Uummannaq.mp. or Narsaq.mp. or Paamiut.mp. or Nanortalik.mp. or Upernavik.mp. or Qasigiannguit.mp.) and (Indigenous Peoples/ or Inuit.mp. or Inuk.mp. or ((Indigen* or Aborigin*) adj3 (population* or people or peoples or person or persons or elder or elders or man or men or woman or women or child* or youth* or clan or clans or tribe or tribes or tribal or family or families or parent* or grandparent* or elder or elders or grandmother* or grandfather* or baby or babies or infant or infants or patient or patients or speakers or speaking or village* or communit*)).mp.)) or (Greenlandic or Kalaallit or Kalaallisut or Tunumiit or Inughuit or Avanersuarmiut).mp. [Greenland]** | **481** |
| **44** | **((exp South East Asia/ or ("Bali" or "Borneo" or "Brunei" or Cambodia or "Flores Island" or "Indonesia" or Java or Komodo or "Laos" or "Lombok" or "Malasia" or "Malay Peninsula" or "Myanmar" or "Mekong Valley" or "New Guinea" or "Philippeans" or "Sabah" or "Singapore" or "Sumatra" or "Sumba" or "Sumbawa" or "Thailand" or "Timor" or "Vietnam").mp.) and (Indigenous Peoples/ or ((Indigen* or aborig*) adj3 (population* or people or peoples or person or persons or elder or elders or man or men or woman or women or child* or youth* or clan or clans or tribe or tribes or tribal or family or families or parent* or grandparent* or elder or elders or grandmother* or grandfather* or baby or babies or infant or infants or patient or patients or speakers or speaking or village* or communit*)).mp.)) or ("Abui" or "Aeta" or "Alfur" or "Ati People" or "Bahau" or "Bali Aga" or "Bali Mula" or "Baliaga" or "Balinese" or "Basap" or "Bataq" or "Batek" or "Bateq" or "Batin" or "Blaan" or "Brao" or "Bru" or "Bru-Van Kieu" or "Bugkalot" or "Bumiputera" or "Bunong" or "Chams" or "Champa" or "Cheq Wong" or "Dasun" or "Dayak" or "Deli Malay" or "Filipina*" or "Filiopino*" or "Gaddang" or "Ibaloi" or "Iban" or "Idan" or "Igorot" or "Jahai" or "Jehai" or "Jakun" or "Javanese Kshatriya" or "Kangeanese" or " Kagayanen" or "Kanayatn" or "Katu" or "Kayan" or "Kavet" or "Kenyah" or "Khmer Krom" or "Kimaragang" or "Klemantan" or "Kreung" or "Kuijau" or "Kuy" or "Kwijau" or "Lampung*" or "Lanoh" or "Lawangan" or "Luangan" or "Lumad" or "Dayak" or "Loloan Malay*" or "Mah Meri" or "Malay Singaporean*" or "Malbog" or "Maluku" or "Mangka'ak" or "Mangyan" or "Maniq" or "Maragang" or "Ma'anyan" or "Minokok" or "Moluccas" or Montagnard or "Murut" or "Naga" or "Negrito" or "New Guinea" or "Ngaju" or "Orang Hulu" or "Orang Kanaq" or "Orang Kuala" or "Orang Seletar" or "Orang Ulu" or "Ot Danum" or "Palawano" or "Palembang*" or " Panay-Bukidnon" or "Pan-ayanon" or "Papua" or "Phnong" or "Punong" or "Pear People" or "Por People" or "Punan" or "Rumanau" or "Sasak" or "Semai" or "Semang" or "Semaq Beri" or "Semelai" or "Senoi" or "Simeulue" or "Suludnon" or "Sunda Island*" or "Tabanwa" or "Takua" or "Tambanuo" or " Tombonuo" or "Taron" or "Temiar*" or "Temoq" or "Temuan" or "Tidung" or "Trone" or "Taaw't Bato").mp.** | **19104** |
| **45** | **(indigenous people/ or ((Indigen* or aborig*) adj3 (population* or people or peoples or person or persons or elder or elders or man or men or woman or women or child* or youth* or clan or clans or tribe or tribes or tribal or family or families or parent* or grandparent* or elder or elders or grandmother* or grandfather* or baby or babies or infant or infants or patient or patients or speakers or speaking or village* or communit*)).mp.) and (pacific islands/ or (Micronesia or "Caroline Island" or Guam or Kiribati or "Mariana Island*" or Nauru or Palau or Melanesia or "East Nusa Tenggara" or Fiji or Fijian or "Loyalty Island*" or "Maluku Islands" or Moluccas or "New Caledonia*" or "New Guinea" or Papua or Rotuma or "Solomon Island*" or Vanuatu or Polynesia or "Austral Island*" or "Cook Island*" or "Chatham Islands" or "Easter Island*" or Hawaii* or "Marquesas Island" or "Norfolk Island" or Samoa or Tahiti or Tokelau or Tonga or Tuvalu or Tuamotu or Kermadec or Banabans or "Native Hawaiian*" or Guamanian or Chamarro* or Chuukese or I-Kiribati or Kosraean* or Maohi or Marshallese or Moriori or Mortlockese or Mwokilese or Namonuito or Nauruans or Ngatikese or Niuean or Paafang or Paluans or Pingelapese or Pohnpeian* or Pollapese or Puluwat or Rapanui or Rotuman or Satwalese or Samoan*or Sonsorolese or Tahitian* or Tobian or Tongan or "Torres Strait Islander*" or Tuvaluan or Ulithian or Woleian or Yapese or Polynesian* or Micronesian* or Melanesian*).mp.)** | **1990** |
| **46** | **((exp Indigenous Peoples/ or ((Indige* or aborig* or nomadic or seminomad* or "semi nomad*") adj3 ("people" or "peoples" or "elder" or "elders" or "grandmother*" or "grandfather*" or "parent*" or "women" or "men" or "woman" or "man" or "child*" or "youth" or "youths" or "baby" or "babies" or "tribe" or "tribes" or "tribal" or "shaman*" or "native" or "patient*")).mp.) and (("Middle East" or "Afghanistan" or "Bahrain" or "Iran" or "Iraq" or "Israel" or ("Jordan" not "Jordan's Principle") or "Kuwait" or "Lebanon" or "Oman" or "Palestine" or "Qatar" or "Saudi Arabia").mp. or middle east/ or afghanistan/ or bahrain/ or iran/ or iraq/ or israel/ or jordan/ or kuwait/ or lebanon/ or oman/ or qatar/ or saudi arabia/ or syria/ or turkey/ or united arab emirates/ or yemen/)) or ("Bedouin*" or "Jahalin" or "al-Kaabneh" or "al-Azazmeh" or "al-Ramadin" or "al-Rshaida").mp.** | **912** |
| **47** | **((Mexico/ or ((Mexico not "New Mexico") or Mexican or Aguascalientes or "Baja California" or Campeche or Chiapas or (Chihuahua not (dog or dogs or pet or pets)) or Coahuila or Colima or Durango or Guanajuato or Guerrero or Hidalgo or Jalisco or Michoacan or Morelos or Nayarit or "Nuevo Leon" or Oaxaca or Puebla or Queretaro or "Quintana Roo" or "San Luis Potosi" or Sinaloa or Sonora or (Tabasco not (sauce* or flavo*)) or Tamaulipas or Tlaxcala or Veracruz or Yucatan or Zatecas).mp.) and (exp indigenous people/ or (Mesoamerindian* or Indigen* or aborig* or "first people*" or "indos mexicano" or "original people*" or "pueblos indigenas").mp.)) or (Aguacatec or Akwa'ala or Abxubal or Ayuukja'ay or "Batzil k'op" or Binizaa or "Chichimeca Jonaz" or Chinantec or Chocho or "Ch ol" or Chontal or "Chuj" or Cochimi or Comcaac or Hamasipini or Harijio or "Ha shuta enima" or "Hach t'an" or Huastecor Hnahnu or Hnatho or Ixcatec or Ixil or Jacaltec or K'akchikel or K'anjobal or Kanjobal or Kaqchikel or Kechi or K'iche or Kikapooa or Kikapu or Kiliwa or "Ko'lew" or "K'op o winik atel" or Kumiai or Lacandon or Laymon or Makurawe or Maya or Maya'wiinik or Mazahua or Mazatec or Me'phaa or Mexicanero or Mexikatlajtolli or "Mixe" or Mixtec or Motocintleco or Mti'pa or Nahuas or Ocuiltec or Otomi or (Oaxaca not Oaxaca-Blinder) or "Pame" or Papago or Tlahuica or Paipai or "Pima Bajo" or Purepecha or P'urhepecha or Qatok or (Quiche not Guatemala) or Q'iche or Raramuri or "Runixa ngiigua" or ("Seri" not "Seri 82") or "Slijuala sihanuk" or Tacuate or Tarahumara or Teenek or Tepehua or Ti'pai or Tlapanec or "Tohono O'odham" or Totonac or Tachiwin or "Tsa jujmi" or Tzotzil or "Tu'un savi" or Tzeltal or "Uza" or "Winik" or Xigue or Yucatec or Zapotec).ti,ab.** | **2494** |
| **48** | **or/33-47** | **77636** |
| **49** | **32 and 48** | **476** |
| **50** | **limit 49 to up=20230701-20240214** | **29** |

**EBSCO CINAHL Searched August 29, 2022**

**Search Mode: Find all my search terms**

**EBSCO CINAHL**

**Search for all my terms**

| **#** | **Query** | **Results** |
| --- | --- | --- |
| **S1** | **(MH "Urinary Incontinence+") OR (MH "Enuresis, Nocturnal") OR (MH "Stress Incontinence")** | **13,100** |
| **S2** | **(MH "Fecal Incontinence")** | **3,359** |
| **S3** | **(MH "Pelvic Organ Prolapse+") OR (MH "Cystocele") OR (MH "Rectal Prolapse") OR (MH "Rectocele") OR (MH "Uterine Prolapse") OR (MH "Vaginal Vault Prolapse")** | **3,677** |
| **S4** | **(MH "Overactive Bladder")** | **2,180** |
| **S5** | **(MH "Urinary Retention")** | **1,755** |
| **S6** | **(MH "Constipation+") OR (MH "Opioid-Induced Constipation")** | **7,269** |
| **S7** | **(MH "Pelvic Floor Disorders") OR (MH "Pelvic Floor Muscles")** | **3,365** |
| **S8** | **(MH "Pelvic Pain")** | **3,495** |
| **S9** | **(MH "Dyspareunia")** | **1,213** |
| **S10** | **(MH "Vulvodynia")** | **83** |
| **S11** | **(MH "Vaginismus")** | **83** |
| **S12** | **(MH "Interstitial Cystitis")** | **733** |
| **S13** | **(MH "Adenomyosis")** | **447** |
| **S14** | **(MH "Surgery, Gynecologic+")** | **20,287** |
| **S15** | **(MH "Colectomy+")** | **3,479** |
| **S16** | **(MH "Rectal Neoplasms+")** | **7,590** |
| **S17** | **(MH "Endometrial Neoplasms") OR (MH "Genital Neoplasms, Female+") OR (MH "Urogenital Neoplasms+") OR (MH "Uterine Neoplasms+") OR (MH "Cervix Neoplasms+")** | **108,592** |
| **S18** | **(MH "Ovarian Neoplasms+")** | **15,863** |
| **S19** | **(MH "Genital Neoplasms, Male+") OR (MH "Penile Neoplasms") OR (MH "Prostatic Neoplasms+") OR (MH "Prostatic Neoplasms, Castration-Resistant") OR (MH "Testicular Neoplasms")** | **39,811** |
| S20 | (MH "Prostatectomy+") | 7,077 |
| **S21** | **((gyn?eologic* or rectal or rectum or genital or prostat* or unterine or uterus or ovarian or endometrial) N3 (surgery or surgical))** | **18,523** |
| **S22** | **(MH "Postoperative Complications+") or (postoperative or "post operative" or "post surg*"** | **245,909** |
| **S23** | **((MH "Postoperative Complications+")) AND (S14 OR S15 OR S16 OR S17 OR S18 OR S19 OR S20 OR S21)** | **8,207** |
| **S24** | **(MH "Bladder, Neurogenic")** | **1,284** |
| **S25** | **(MH "Sexual Dysfunction, Male+") OR (MH "Sexual Dysfunction, Female+") OR (MH "Sexual Dysfunction, Psychological")** | **12,351** |
| **S26** | **(MH "Enuresis, Nocturnal")** | **319** |
| **S27** | **(MH "Hirschsprung Disease+")** | **824** |
| **S28** | **(MH "Gender Affirmation Surgery")** | **411** |
| **S29** | **(MH "Lichen Sclerosus et Atrophicus")** | **222** |
| **S30** | **(MH "Bladder Fistula+") OR (MH "Vaginal Fistula+") OR (MH "Urinary Fistula+")** | **1,160** |
| **S31** | **(MH "Pelvic Floor Muscles") OR (MH "Pelvic Floor Disorders")** | **3,365** |
| **S32** | **(S1 OR S2 OR S3 OR S4 OR S5 OR S6 OR S7 OR S8 OR S9 OR S10 OR S11 OR S12 OR S13 OR S23 OR S24 OR S25 OR S26 OR S27 OR S28 OR S29 OR S30 OR S31)** | **54,848** |
| **S33** | **(prolaps* N3 ("pelvic organ" or uterine or uterus or bladder or rectal or rectum or urethra* or apical))** | **4,608** |
| **S34** | **incontinence** | **23,249** |
| **S35** | **uterocele or cystocele or rectocele or enterocele or urethrocele or sigmoidocele** | **538** |
| **S36** | **bladder* N3 (overactive or urgency or urgent or frequency or heistency or retention)** | **3,199** |
| **S37** | **bowel N3 (frequency or urgency)** | **416** |
| **S38** | **constipation** | **12,047** |
| **S39** | **encopresis** | **171** |
| **S40** | **(dyssynergia N3 (bowel* or bladder*))** | **18** |
| **S41** | **"vaginal wind"** | **4** |
| **S42** | **"pelvic girdle pain"** | **344** |
| **S43** | **dyspareunia or vulvodynia or vaginismus or vestibulodynia or "interstitial cystitis" or "painful bladder syndrome" or "proctalgia fugax" or prostatitis or endometriosis or adenomyosis or "pudendal neuralgia" or anismus** | **14,011** |
| **S44** | **neurogenic bladder*** | **1,964** |
| **S45** | **("sexual dysfunction" or "erectile dysfunction" or "persistent genital arousal" or impotence)** | **13,717** |
| **S46** | **("bed wett*" or bedwett* or encopresis or "hirshprung* disease")** | **463** |
| **S47** | **("gender confirmation surg*" or "sex reassignment surg*")** | **149** |
| **S48** | **"lichen sclerosis"** | **39** |
| **S49** | **nocturia** | **1,049** |
| **S50** | **((fistula* or fistulae) N3 (bladder* or urinary or vagina* or rectovaginal* or vesicovaginal*))** | **1,633** |
| **S51** | **"levator ani avulsion"** | **14** |
| **S52** | **"Lower urinary tract symptom*" or dysuria or "LUTS"** | **3,355** |
| **S53** | **"voiding dysfunction"** | **463** |
| **S54** | **(S32 OR S33 OR S34 OR S35 OR S36 OR S37 OR S38 OR S39 OR S40 OR S41 OR S42 OR S43 OR S44 OR S45 OR S46 OR S47 OR S48 OR S49 OR S50 OR S51 OR S52 OR S53)** | **83,483** |
| **S55** | **(MH "Aboriginal Canadians+") OR (MH "First Nations of Canada") OR ( ( (MH "Eskimos") OR (MH "Native Americans") OR (MH "Indigenous Peoples+") or (MH "Health Services, Indigenous") or (MH "Indigenous Health") or (MH “Ethnopharmacology”) or Athapaskan or Saulteaux or Wakashan or Cree or Dene or Inuit or Innu or Inuk or Inuvialuit* or Haida or Ktunaxa or Tsimshian or Gitsxan or “Nisga'a” or Haisla or Heiltsuk or Oweenkeno or “Kwakwaka'wakw” or “Nuu chah nulth” or “Tsilhqot'in” or Dakelh or “Wet'suwet'en” or Sekani or “Dunne-za” or Dene or Tahltan or Kaska or Tagish or Tutchone or Nuxalk or Salish or “Stl'atlimc” or “Nlaka'pamux” or Okanagan or “Sec wepmc” or Tlingit or Anishinaabe or Blackfoot or Nakoda or Tasttine or “Tsuu T'inia” or “Gwich'in” or Han or Tagish or Tutchone or Algonquin or Nipissing or Ojibwa or Potawatomi or Innu or Maliseet or “Mi'kmaq” or Micmac or “Mic mac” or Passamaquoddy or Haudenosaunee or Cayuga or Mohawk or Oneida or Onodaga or Seneca or Tuscarora or Wyandot or Aboriginal* or Indigenous* or Metis or “red road” or "on reserve" or “off reserve” or “First Nation” or “First Nations” or Amerindian or (urban N3 (Indian* or Native* or Aboriginal*)) or ethnomedicine or “country food*” or “residential school*” or ((MH “Medicine, Traditional”) or "traditional medicine*" ) not Chinese ) or ( MH“Shamanism”) or shaman* or “traditional heal*” or “traditional food*” or “medicine man” or “medicine woman” or autochtone* or (Native* N1 (American* or man or men or women or woman or boy* or girl* or adolescent* or youth or youths or person* or adult or people* or Indian* or Nation or Nations or tribe* or tribal or band or bands or elder or elders or patient*)) ) ) AND ( ( (MH “Canada+”) or (Canad* or “British Columbia” or “Colombie Britannique” or Alberta or Saskatchewan or Manitoba or Ontario or Quebec or “Nova Scotia” or “New Brunswick” or Newfoundland or Labrador or “Prince Edward Island” or “Yukon Territory” or NWT or “Northwest Territories” or Nunavut or Nunavik or Nunatsiavut or NunatuKavut)) )** | **5,540** |
| **S56** | **((MH "Australia+") or (Australia* or "Northern Territory" or "Australian Capital Territory" or Tasmania or "New South Wales" or "Victoria" or "Queensland")) and ( Aborigin* or Indigen*)) or (MH "First Nations of Australia") OR (MH "Aboriginal Australians") OR (MH "Torres Strait Islanders") or "Torres Strait Islander*"** | **9,575** |
| **S57** | **(MH Maori) or ("Maori" or "tangata whenua") or (( (MH New Zealand) or (New Zealand or Aukland).mp.) and ( ("Aborig*" or "Indig*")))** | **3,285** |
| **S58** | **("Acatec" OR "Aguacateco" OR "Amuzgo" OR " Bokota " OR "Boruca " OR "Bribri" OR "Bri Bri" OR "Buglere" OR "Cabecar" OR " Cakchiquel " OR "Changuena" OR "Chatino" OR "Chiapanec" OR "Chicomuceltec" OR "Chinantee" OR "Chocho" OR "Cholti" OR "Ch'olti" OR "Ch'olti'an" OR "Chontal" OR "Chorotega " OR "Chorti " OR "Chuj " OR "Chumbia " OR "Corobici" OR ("Cueva" not Spain) OR "Cuicatec" OR "Cuitlatee" OR "Cuytec" OR "Dorasque " OR "Embera" OR "Garifuna " OR "Guatuso" OR "Guaymi" OR "Guaymis" OR "Guetar " OR "Huastec " OR " Huave" OR "Huetar" OR "Itzaj" OR "Ixil " OR "Jacalteco " OR "Jonaz " OR "Kanjobal" OR "Kekchi" OR "Kuna" OR "Maleku " OR "Mangue " OR "Matambu" OR "Matlatzinca" OR "Mazahua " OR "Motozintlec" OR "Mayan" OR "Mayangna " OR " Miskito " OR " Mixtec" OR "Mopan" OR "Nahua" OR "Nahuatl" OR "Ngabe" OR "Otomi" OR "Pantec" OR "Paya" OR "Popoloca" OR "Popoloc" OR "Poqomam" OR " Poqomchi" OR "Q'eqchi'" OR "Quiche" OR "Quitirrisi" OR "Sacapulteco" OR "Sipacapense" OR "Subtiaba" OR "Tacaneco" OR "Tarasco" OR "Tamaulipec" OR "Tamazultec" OR "Tecoxquin" OR "Tectiteco" OR "Tecual" OR "Tecuexe" OR "Tepehura" OR "Tepuztecor" OR "Teribe" OR "Terraba" OR "Totonac " OR "Trique" OR " Tzeltal " OR "Tzotzil " OR " Tzutujil " OR "Ulwa " OR "Uspantec*" OR "Uspanteko" OR "Voto " OR "Xinca" OR "Waunana " OR "Wounaan" OR "Yucatec " OR "Zapotec " OR "Zoque") OR (((MH "Central America") OR "(MH "Belize") OR (MH "Costa Rica") OR (MH "El Salvador") OR (MH "Guatemala") OR (MH "Honduras") OR (MH "Nicaragua") OR (MH "Panama") OR (MH "Panama Canal Zone") OR "Belize" OR "or Belizean*" OR ""Costa Rica*"" OR""El Salvador" OR " Hondura*" OR "Nicaragua*" OR "Panama*" OR "Guatemala*" OR "Achi" OR "Maya*" OR "Mixe" OR "Pame" OR "Pipil" OR "Pech" OR "Chol" OR "Ch'olan" OR "Cora" OR "Cuna" OR " Mam" OR "Rama) N5 (Indian" OR "Indians" OR "Amerindian*" OR "Amerindio*" OR "Aboriginal*" OR "Indigenous" OR "Indigena*" OR "Aborigen*" OR "Mestizo" OR "tribe" OR "tribes" OR "tribal" OR "traditional medicine*" OR "shaman*)) NOT ("Mexico" OR (MH Mexico)** | **61,136** |
| **S59** | **(Greenland/ or Greenland* or "Kalaallit Nunaat" or "Nuuk" or "Sisimiut" or "Ilulissat" or "Qaqortoq Aasiaat" or "Maniitsoq" or "Tasiilaq" or "Uummannaq" or "Narsaq" or "Paamiut" or "Nanortalik" or "Upernavik" or "Qasigiannguit") and (Indigenous Peoples/ or "Inuit" or "Inuk" or ((Indigen* or Aborigin*) adj3 (population* or people or peoples or person or persons or elder or elders or man or men or woman or women or child* or youth* or clan or clans or tribe or tribes or tribal or family or families or parent* or grandparent* or elder or elders or grandmother* or grandfather* or baby or babies or infant or infants or patient or patients or speakers or speaking or village* or communit*)).mp.) or (Greenlandic or "Kalaallit" or "Kalaallisut" or "Tunumiit" or "Inughuit" or "Avanersuarmiut").mp.** | **119** |
| **S60** | **((MH "Bangladesh") OR (MH "Bhutan") OR (MH "India") OR (MH "Nepal") OR (MH "Pakistan") OR (MH "Sri Lanka") or (India or Bangladesh* or (Bhutan* not bhutanensis) or (Nepal* not nepalensis) or Pakistan* or "Sri Lanka*")) AND ((MH "Indigenous Peoples+") or ((Indigenous* or aborigin* or tribe or tribal or tribes) N3 (population* or people* or person* or elder* or man or men or woman or women or child* or youth* or clan or clans or tribe or tribes or tribal or family or families or parent* or grandparent* or elder or elders or grandmother* or grandfather* or baby or babies or infant or infants or patient or patients or speakers or speaking or village* or communit*))) OR ("Adivasis" OR "Adnamanese" OR "Andaman" OR "Baluch" OR "Baluchis" OR "Bodo" OR "Boro" OR "Boros" OR "Bote" OR "Brahuis" OR "Chakmas" OR "Chepang" OR "Chhantyal" OR "Damai" OR "Dewan" OR "Ghale" OR "Gurkha" OR "Gurung" OR "Hayu" OR "Hyolmo" OR "Jarawa" OR "Jirel" OR "Jumma" OR "Kalash" OR "Khas" OR "Kirati" OR "Koinch" OR "Kulung" OR "Kusunda" OR "Limbu" OR "Lohorung" OR "Magar" OR "Makrani" OR "Mangar" OR "Marma" OR "Miji" OR "Mongar" OR "Mro" OR "Naga" OR "Nepami" OR "Newar" OR "Nicobar" OR "Onge" OR "Rai" OR "Rang" OR "Raute" OR "Sajolang" OR "Santhal" OR "Sentinelese" OR "Sindhis" OR "Sulemani" OR "Sunuwar" OR "Tamang" OR "Thakali" OR "Thangmi" OR "Tharu" OR "Tripura" OR "Tumbahangphe" OR "Wanniyala-Aetto" OR "Yakkha" OR "Yolmopa")** | **8,667** |
| **S61** | **(((MH Mexico+) or ((Mexico not "New Mexico") or Mexican or Aguascalientes or "Baja California" or "Campeche" OR "Chiapas" OR ("Chihuahua" not ("dog" OR "dogs" OR "pet" OR "pets")) OR "Coahuila" OR "Colima" OR "Durango" OR "Guanajuato" OR "Guerrero" OR "Hidalgo" OR "Jalisco" OR "Michoacan" OR "Morelos" OR "Nayarit" OR "Nuevo Leon" OR "Oaxaca" OR "Puebla" OR "Queretaro" OR "Quintana Roo" OR "San Luis Potosi" OR "Sinaloa" OR "Sonora" OR ("Tabasco" not ("sauce*" OR "flavo*")) OR "Tamaulipas" OR "Tlaxcala" OR "Veracruz" OR "Yucatan" OR "Zatecas)) and ((MH "Indigenous Peoples+") OR ("Mesoamerindian*" OR "Indigen*" OR "aborig*" OR "first people*" OR "indos mexicano" OR "original people*" OR "pueblos indigenas"))) OR "(Aguacatec" OR "Akwa'ala" OR "Abxubal" OR "Ayuukja'ay" OR "Batzil k'op" OR "Binizaa" OR "Chichimeca Jonaz" OR "Chinantec" OR "Chocho" OR "Ch ol" OR "Chontal" OR "Chuj" OR "Cochimi" OR "Comcaac" OR "Hamasipini" OR "Harijio" OR "Ha shuta enima" OR "Hach t'an" OR "Huastecor Hnahnu" OR "Hnatho" OR "Ixcatec" OR "Ixil" OR "Jacaltec" OR "K'akchikel" OR "K'anjobal" OR "Kanjobal" OR "Kaqchikel" OR "Kechi" OR "K'iche" OR "Kikapooa" OR "Kikapu" OR "Kiliwa" OR "Ko'lew" OR "K'op o winik atel" OR "Kumiai" OR "Lacandon" OR "Laymon" OR "Makurawe" OR "Maya" OR "Maya'wiinik" OR "Mazahua" OR "Mazatec" OR "Me'phaa" OR "Mexicanero" OR "Mexikatlajtolli" OR "Mixe" OR "Mixtec" OR "Motocintleco" OR "Mti'pa" OR "Nahuas" OR "Ocuiltec" OR "Otomi" OR ("Oaxaca" not Oaxaca-Blinder) OR "Pame" OR "Papago" OR "Tlahuica" OR "Paipai" OR "Pima Bajo" OR "Purepecha" OR "P'urhepecha" OR "Qatok" OR ("Quiche" not "Guatemala") OR "Q'iche" OR "Raramuri" OR "Runixa ngiigua" OR ("Seri" not "Seri 82") OR "Slijuala sihanuk" OR "Tacuate" OR "Tarahumara" OR "Teenek" OR "Tepehua" OR "Ti'pai" OR "Tlapanec" OR "Tohono O'odham" OR "Totonac" OR "Tachiwin" OR "Tsa jujmi" OR "Tzotzil" OR "Tu'un savi" OR "Tzeltal" OR "Uza" OR "Winik" OR "Xigue" OR "Yucatec" OR "Zapotec")** | **25,592** |
| **S62** | **( ((MH "Indigenous Peoples+" or ((Indige* or aborig* or nomadic or seminomad* or "semi nomad*") N3 ("people" OR "peoples" OR "elder" OR "elders" OR "grandmother*" OR "grandfather*" OR "parent*" OR "women" OR "men" OR "woman" OR "man" OR "child*" OR "youth" OR "youths" OR "baby" OR "babies" OR "tribe" OR "tribes" OR "tribal" OR "shaman*" OR "native" OR "patient*"))) ) AND ( ((MH "Middle East") OR (MH "Afghanistan") OR (MH "Bahrain") OR (MH "Iran") OR (MH "Iraq") OR (MH "Israel") OR (MH "Jordan") OR (MH "Kuwait") OR (MH "Lebanon") OR (MH "Oman") OR (MH "Qatar") OR (MH "Saudi Arabia") OR (MH "Syria") OR (MH "Turkey") OR (MH "United Arab Emirates") OR (MH "Yemen") or "Middle East" or "Afghanistan" or "Bahrain" or "Iran" or "Iraq" or "Israel" or ("Jordan" NOT "Jordan's Principle") or "Kuwait" or "Lebanon" or "Oman" or "Palestine" or "Qatar" or "Saudi Arabia")) )** | **226** |
| **S63** | **TI("Bedouin*" OR "Jahalin" OR "al-Kaabneh" OR "al-Azazmeh" OR "al-Ramadin" OR "al-Rshaida") OR AB"Bedouin*" OR "Jahalin" OR "al-Kaabneh" OR "al-Azazmeh" OR "al-Ramadin" OR "al-Rshaida")** | **333** |
| **S64** | **TI("Saami" or "Sampi" or ("Sami" not "Ulus") or "Samis" or "Southernsami*" or "Umesami*" or "Pitesami*" or "Lulesami*" or "Northernsami*" or "Enaresami*" or "Kolasami*" or "Lapp" or "Lapps" or "Lappish" or "Lappland" or ("Lapland*" not longspur) or "Lappalainen*" or "Saamelainen*" or "reindeer herd*" or "reindeer culture*" or "reindeer pastoral*" or "Lappbys" or "Samebys" or "reinbeitesdistrikt" or "paliskunta" or "siida") or AB ("Saami" or "Sampi" or ("Sami" not "Ulus") or "Samis" or "Southernsami*" or "Umesami*" or "Pitesami*" or "Lulesami*" or "Northernsami*" or "Enaresami*" or "Kolasami*" or "Lapp" or "Lapps" or "Lappish" or "Lappland" or ("Lapland*" not longspur) or "Lappalainen*" or "Saamelainen*" or "reindeer herd*" or "reindeer culture*" or "reindeer pastoral*" or "Lappbys" or "Samebys" or "reinbeitesdistrikt" or "paliskunta" or "siida") or ((("Fennoscandia" or "Finnmark" or "Scandinavia" or "Nordic" or "Sweden" or "Norway" or "Finland" or "Swedish" or "Finnish" or "Norwegian" or "Norge" or "Svensk*" or "Suomi" or "Barents Region" or (Kola not (garcinia or gotu)) or "Arctic Europe*" or "Polar Europe*" or "North* Europ*") or (MH "Scandinavia") OR (MH "Finland") OR (MH "Norway") OR (MH "Sweden") ) and ((traditional N3 (food* or heal* or medicine* or shaman*)) or ((Indigenous* or "aborig*") N3 (people* or person* or mother* or father* or parent* or grandparent* or grandmother* or grandfather* or child* or boy or boys or girl* or youth* or healer* or patient or patients or famil* or herder*or elder or elders or village* or communit*))))** | **509** |
| **S65** | **(MH "Indigenous Peoples+")** | **23,358** |
| **S66** | **(((MH "Indigenous Peoples+") or ((Indigen* or aborig*) N3 ("population*" or people or peoples or person or persons or elder or elders or man or men or woman or women or child* or youth* or clan or clans or tribe or tribes or tribal or family or families or parent* or grandparent* or elder or elders or "grandmother*" or "grandfather*" or "baby" or "babies" or "infant" or "infants" or "patient" or "patients" or speakers or "speaking" or "village*" or "communit*"))) and (((MH "Pacific Islands+") OR (MH "Melanesia+") OR (MH "Micronesia+") OR (MH "Polynesia+") ) or (Micronesia or "Caroline Island" or "Guam" or "Kiribati" or "Mariana Island*" or "Nauru" or "Palau" or Melanesia or "East Nusa Tenggara" or "Fiji" or "Fijian" or "Loyalty Island*" or "Maluku Islands" or Moluccas or "New Caledonia*" or "New Guinea" or "Papua" or "Rotuma" or "Solomon Island*" or "Vanuatu" or Polynesia or "Austral Island*" or "Cook Island*" or "Chatham Islands" or "Easter Island*" or "Hawaii*" or "Marquesas Island" or "Norfolk Island" or "Samoa" or "Tahiti" or "Tokelau" or "Tonga" or "Tuvalu" or "Tuamotu" or "Kermadec" or "Banabans" or "Native Hawaiian*" or "Guamanian" or "Chamarro*" or "Chuukese" or "I-Kiribati" or "Kosraean*" or "Maohi" or "Marshallese" or "Moriori" or "Mortlockese" or "Mwokilese" or "Namonuito" or "Nauruans" or "Ngatikese" or "Niuean" or "Paafang" or "Paluans" or "Pingelapese" or "Pohnpeian*" or "Pollapese" or "Puluwat" or "Rapanui" or "Rotuman" or "Satwalese" or "Samoan*" or "Sonsorolese" or "Tahitian*" or "Tobian" or "Tongan" or "Torres Strait Islander*" or "Tuvaluan" or "Ulithian" or "Woleian" or "Yapese" or "Polynesian*" or "Micronesian*" or "Melanesian*")) )** | **4,325** |
| **S67** | **TI(("Ainus" OR "Ainu" OR "Aleuts" OR "Alyutors" OR "Chukchis" OR "Chuvans" OR "Dolgans" OR "Enets" OR "Entsy" OR "Yupik" OR "Yup'ik" OR "Yuit" OR "Yupigyt" OR "Chaplino" OR "Naukan" OR "Itelmens" OR "Kamchadals" OR "Kereks" OR "Komi" OR "Koryaks" OR "Nenets" OR "Nentsy" OR "Nganasans" OR "Tavgi" OR "Sami" OR "Veps" OR "Yukaghirs" OR "Chulyms" OR "Evenks" OR "Tungus" OR "Evens" OR "Kets" OR "Khantys" OR "Mansi" OR "Vguls" OR "Selkups" OR "Teleuts" OR "Nanais" OR "Nanaitsy" OR "Negidal" OR "Nivikh" OR "Oroch" OR "Orok" OR "Taz" OR "Udege" OR "Ulch" OR "Kumadins" OR "Chelkans" OR "Shorians" OR "Soyots" OR "Telengits" OR "Tofalars" OR "Tugalars" OR "Tufans" OR "Todzhins" OR "Laks" OR "Tabasarans" OR "Turuls" OR "Aguls" OR "Tsakhurs" OR "Kumyks" OR "Nogais" OR "Andis" OR "Akhvakh" OR "Archins" OR "Bagvalals" OR "Bezhta" OR "Botlikhs" OR "Chamalals" OR "Godoberi" OR "Hinukh" OR "Hunzibs" OR "Khwarshi" OR "Karata" OR "Tindis" OR "Tsez" OR "Abazin" OR "Besermyan" OR "Izhorians" OR "Karelians" OR "Nagaybaks" OR "Setos" OR "Shapsugs" OR "Quratay")) OR AB (("Ainus" OR "Ainu" OR "Aleuts" OR "Alyutors" OR "Chukchis" OR "Chuvans" OR "Dolgans" OR "Enets" OR "Entsy" OR "Yupik" OR "Yup'ik" OR "Yuit" OR "Yupigyt" OR "Chaplino" OR "Naukan" OR "Itelmens" OR "Kamchadals" OR "Kereks" OR "Komi" OR "Koryaks" OR "Nenets" OR "Nentsy" OR "Nganasans" OR "Tavgi" OR "Sami" OR "Veps" OR "Yukaghirs" OR "Chulyms" OR "Evenks" OR "Tungus" OR "Evens" OR "Kets" OR "Khantys" OR "Mansi" OR "Vguls" OR "Selkups" OR "Teleuts" OR "Nanais" OR "Nanaitsy" OR "Negidal" OR "Nivikh" OR "Oroch" OR "Orok" OR "Taz" OR "Udege" OR "Ulch" OR "Kumadins" OR "Chelkans" OR "Shorians" OR "Soyots" OR "Telengits" OR "Tofalars" OR "Tugalars" OR "Tufans" OR "Todzhins" OR "Laks" OR "Tabasarans" OR "Turuls" OR "Aguls" OR "Tsakhurs" OR "Kumyks" OR "Nogais" OR "Andis" OR "Akhvakh" OR "Archins" OR "Bagvalals" OR "Bezhta" OR "Botlikhs" OR "Chamalals" OR "Godoberi" OR "Hinukh" OR "Hunzibs" OR "Khwarshi" OR "Karata" OR "Tindis" OR "Tsez" OR "Abazin" OR "Besermyan" OR "Izhorians" OR "Karelians" OR "Nagaybaks" OR "Setos" OR "Shapsugs" OR "Quratay")) or (((MH "Indigenous Peoples+") or ((Indigenous or Aborig* or tribe or tribal or tribes) N3 (population* or people* or person* or elder* or man or men or woman or women or child* or youth* or clan or clans or tribe or tribes or tribal or family or families or parent* or grandparent* or elder or elders or grandmother* or grandfather* or baby or babies or infant or infants or patient or patients or speakers or speaking or village* or communit*))) and (Russia. or (MH "Russia")) )** | **816** |
| **S68** | **TI ( ( ("Abipon" OR "Achuar" OR "Achuagua" OR "Akawaio" OR "Amarizana" OR "Andoque" OR "Akawaio" OR "Akuriyo" OR "Anauya" OR "Araona" OR "Arawak" OR "Ayamn" OR "Aguaruna" OR "Amahuaca" OR "Amarakaeri" OR "Andoa" OR "Arabela" OR "Arawak" OR "Arhuaco" OR "Ashaninca" OR "Asheninca" OR "Atsahuaca" OR "Aymara" OR "Ayoreo" OR "Bakairi" OR "Baniva" OR "Barasana" OR "Baniwa" OR "Baure" OR "Bororo" OR "Cabiyari" OR "Cacataibo" OR "Caquinte" OR "Cacua" OR "Cahuarano" OR "Caiua" OR "Camara Indians" OR "Camaracoto" OR "Camsa" OR "Canamari" OR "Candoshi" OR "Canela" OR "Canichana" OR "Capanahua" OR "Carapana" OR "Cariay" OR " Carib " OR "Carijona" OR "Carutana" OR "Cashibo" OR "Cashinahua" OR "Cawishana" OR "Cavinena" OR "Caxuiana" OR "Cayuvava" OR "Chontaquiro" OR "Cocama" OR " Cubeo " OR "Curipaco" OR "Chacobo" OR "Chaima" OR ("Chana" not "striatus") OR "Chapacura" OR "Charrua" OR "Chimila" OR "Chitonahua" OR "Chorote" OR "Chipaya" OR "Chiquitano" OR "Chulupi" OR "Carare" OR "Coconuco" OR "Cofan" OR "Coreguaje" OR "Coyaima" OR "Chamacoco" OR "Chamicuro" OR "Chayahuita" OR "Cocama" OR " Culina " OR "Culino" OR "Cubeo" OR "Cuiba" OR " Cuiva " OR "Cumanagoto" OR "Curripaco" OR " Deni " OR "Desano" OR "Embera" OR "Guarani" OR "Guajajara" OR " Guana " OR "Guanano" OR "Guarayo" OR "Guarayu" OR "Guahibo" OR "Guajiro" OR "Guambiano" OR "Guanano" OR "Guayabero" OR "Guarequena" OR "Guinao" OR " Guana " OR "Gayon" OR "Guahibo" OR "Hixkaryana" OR "Huachipairi" OR "Huambisa" OR "Huarayo" OR "Lauanaua" OR "Ikpeng" OR "Ingariko" OR "Irantxe" OR "Itonama" OR "Inapari" OR "Iquito" OR "Isconahua" OR "Jumana" OR "Japreria" OR "Jirajara" OR "Juruti" OR "Jaqaru" OR "Jebero" OR "Kadiweu" OR "Kaingang" OR "Kamayura" OR "Karaja" OR "Karipuna" OR "Kariri" OR "Katukina" OR "Kaxarari" OR "Kayabi" OR "Kayapo" OR "Kuikuro alapalo" OR "Kulina" OR " Kaiwa " OR "Kallawaya" OR "Kogui" OR "Kuna " OR "Kaweskar" OR "Lule" OR "Macuna" OR "Maipure" OR "Mapuche" OR "Mataco" OR "Mocovi" OR "Machinere" OR "Machinerev" OR "Machiguenga" OR "Macushi" OR "Macuna" OR "Madi" OR "Malayo" OR "Mamainde" OR "Manao" OR "Mandauaca" OR "Mandawaka" OR "Mapidian" OR "Mapuche" OR "Mapidian" OR "Maquiritare" OR "Maquiritari" OR "Maragua" OR "Marawan" OR "Mariate" OR "Marubo" OR "Mastanahua" OR "Mataco" OR "Matipuhy" OR "Matis" OR "Matses" OR "Mawakua" OR "Mawakwa" OR "Maxakali" OR "Mehinaku" OR "Miranha" OR "Moronawa" OR "Munduruku" OR "Movima" OR "Muellama" OR "Muinane" OR "Mapoyo" OR " Mashco Piro " OR "Muniche" OR "Nambikwara" OR "Nocaman" OR "Nuquini" OR "Nomatsiguenga" OR "Nanti" OR "Ocaina" OR "Omagua" OR "Orejon" OR "Opon" OR "Pacahuara" OR "Pae" OR "Paicone" OR "Palicur" OR "Panare" OR "Pano" OR "Pares" OR "Paumari" OR "Pemon " OR "Pilaga" OR "Puelche" OR "Pauna" OR "Pauserna" OR "Piapoco" OR "Piraha" OR "Piratapuyo" OR "Pisabo" OR "Piaroa" OR "Pijao" OR "Piratapuyo" OR "Paraujano" OR "Pemon" OR "Pemono" OR "Piapoco" OR "Puinave" OR "Patamona" OR "Poyanawa" OR "Puinave" OR "Puquina" OR "Quechua" OR "Quichua" OR "Retuara" OR "Resigaro" OR "Reyesano" OR "Sabanes" OR "Saliba " OR "Saluma" OR "Sarave" OR "Secoya" OR "Selknam" OR " Sensi " OR "Shaninawa" OR "Shapra" OR "Sharanahua" OR "Shebayo" OR "Shiwiar" OR "Shikiana" OR "Sikiana" OR "Siriono" OR "Sinsiga" OR " Siona " OR "Suruwaha" OR "Tacano" OR "Tamanaco" OR "Tiahuanaco" OR "Tariano" OR "Tehuelche" OR "Tariano" OR "Tatuyo" OR "Tembe" OR "Terena" OR "Telembi" OR "Ticuna" OR "Ticuna" OR "Tiriyo" OR "Tiwanaku" OR "Tiwanaku" OR "Torom" OR "Totoro " OR "Tsimane" OR "Tuberao" OR "Tucano " OR "Tunebo" OR "Tuxinawa" OR "Tuyuca" OR "Uainuma" OR "Urarina" OR "Vilela" OR "Waimaha" OR "Waiampi" OR "Waiwai" OR "Wapishana" OR "Waraiku" OR "Warekena" OR "Waura" OR "Wayampi" OR "Wayana" OR "Wirina" OR "Waimaha" OR "Waunana" OR "Wiwa" OR "Warao" OR "Wayuu" OR "Witoto" OR "Xavante" OR "Xipaya" OR "Xiriana" OR "Xokleng" OR "Yabaana" OR "Yaminawa" OR "Yaminahua" OR "Yaruma" OR "Yawalapiti" OR "Yuracare" OR "Yabarana" OR "Yavitero" OR "Yine" OR "Yamana" OR "Yaghan" OR "Yucuna" OR "Yurumangui" OR "Yukpa" OR "Yanesha" OR "Yoranahua" OR "Yagua" OR "Yaminahua" OR "Zaparo" OR "Zamuco" OR ( ( "Inga" OR "Maca" OR "Leco" OR "Mojo" OR "Uro" OR "Maco" OR "Lengua" OR "Toba " OR "Zoe" OR "Ona" OR "Catio" OR "Passe" OR "Bari" OR "Awa" OR "Bora" OR "Bara" OR "Remo" OR "Pano" OR "Sape" ) N3 ("Indians" OR "Indian" OR "Indigenous" OR "Amerindian*" OR "Aborigin*" OR "people" OR "peoples" OR "elder" OR "elders" OR "grandmother*" OR "grandfather*" OR "parent*" OR "women" OR "men" OR "woman" OR "man" OR "child*" OR "youth" OR "youths" OR "baby" OR "babies" OR "tribe" OR "tribes" OR "tribal" OR "shaman*" OR "native" OR "patient*)) OR "Trio Indians" OR "More Indians" OR "Bare Indians" ) ) OR AB ( ( ("Abipon" OR "Achuar" OR "Achuagua" OR "Akawaio" OR "Amarizana" OR "Andoque" OR "Akawaio" OR "Akuriyo" OR "Anauya" OR "Araona" OR "Arawak" OR "Ayamn" OR "Aguaruna" OR "Amahuaca" OR "Amarakaeri" OR "Andoa" OR "Arabela" OR "Arawak" OR "Arhuaco" OR "Ashaninca" OR "Asheninca" OR "Atsahuaca" OR "Aymara" OR "Ayoreo" OR "Bakairi" OR "Baniva" OR "Barasana" OR "Baniwa" OR "Baure" OR "Bororo" OR "Cabiyari" OR "Cacataibo" OR "Caquinte" OR "Cacua" OR "Cahuarano" OR "Caiua" OR "Camara Indians" OR "Camaracoto" OR "Camsa" OR "Canamari" OR "Candoshi" OR "Canela" OR "Canichana" OR "Capanahua" OR "Carapana" OR "Cariay" OR " Carib " OR "Carijona" OR "Carutana" OR "Cashibo" OR "Cashinahua" OR "Cawishana" OR "Cavinena" OR "Caxuiana" OR "Cayuvava" OR "Chontaquiro" OR "Cocama" OR " Cubeo " OR "Curipaco" OR "Chacobo" OR "Chaima" OR ("Chana" not "striatus") OR "Chapacura" OR "Charrua" OR "Chimila" OR "Chitonahua" OR "Chorote" OR "Chipaya" OR "Chiquitano" OR "Chulupi" OR "Carare" OR "Coconuco" OR "Cofan" OR "Coreguaje" OR "Coyaima" OR "Chamacoco" OR "Chamicuro" OR "Chayahuita" OR "Cocama" OR " Culina " OR "Culino" OR "Cubeo" OR "Cuiba" OR " Cuiva " OR "Cumanagoto" OR "Curripaco" OR " Deni " OR "Desano" OR "Embera" OR "Guarani" OR "Guajajara" OR " Guana " OR "Guanano" OR "Guarayo" OR "Guarayu" OR "Guahibo" OR "Guajiro" OR "Guambiano" OR "Guanano" OR "Guayabero" OR "Guarequena" OR "Guinao" OR " Guana " OR "Gayon" OR "Guahibo" OR "Hixkaryana" OR "Huachipairi" OR "Huambisa" OR "Huarayo" OR "Lauanaua" OR "Ikpeng" OR "Ingariko" OR "Irantxe" OR "Itonama" OR "Inapari" OR "Iquito" OR "Isconahua" OR "Jumana" OR "Japreria" OR "Jirajara" OR "Juruti" OR "Jaqaru" OR "Jebero" OR "Kadiweu" OR "Kaingang" OR "Kamayura" OR "Karaja" OR "Karipuna" OR "Kariri" OR "Katukina" OR "Kaxarari" OR "Kayabi" OR "Kayapo" OR "Kuikuro alapalo" OR "Kulina" OR " Kaiwa " OR "Kallawaya" OR "Kogui" OR "Kuna " OR "Kaweskar" OR "Lule" OR "Macuna" OR "Maipure" OR "Mapuche" OR "Mataco" OR "Mocovi" OR "Machinere" OR "Machinerev" OR "Machiguenga" OR "Macushi" OR "Macuna" OR "Madi" OR "Malayo" OR "Mamainde" OR "Manao" OR "Mandauaca" OR "Mandawaka" OR "Mapidian" OR "Mapuche" OR "Mapidian" OR "Maquiritare" OR "Maquiritari" OR "Maragua" OR "Marawan" OR "Mariate" OR "Marubo" OR "Mastanahua" OR "Mataco" OR "Matipuhy" OR "Matis" OR "Matses" OR "Mawakua" OR "Mawakwa" OR "Maxakali" OR "Mehinaku" OR "Miranha" OR "Moronawa" OR "Munduruku" OR "Movima" OR "Muellama" OR "Muinane" OR "Mapoyo" OR " Mashco Piro " OR "Muniche" OR "Nambikwara" OR "Nocaman" OR "Nuquini" OR "Nomatsiguenga" OR "Nanti" OR "Ocaina" OR "Omagua" OR "Orejon" OR "Opon" OR "Pacahuara" OR "Pae" OR "Paicone" OR "Palicur" OR "Panare" OR "Pano" OR "Pares" OR "Paumari" OR "Pemon " OR "Pilaga" OR "Puelche" OR "Pauna" OR "Pauserna" OR "Piapoco" OR "Piraha" OR "Piratapuyo" OR "Pisabo" OR "Piaroa" OR "Pijao" OR "Piratapuyo" OR "Paraujano" OR "Pemon" OR "Pemono" OR "Piapoco" OR "Puinave" OR "Patamona" OR "Poyanawa" OR "Puinave" OR "Puquina" OR "Quechua" OR "Quichua" OR "Retuara" OR "Resigaro" OR "Reyesano" OR "Sabanes" OR "Saliba " OR "Saluma" OR "Sarave" OR "Secoya" OR "Selknam" OR " Sensi " OR "Shaninawa" OR "Shapra" OR "Sharanahua" OR "Shebayo" OR "Shiwiar" OR "Shikiana" OR "Sikiana" OR "Siriono" OR "Sinsiga" OR " Siona " OR "Suruwaha" OR "Tacano" OR "Tamanaco" OR "Tiahuanaco" OR "Tariano" OR "Tehuelche" OR "Tariano" OR "Tatuyo" OR "Tembe" OR "Terena" OR "Telembi" OR "Ticuna" OR "Ticuna" OR "Tiriyo" OR "Tiwanaku" OR "Tiwanaku" OR "Torom" OR "Totoro " OR "Tsimane" OR "Tuberao" OR "Tucano " OR "Tunebo" OR "Tuxinawa" OR "Tuyuca" OR "Uainuma" OR "Urarina" OR "Vilela" OR "Waimaha" OR "Waiampi" OR "Waiwai" OR "Wapishana" OR "Waraiku" OR "Warekena" OR "Waura" OR "Wayampi" OR "Wayana" OR "Wirina" OR "Waimaha" OR "Waunana" OR "Wiwa" OR "Warao" OR "Wayuu" OR "Witoto" OR "Xavante" OR "Xipaya" OR "Xiriana" OR "Xokleng" OR "Yabaana" OR "Yaminawa" OR "Yaminahua" OR "Yaruma" OR "Yawalapiti" OR "Yuracare" OR "Yabarana" OR "Yavitero" OR "Yine" OR "Yamana" OR "Yaghan" OR "Yucuna" OR "Yurumangui" OR "Yukpa" OR "Yanesha" OR "Yoranahua" OR "Yagua" OR "Yaminahua" OR "Zaparo" OR "Zamuco" OR ( ( "Inga" OR "Maca" OR "Leco" OR "Mojo" OR "Uro" OR "Maco" OR "Lengua" OR "Toba " OR "Zoe" OR "Ona" OR "Catio" OR "Passe" OR "Bari" OR "Awa" OR "Bora" OR "Bara" OR "Remo" OR "Pano" OR "Sape" ) N3 ("Indians" OR "Indian" OR "Indigenous" OR "Amerindian*" OR "Aborigin*" OR "people" OR "peoples" OR "elder" OR "elders" OR "grandmother*" OR "grandfather*" OR "parent*" OR "women" OR "men" OR "woman" OR "man" OR "child*" OR "youth" OR "youths" OR "baby" OR "babies" OR "tribe" OR "tribes" OR "tribal" OR "shaman*" OR "native" OR "patient*)) OR "Trio Indians" OR "More Indians" OR "Bare Indians" ) ) OR ( ((MH Native Americans) OR ("Indian*" OR "Amerindian" OR "Aboriginal*" OR "Indigenas" OR "Indigenous")) and ((MH South American+) OR "Argentin*" OR "Bolivia*" OR "Brazil*" OR "Chile*" OR "Colombia*" OR "Ecuador*" OR " French Guiana " OR "Guyana*" OR "Paraguay*" OR "Peru" OR "Peruvian" OR "Suriname" OR "Uruguay*" OR "Venezuela*" OR " Amazon Region " OR "Amazonia" OR "Andes" OR "Andean") )** | **1,541** |
| **S69** | **(((MH "Asia, Southeastern+") or ("Bali" or "Borneo" or "Brunei" or Cambodia or "Flores Island" or "Indonesia" or Java or Komodo or "Laos" or "Lombok" or "Malasia" or "Malay Peninsula" or "Myanmar" or "Mekong Valley" or "New Guinea" or "Philippeans" or "Sabah" or "Singapore" or "Sumatra" or "Sumba" or "Sumbawa" or "Thailand" or "Timor" or "Vietnam")) and ((MH "Indigenous Peoples+")or ((Indigen* or aborig* or tribe or tribes or triabal) N3 (population* or people or peoples or person or persons or elder or elders or man or men or woman or women or child* or youth* or clan or clans or family or families or parent* or grandparent* or elder or elders or grandmother* or grandfather* or baby or babies or infant or infants or patient or patients or speakers or speaking or village* or communit*)))) or TI ("Abui" or "Aeta" or "Alfur" or "Ati People" or "Bahau" or "Bali Aga" or "Bali Mula" or "Baliaga" or "Balinese" or "Basap" or "Bataq" or "Batek" or "Bateq" or "Batin" or "Blaan" or "Brao" or "Bru" or "Bru-Van Kieu" or "Bugkalot" or "Bumiputera" or "Bunong" or "Chams" or "Champa" or "Cheq Wong" or "Dasun" or "Dayak" or "Deli Malay" or "Filipina*" or "Filiopino*" or "Gaddang" or "Ibaloi" or "Iban" or "Idan" or "Igorot" or "Jahai" or "Jehai" or "Jakun" or "Javanese Kshatriya" or "Kangeanese" or " Kagayanen" or "Kanayatn" or "Katu" or "Kayan" or "Kavet" or "Kenyah" or "Khmer Krom" or "Kimaragang" or "Klemantan" or "Kreung" or "Kuijau" or "Kuy" or "Kwijau" or "Lampung*" or "Lanoh" or "Lawangan" or "Luangan" or "Lumad" or "Dayak" or "Loloan Malay*" or "Mah Meri" or "Malay Singaporean*" or "Malbog" or "Maluku" or "Mangka'ak" or "Mangyan" or "Maniq" or "Maragang" or "Ma'anyan" or "Minokok" or "Moluccas" or Montagnard or "Murut" or "Naga" or "Negrito" or "New Guinea" or "Ngaju" or "Orang Hulu" or "Orang Kanaq" or "Orang Kuala" or "Orang Seletar" or "Orang Ulu" or "Ot Danum" or "Palawano" or "Palembang*" or " Panay-Bukidnon" or "Pan-ayanon" or "Papua" or "Phnong" or "Punong" or "Pear People" or "Por People" or "Punan" or "Rumanau" or "Sasak" or "Semai" or "Semang" or "Semaq Beri" or "Semelai" or "Senoi" or "Simeulue" or "Suludnon" or "Sunda Island*" or "Tabanwa" or "Takua" or "Tambanuo" or " Tombonuo" or "Taron" or "Temiar*" or "Temoq" or "Temuan" or "Tidung" or "Trone" or "Taaw't Bato") OR AB ("Abui" or "Aeta" or "Alfur" or "Ati People" or "Bahau" or "Bali Aga" or "Bali Mula" or "Baliaga" or "Balinese" or "Basap" or "Bataq" or "Batek" or "Bateq" or "Batin" or "Blaan" or "Brao" or "Bru" or "Bru-Van Kieu" or "Bugkalot" or "Bumiputera" or "Bunong" or "Chams" or "Champa" or "Cheq Wong" or "Dasun" or "Dayak" or "Deli Malay" or "Filipina*" or "Filiopino*" or "Gaddang" or "Ibaloi" or "Iban" or "Idan" or "Igorot" or "Jahai" or "Jehai" or "Jakun" or "Javanese Kshatriya" or "Kangeanese" or " Kagayanen" or "Kanayatn" or "Katu" or "Kayan" or "Kavet" or "Kenyah" or "Khmer Krom" or "Kimaragang" or "Klemantan" or "Kreung" or "Kuijau" or "Kuy" or "Kwijau" or "Lampung*" or "Lanoh" or "Lawangan" or "Luangan" or "Lumad" or "Dayak" or "Loloan Malay*" or "Mah Meri" or "Malay Singaporean*" or "Malbog" or "Maluku" or "Mangka'ak" or "Mangyan" or "Maniq" or "Maragang" or "Ma'anyan" or "Minokok" or "Moluccas" or Montagnard or "Murut" or "Naga" or "Negrito" or "New Guinea" or "Ngaju" or "Orang Hulu" or "Orang Kanaq" or "Orang Kuala" or "Orang Seletar" or "Orang Ulu" or "Ot Danum" or "Palawano" or "Palembang*" or " Panay-Bukidnon" or "Pan-ayanon" or "Papua" or "Phnong" or "Punong" or "Pear People" or "Por People" or "Punan" or "Rumanau" or "Sasak" or "Semai" or "Semang" or "Semaq Beri" or "Semelai" or "Senoi" or "Simeulue" or "Suludnon" or "Sunda Island*" or "Tabanwa" or "Takua" or "Tambanuo" or " Tombonuo" or "Taron" or "Temiar*" or "Temoq" or "Temuan" or "Tidung" or "Trone" or "Taaw't Bato")** | **1,762** |
| **S70** | **(MM "Native Americans") OR (MH "Medicine, Native American Traditional") OR (MM "Health Services, Indigenous") OR (MH "Indigenous Health") or TI ("A' ani" or "Absaroka" or "Haaninin" or "Atsina" or "Gros Ventre" or "Acopsel" or "Tlacopsel" or "Lacopsel" or "Ahtna" or "Ahtena" or "Akenitsi" or "Occaneechi" or "Akokisa" or "Horcoquisa" or "Orcoquizas" or "Aleut" or "Unangax" or "Unangan" or "Alibamu" or "Alabama Alsea" or "Alutiiq" or "Sugpiag" or "Pacific Yupik" or "Amahami" or "Awaxawi" or "Androscoggin" or "Arosaguntacook" or "Ameriscoggin" or "Anishinaabeg" or "Chippewa" or "Anihsinape" or "Saulteaux" or "Apalachee" or "Aranama" or "Texan Coahuilteca" or "Tamique" or "Arikara" or "Sahnish" or "Arickaree" or "Adakadaho" or "Assiniboine" or "Hohe" or "Nakota" or "Nakoda" or "Nakona" or "Atsa' Kudok-wa" or "Awatixa" or "Bannock" or "Snake Indian*" or "Bidai" or "Quasmigdo" or "Biloxi" or "Blackfoot" or "Niitsitapi" or "Sikasikaitsitapi" or "Cahto" or "Kaipomo" or "Cahuilla" or "Ivilyuqaletem" or "Ivilyuat" or "Catawba" or "Inna" or "Iswa" or "Chemehuevi" or "Chickasaw" or "Chilula Chimakum" or "Aqokulo" or "Chimariko" or "Chiricahua" or "Tsokanende" or "Chitimacha" or "Chetimachan" or "Sitimacha" or "Chowanoke" or "Roanoke" or "Chumash" or "Ciboney" or "Taino Ciwat" or "Clatsop" or "Coos" or "Coosa" or "Uchis" or "Chiaha" or "Coste" or "Talisi" or "Coquille" or "Kokwell" or "Coso" or "Cowlitz" or "Taitnapam" or "Crow Nation" or "Cui Ui Ticutta" or "Cupeno" or "Kuupangaxwichem" or "Cupa" or "Cup' ig" or "Nunivak" or "Dakota Oyate" or "Lakota" or "Nakota" or "Santee" or "Teton" or "Sioux" or "Deadose" or "Deg Xina" or "Deg Xit' an" or "Kaiyuhkhotana" or "Deg Hit' an" or "Dena' ina" or "Tanaina" or "Dichinanek' Hwt' ana" or "Upper Kuskokwim Athabascan*" or "Kolchan" or "Goltsan" or "Tundra Kolosh" or "Do lkabya" or "Duwamish" or "Esselen" or "Eyak" or "Gidi' tikadi" or "Guwevkabaya" or "Gwich' in" or "Kutchin" or "Haida" or "Xaadas" or "Xaat" or "Halchidhoma" or "Havasupai" or "Green Water People" or "Hiratsa" or "Hiraaca" or "Ho-chaaqa" or "Winnebago" or "Holikachuk" or "Innoko" or "Tlegon-khotana" or "Hopi" or "Houma-Louisiana" or "Huaco" or "Waco" or "Hualapai" or "Hupa" or "Natinixwe" or "Natinook-wa" or "Hwech' in" or "Hankutchin" or "Iroquois Confederacy" or "Hodinoso ni" or "Illinois Confedera*" or "Ilinoweg" or "Illini" or "Inupiat" or "Inuit" or "Ioway" or "Baxoje" or "Jicarilla" or "Juaneno" or "Acjachemen" or "Jumano" or "Kalapuya" or "Clackama" or "Kalispel" or "Pend d' Oreilles" or "Qlispe" or "Karuk" or "Karok" or "Chum-ne" or "Katkoc" or "Kansa" or "Kanza" or "Kawaiisu" or "Nuwa" or "Kennebec" or "Kinipekw Kittitas" or "Klickitat" or "Qwu' lh-hwai-pum" or "Awi-adshi" or "Mahane" or "Wahnookt" or "Koa' aga' itoka" or "Keresan" or "Kichai" or "Kitsai" or "Keechi" or "K' itaish" or "Kiowa" or "Gaigwu" or "Cauigu" or "Kutjau" or "Kwu-da" or "Tep-da" or "Kitanemuk" or "Kittitas" or "Klickitat" or "Qwu' lh-hwai-pum" or "Awi-adshi" or "Mahane" or "Wahnookt" or "Koa' aga' itoka" or "Konkow" or "Koop Ticutta" or "Koyukon" or "Ktunaxa" or "Kootenai" or "Flathead" or "Kucadikadi" or "Kotsa' va" or "Kumeyaay" or "Tipai-Ipai" or "Kamia" or "Diegueno" or "Kwapa" or "Cocopah" or "Cucapa" or "Xawitt kwnchawaay" or "Lassik" or "Lenape" or "Leni-Lenape" or "Lipan" or "Luiseno" or "Payomkawichum" or "Madqwadabaya" or "Desert Yavapai" or "Mahican" or "Mohicans" or "Makah" or "Makuhadokado" or "Maliseet" or "Wolistoqiag" or "Manahoac" or "Mahock" or "Meipontsky" or "Mandan" or "Mattole" or "Bear River" or "Tul' bush" or "Ni' ekeni" or "Meherrin" or "Menominee" or "Mackinac" or "Mescalero" or "Myaamiaki" or "Kickapoo" or "Twigtwee" or "Missouria" or "Miwok" or "Miwuk" or "Moadokado" or "Modoc" or "Mohave" or "Aha Makhav" or "Mohawk" or "Kaneng' hega" or "Molala" or "Molale" or "Molele" or "Nyyhmy" or "Moosonee" or "Moose Cree" or "Monsonis" or "Multnomah" or "Chinook" or "Nabedache" or "Nabaydacu" or "Wawadishe" or "Nabiltse" or "Dakubetede" or "Nacho Nyak Dun" or "Tutchone" or "Nacono" or "Na' isha" or "Nanticoke" or "Navajo" or "Ndee" or "Nial" or "Niimiipu" or "Nez Perce" or "Watapala" or "Watapahlute" or "Nisenan" or "Nisqually" or "Nomlaki" or "Noamlakee" or "Central Wintun" or "Nongatl" or "Nottoway" or "Cheroenhaka" or "Northern Cheyenne" or "Ohlone" or "Costanoan" or "Omaha" or "O' odham" or "Pima" or "Papago" or "Osage" or "Otoe" or "Otse" or "Ozav Dika" or "Palus" or "Passamaquoddy" or "Pestomuhkati" or "Patiri" or "Petaros" or "Pastia" or "Patwin" or "Southern Wintun" or "Panis" or "Skidi" or "Pedee" or "Penobscot" or "Petun Piipaash" or "Kokmalik' op" or "Piscatawa" or "Doeg" or "Conoy" or "Pit River" or "Pomo" or "Kashaya" or "Ponca" or "Ponka" or "Pottawatomi" or "Bodewadmik" or "Powhatan" or "Puyallup" or "Spuyalepabs" or "Quapaw" or "Ugahxpa" or "Quechan" or "Yuma" or "Kwtsaan" or "Quileute" or "Salinan" or "Saponi" or "Monacan" or "Sapon" or "Eastern Blackfoot" or "Christanna" or "Sawawatodo" or "Serrano" or "Taaqtam" or "Maarenga' yam" or "Yuhaviatam" or "Shasta" or "Chasta" or "Sasti" or "Shoshone" or "Siletz" or "Sinkine" or "Sinkyone" or "Siuslaw Umpqua" or "Skitswish" or "Schitsu' umash" or "Snohomish" or "Snuqualmi" or "Sokoki" or "Missiquoi" or "Stillaguamish" or "Stoluckwamish" or "Suquamish" or "Sutaio" or "Swinomish" or "Skagit" or "Syilx" or "Okanagan" or "Sotaae" or "Taga Ticutta" or "Takelma" or "Dagelma" or "Taltushtuntede" or "Galice" or "Tanan Gwich' in" or "Taos" or "Taovaya" or "Tataviam" or "Alliklik" or "Tawakoni" or "Tahuacano" or "Tenino" or "Thawikila" or "Hathawekela" or "Fort Ancient" or "Tigua" or "Tillamook" or "Nehalem" or "Timbisha" or "Panamint" or "Timpanogos" or "Tlingit" or "Toi Ticutta" or "Tolowa" or "Talawa Dini' " or "Tongva" or "Gabrieleno" or "Fernandeno" or "Tobikhar" or "Tonkawa" or "Ticanwatic" or "Tsikip" or "Appalousa" or "Opelousa" or "Tsitsistas" or "Tubatulabal" or "Tukabatchee" or "Tuscarora" or "Tomahittan" or "Kuskarawock" or "Tutelo" or "Tutero" or "Totteroy" or "Tutera" or "Yusan" or "Tututni" or "Umatilla" or "Umpqua" or "Waccamaw" or "Waxmaw" or "Wadatika" or "Harney Valley Paiute" or "Wailiki" or "Waluulapam" or "Walla Walla" or "Walpapi" or "Huipui" or "Wampanoag" or "Massasoit" or "Wanapum" or "Wappo" or "Washoe" or "Wichita" or "Willapa" or "Kwalhioqua" or "Wi pukba" or "Verde Valley Yavapai" or "Wintu" or "Northern Wintun" or "Wiyot" or "Wee' at" or "Weyet" or "Yakama" or "Yamosopo Tuviwarai" or "Yaqui" or "Yoeme" or "Yatasi" or "Yattasih" or "Yavbe' " or "Yavapai" or "Ysleta del Sur" or "Yojuane" or "Yokuts" or "Mariposa" or "Yuki" or "Yupighyt" or "Yup'ik" or "Yupik" or "Yurok" or "Olekwo'l" or "Zuni" or (("Applegate" or "Delaware" or "Iowa" or "Ishak" or "Kaw" or "Kato" or "Spokane" or "Miami" or "Arkansas" or "Tali" or "Tunica" or ("Han" not (China or Chinese)) or "Pawnee" or "Coeur D' Alene" or "Piscataway" or "Ree" or "Tula" ) N3 (reservation* or nation or people or peoples or population or man or men or woman or women or child* or youth* or elder or elders or parent or parents or grandparent* or grandfather* or grandmother*or patient or patients or speaker* or speaking or village* or communit* or tribe or tribes or tribal or Indian*))) OR AB ("A' ani" or "Absaroka" or "Haaninin" or "Atsina" or "Gros Ventre" or "Acopsel" or "Tlacopsel" or "Lacopsel" or "Ahtna" or "Ahtena" or "Akenitsi" or "Occaneechi" or "Akokisa" or "Horcoquisa" or "Orcoquizas" or "Aleut" or "Unangax" or "Unangan" or "Alibamu" or "Alabama Alsea" or "Alutiiq" or "Sugpiag" or "Pacific Yupik" or "Amahami" or "Awaxawi" or "Androscoggin" or "Arosaguntacook" or "Ameriscoggin" or "Anishinaabeg" or "Chippewa" or "Anihsinape" or "Saulteaux" or "Apalachee" or "Aranama" or "Texan Coahuilteca" or "Tamique" or "Arikara" or "Sahnish" or "Arickaree" or "Adakadaho" or "Assiniboine" or "Hohe" or "Nakota" or "Nakoda" or "Nakona" or "Atsa' Kudok-wa" or "Awatixa" or "Bannock" or "Snake Indian*" or "Bidai" or "Quasmigdo" or "Biloxi" or "Blackfoot" or "Niitsitapi" or "Sikasikaitsitapi" or "Cahto" or "Kaipomo" or "Cahuilla" or "Ivilyuqaletem" or "Ivilyuat" or "Catawba" or "Inna" or "Iswa" or "Chemehuevi" or "Chickasaw" or "Chilula Chimakum" or "Aqokulo" or "Chimariko" or "Chiricahua" or "Tsokanende" or "Chitimacha" or "Chetimachan" or "Sitimacha" or "Chowanoke" or "Roanoke" or "Chumash" or "Ciboney" or "Taino Ciwat" or "Clatsop" or "Coos" or "Coosa" or "Uchis" or "Chiaha" or "Coste" or "Talisi" or "Coquille" or "Kokwell" or "Coso" or "Cowlitz" or "Taitnapam" or "Crow Nation" or "Cui Ui Ticutta" or "Cupeno" or "Kuupangaxwichem" or "Cupa" or "Cup' ig" or "Nunivak" or "Dakota Oyate" or "Lakota" or "Nakota" or "Santee" or "Teton" or "Sioux" or "Deadose" or "Deg Xina" or "Deg Xit' an" or "Kaiyuhkhotana" or "Deg Hit' an" or "Dena' ina" or "Tanaina" or "Dichinanek' Hwt' ana" or "Upper Kuskokwim Athabascan*" or "Kolchan" or "Goltsan" or "Tundra Kolosh" or "Do lkabya" or "Duwamish" or "Esselen" or "Eyak" or "Gidi' tikadi" or "Guwevkabaya" or "Gwich' in" or "Kutchin" or "Haida" or "Xaadas" or "Xaat" or "Halchidhoma" or "Havasupai" or "Green Water People" or "Hiratsa" or "Hiraaca" or "Ho-chaaqa" or "Winnebago" or "Holikachuk" or "Innoko" or "Tlegon-khotana" or "Hopi" or "Houma-Louisiana" or "Huaco" or "Waco" or "Hualapai" or "Hupa" or "Natinixwe" or "Natinook-wa" or "Hwech' in" or "Hankutchin" or "Iroquois Confederacy" or "Hodinoso ni" or "Illinois Confedera*" or "Ilinoweg" or "Illini" or "Inupiat" or "Inuit" or "Ioway" or "Baxoje" or "Jicarilla" or "Juaneno" or "Acjachemen" or "Jumano" or "Kalapuya" or "Clackama" or "Kalispel" or "Pend d' Oreilles" or "Qlispe" or "Karuk" or "Karok" or "Chum-ne" or "Katkoc" or "Kansa" or "Kanza" or "Kawaiisu" or "Nuwa" or "Kennebec" or "Kinipekw Kittitas" or "Klickitat" or "Qwu' lh-hwai-pum" or "Awi-adshi" or "Mahane" or "Wahnookt" or "Koa' aga' itoka" or "Keresan" or "Kichai" or "Kitsai" or "Keechi" or "K' itaish" or "Kiowa" or "Gaigwu" or "Cauigu" or "Kutjau" or "Kwu-da" or "Tep-da" or "Kitanemuk" or "Kittitas" or "Klickitat" or "Qwu' lh-hwai-pum" or "Awi-adshi" or "Mahane" or "Wahnookt" or "Koa' aga' itoka" or "Konkow" or "Koop Ticutta" or "Koyukon" or "Ktunaxa" or "Kootenai" or "Flathead" or "Kucadikadi" or "Kotsa' va" or "Kumeyaay" or "Tipai-Ipai" or "Kamia" or "Diegueno" or "Kwapa" or "Cocopah" or "Cucapa" or "Xawitt kwnchawaay" or "Lassik" or "Lenape" or "Leni-Lenape" or "Lipan" or "Luiseno" or "Payomkawichum" or "Madqwadabaya" or "Desert Yavapai" or "Mahican" or "Mohicans" or "Makah" or "Makuhadokado" or "Maliseet" or "Wolistoqiag" or "Manahoac" or "Mahock" or "Meipontsky" or "Mandan" or "Mattole" or "Bear River" or "Tul' bush" or "Ni' ekeni" or "Meherrin" or "Menominee" or "Mackinac" or "Mescalero" or "Myaamiaki" or "Kickapoo" or "Twigtwee" or "Missouria" or "Miwok" or "Miwuk" or "Moadokado" or "Modoc" or "Mohave" or "Aha Makhav" or "Mohawk" or "Kaneng' hega" or "Molala" or "Molale" or "Molele" or "Nyyhmy" or "Moosonee" or "Moose Cree" or "Monsonis" or "Multnomah" or "Chinook" or "Nabedache" or "Nabaydacu" or "Wawadishe" or "Nabiltse" or "Dakubetede" or "Nacho Nyak Dun" or "Tutchone" or "Nacono" or "Na' isha" or "Nanticoke" or "Navajo" or "Ndee" or "Nial" or "Niimiipu" or "Nez Perce" or "Watapala" or "Watapahlute" or "Nisenan" or "Nisqually" or "Nomlaki" or "Noamlakee" or "Central Wintun" or "Nongatl" or "Nottoway" or "Cheroenhaka" or "Northern Cheyenne" or "Ohlone" or "Costanoan" or "Omaha" or "O' odham" or "Pima" or "Papago" or "Osage" or "Otoe" or "Otse" or "Ozav Dika" or "Palus" or "Passamaquoddy" or "Pestomuhkati" or "Patiri" or "Petaros" or "Pastia" or "Patwin" or "Southern Wintun" or "Panis" or "Skidi" or "Pedee" or "Penobscot" or "Petun Piipaash" or "Kokmalik' op" or "Piscatawa" or "Doeg" or "Conoy" or "Pit River" or "Pomo" or "Kashaya" or "Ponca" or "Ponka" or "Pottawatomi" or "Bodewadmik" or "Powhatan" or "Puyallup" or "Spuyalepabs" or "Quapaw" or "Ugahxpa" or "Quechan" or "Yuma" or "Kwtsaan" or "Quileute" or "Salinan" or "Saponi" or "Monacan" or "Sapon" or "Eastern Blackfoot" or "Christanna" or "Sawawatodo" or "Serrano" or "Taaqtam" or "Maarenga' yam" or "Yuhaviatam" or "Shasta" or "Chasta" or "Sasti" or "Shoshone" or "Siletz" or "Sinkine" or "Sinkyone" or "Siuslaw Umpqua" or "Skitswish" or "Schitsu' umash" or "Snohomish" or "Snuqualmi" or "Sokoki" or "Missiquoi" or "Stillaguamish" or "Stoluckwamish" or "Suquamish" or "Sutaio" or "Swinomish" or "Skagit" or "Syilx" or "Okanagan" or "Sotaae" or "Taga Ticutta" or "Takelma" or "Dagelma" or "Taltushtuntede" or "Galice" or "Tanan Gwich' in" or "Taos" or "Taovaya" or "Tataviam" or "Alliklik" or "Tawakoni" or "Tahuacano" or "Tenino" or "Thawikila" or "Hathawekela" or "Fort Ancient" or "Tigua" or "Tillamook" or "Nehalem" or "Timbisha" or "Panamint" or "Timpanogos" or "Tlingit" or "Toi Ticutta" or "Tolowa" or "Talawa Dini' " or "Tongva" or "Gabrieleno" or "Fernandeno" or "Tobikhar" or "Tonkawa" or "Ticanwatic" or "Tsikip" or "Appalousa" or "Opelousa" or "Tsitsistas" or "Tubatulabal" or "Tukabatchee" or "Tuscarora" or "Tomahittan" or "Kuskarawock" or "Tutelo" or "Tutero" or "Totteroy" or "Tutera" or "Yusan" or "Tututni" or "Umatilla" or "Umpqua" or "Waccamaw" or "Waxmaw" or "Wadatika" or "Harney Valley Paiute" or "Wailiki" or "Waluulapam" or "Walla Walla" or "Walpapi" or "Huipui" or "Wampanoag" or "Massasoit" or "Wanapum" or "Wappo" or "Washoe" or "Wichita" or "Willapa" or "Kwalhioqua" or "Wi pukba" or "Verde Valley Yavapai" or "Wintu" or "Northern Wintun" or "Wiyot" or "Wee' at" or "Weyet" or "Yakama" or "Yamosopo Tuviwarai" or "Yaqui" or "Yoeme" or "Yatasi" or "Yattasih" or "Yavbe' " or "Yavapai" or "Ysleta del Sur" or "Yojuane" or "Yokuts" or "Mariposa" or "Yuki" or "Yupighyt" or "Yup'ik" or "Yupik" or "Yurok" or "Olekwo'l" or "Zuni" or (("Applegate" or "Delaware" or "Iowa" or "Ishak" or "Kaw" or "Kato" or "Spokane" or "Miami" or "Arkansas" or "Tali" or "Tunica" or ("Han" not (China or Chinese)) or "Pawnee" or "Coeur D' Alene" or "Piscataway" or "Ree" or "Tula" ) N3 (reservation* or nation or people or peoples or population or man or men or woman or women or child* or youth* or elder or elders or parent or parents or grandparent* or grandfather* or grandmother*or patient or patients or speaker* or speaking or village* or communit* or tribe or tribes or tribal or Indian*)))** | **13,028** |
| **S71** | **( ( ((MH "Indigenous Peoples+") or ("indigenous people* " or aborigi* or tribe or tribal or tribes)) ) AND ( ((MH "Africa+") or ((Africa* not "African American*") or Algeria or Angola or Benin or Botswana or "Burkina Faso" or Burundi or Cameroon or "Cape Verde" or "Cabo Verde" or "Central African Republic" or Chad or Comoros or Congo or Djibouti or Egypt or "Equitorial Guinea" or Eritrea or Eswatini or Ethiopia or Gabon or Gambia or Ghana* or Guinea or "Guinea Bissau" or "Ivory Coast" or Kenya or Lesotho or Liberia or Libya or Madagascar or Malawi or Mali or Mauritania or Mauritius or Mayotte or Morocco or Mozambique or Namibia or "Niger" or Nigeria or Reunion or Rwanda or "Saint Helena" or Ascension or "Tristan de Cunha" or "Sao Tome" or Principe or Senegal or Seychelles or "Sierra Leone" or Somalia or "South Africa" or Sudan or Tanzania or Togo or Tunisia or Uganda or "Western Sahara" or Zambia or Zimbabwe)) ) ) OR TI ( ("Abakuria" or "Abaluhya" or "Abagusii" or "Abakuria" or "Aembu" or "Agikuyu" or "Akamba" or "Anuak" or "Anywaa" or "Amazigh" or "Ambala" or "Ambeere" or "Ambundu" or "Ambuun" or "Amharan" or "Angba" or "Baaka" or "Baamba" or "Babindi" or "Babini" or "Baboma" or "Bachokwe" or "Bacwa" or "Bafumbira" or "Baganda" or "Bagyele" or "Bagwere" or "Bagyeli" or "Bakiga" or "Bakola" or "Baholo" or "Bakalanga" or "Bakiga" or "Bakolo" or "Bakongo" or "Bakonjo" or "Baluba" or "Balunda" or "Balovale" or "Bamasaba" or "Bambuti" or "Bangala" or "Bangoli" or "Bangungu" or "Bantu" or "Banyankole" or "Banyarwanda" or "Banyole" or "Banyoro" or "Bapende" or "Bapedi" or "Barabaig" or "Barombi" or "Barundi" or "Baruuli" or "Basamia" or "Basoga" or "Batswana" or "Batooro" or "Batsamba" or "Batswana" or "Batwa" or "Bayaka" or "Bedzan" or "Bazombe" or "Bebayaka" or "Bedzan" or "Bhaca" or "Biaka" or "Borana" or "Chewa" or "Copts" or "Cormorian" or "Cushitic" or "Dahalo" or "Datooga" or "Dikidiki" or "Dogon" or "Ewondo" or "Fulani" or "Fuliru" or "Ganguela" or "Gciriku" or "Gyele" or "Hadza" or "Hadzabe" or "Haillom" or "Haratin" or "Herero" or "Himba" or "Hlubi" or "Iriryen" or "Iqvayliyen" or "Kabyle*" or "Kalenjin" or "Kanioka" or "Kanioka" or "Kaonde" or "Karamojong" or "Kavango" or "Kereuyu" or "Khoikhoi" or "KhoiSan" or "Kikuyu Kwangali" or "Lokele" or "Lowme" or "Lotuko" or "Lwalwa" or "Maasai" or "Makonde" or "Makua" or "Mande" or "Masalit" or "Matumbi" or "Mayeuyi" or "Mayeyi" or "Mbenga" or "Mbukushu" or "Mbochi" or "Mboro" or "Mbuti" or "Medzan" or "Mijikenda" or "Mozabite*" or "Nafusa" or "Ndebele" or "Ngombe" or "Namaqua" or "Nyanga" or "Nyamwezi" or "Ogiek" or "Ovambo" or "Ovimbundu" or "Phuthi" or "Pokomo" or "Rendille" or "Riffian" or "Riffians" or "Sakuma" or "Samburu" or "Sandawe" or "Sangha" or "Sango" or "Sengwer" or "Serer" or "Sesotho" or "Shangaan" or "Shawiya" or "Shenwa" or "Shi" or "Shilluk" or "Sukua" or "Sukus" or "Swahili" or "Tabwa" or "Tambuka" or "Taveta" or "Thembu" or "Tigrayan" or "Topoke" or "Tsonga" or "Toubou" or "Tuareg" or "Tumbuka" or "Ugana" or "Wochua" or "Xhosa" or "Xindonga" or "Yoruba" or "Zenati" or "Zuwara" or (("Indigenous" or "Afar" or "Afars" or "Aka" or "Akie" or "Ameru" or "Asua" or "Ateker" or "Atwot" or "Awjila" or "Bafia" or "Baka" or "Bakongo" or "Bakwe" or "Balunda" or "Balovale" or "Bango" or "Bassa" or "Beja" or "Bekpak" or "Bemba" or "Bembe" or "Benet" or "Berber" or "Berbers" or "Bira" or "Bowe" or "Bubi" or "Budja" or "Bulu" or "Bunrun" or "Chaga" or "Chopi" or "Damara" or "Dinka" or "Djerba" or "Duala" or "Dzing" or "Efe" or "Elmolo" or "Fang" or "Foora" or "Fula" or "Fur" or "Ghomara" or "Ghadames" or "Gllana" or "Glu" or "Gogo" or "Gongo" or "Haya" or "Havu" or "Hema" or "Hima" or "Hunde" or "Hutu" or "Huva" or "Iboko" or "Igbo" or "Ijo" or "Jieng" or "Kadu" or "Kande" or "Kango" or "Katla" or "Kgaga" or "Khoe" or "Kola" or "Komo" or "Kota" or "Kua" or "Kuba" or "Kwango" or "Kx'z" or "Kxoe" or "Lala" or "Lozi" or "Luo" or "Luba" or "Lupu" or "Masmuda" or "Matmata" or "Mbala" or "Mbam" or "Mbo" or "Mbolo" or "Mbuza" or "Mongo" or "Mpondo" or "Myene" or "Naadh" or "Nama" or "Nande" or "Naro" or "Ngoni" or "Ndau" or "Ndebele" or "Ngoli" or "Ngondi" or "Ngoni" or "Nguni" or "Nkoya" or "Nkumu" or "Nuba" or "Nubian" or "Nuer" or "Nzebi" or "Ogoni" or "Omoro" or "Oroko" or "Pygmy" or "Popoi" or "Poto" or "Puru" or "Rashad" or (San not ("San Francisco" or "San Diego" or "San Antonio")) or "Sango" or "Sanhaja" or "Sena" or "Shilha" or "Shira" or "Shona" or "Shua" or "Sokna" or "Somali*" or "Sotho" or "Sua" or "Subu" or "Swazi" or "Taitaa" or "Tchokwe" or "Teke" or "Tembo" or "Tetela" or (Tonga and Africa*) or "Tshwa" or "Tsoa" or "Twa" or "Turkana" or "Tuu" or "Venda" or "Vira" or "Watta" or "Wakuti" or "Yaaku" or "Yaka" or "Yakoma" or "Yanzi" or "Yao" or "Yeke" or "Yela" or "Yeyi" or "Zulu") adj3 (population* or people or peoples or person or persons or elder or elders or man or men or woman or women or child* or youth* or clan or clans or tribe or tribes or tribal or family or families or parent* or grandparent* or elder or elders or grandmother* or grandfather* or baby or babies or infant or infants or patient or patients or speakers or speaking or village* or communit*))) ) OR AB ( ("Abakuria" or "Abaluhya" or "Abagusii" or "Abakuria" or "Aembu" or "Agikuyu" or "Akamba" or "Anuak" or "Anywaa" or "Amazigh" or "Ambala" or "Ambeere" or "Ambundu" or "Ambuun" or "Amharan" or "Angba" or "Baaka" or "Baamba" or "Babindi" or "Babini" or "Baboma" or "Bachokwe" or "Bacwa" or "Bafumbira" or "Baganda" or "Bagyele" or "Bagwere" or "Bagyeli" or "Bakiga" or "Bakola" or "Baholo" or "Bakalanga" or "Bakiga" or "Bakolo" or "Bakongo" or "Bakonjo" or "Baluba" or "Balunda" or "Balovale" or "Bamasaba" or "Bambuti" or "Bangala" or "Bangoli" or "Bangungu" or "Bantu" or "Banyankole" or "Banyarwanda" or "Banyole" or "Banyoro" or "Bapende" or "Bapedi" or "Barabaig" or "Barombi" or "Barundi" or "Baruuli" or "Basamia" or "Basoga" or "Batswana" or "Batooro" or "Batsamba" or "Batswana" or "Batwa" or "Bayaka" or "Bedzan" or "Bazombe" or "Bebayaka" or "Bedzan" or "Bhaca" or "Biaka" or "Borana" or "Chewa" or "Copts" or "Cormorian" or "Cushitic" or "Dahalo" or "Datooga" or "Dikidiki" or "Dogon" or "Ewondo" or "Fulani" or "Fuliru" or "Ganguela" or "Gciriku" or "Gyele" or "Hadza" or "Hadzabe" or "Haillom" or "Haratin" or "Herero" or "Himba" or "Hlubi" or "Iriryen" or "Iqvayliyen" or "Kabyle*" or "Kalenjin" or "Kanioka" or "Kanioka" or "Kaonde" or "Karamojong" or "Kavango" or "Kereuyu" or "Khoikhoi" or "KhoiSan" or "Kikuyu Kwangali" or "Lokele" or "Lowme" or "Lotuko" or "Lwalwa" or "Maasai" or "Makonde" or "Makua" or "Mande" or "Masalit" or "Matumbi" or "Mayeuyi" or "Mayeyi" or "Mbenga" or "Mbukushu" or "Mbochi" or "Mboro" or "Mbuti" or "Medzan" or "Mijikenda" or "Mozabite*" or "Nafusa" or "Ndebele" or "Ngombe" or "Namaqua" or "Nyanga" or "Nyamwezi" or "Ogiek" or "Ovambo" or "Ovimbundu" or "Phuthi" or "Pokomo" or "Rendille" or "Riffian" or "Riffians" or "Sakuma" or "Samburu" or "Sandawe" or "Sangha" or "Sango" or "Sengwer" or "Serer" or "Sesotho" or "Shangaan" or "Shawiya" or "Shenwa" or "Shi" or "Shilluk" or "Sukua" or "Sukus" or "Swahili" or "Tabwa" or "Tambuka" or "Taveta" or "Thembu" or "Tigrayan" or "Topoke" or "Tsonga" or "Toubou" or "Tuareg" or "Tumbuka" or "Ugana" or "Wochua" or "Xhosa" or "Xindonga" or "Yoruba" or "Zenati" or "Zuwara" or (("Indigenous" or "Afar" or "Afars" or "Aka" or "Akie" or "Ameru" or "Asua" or "Ateker" or "Atwot" or "Awjila" or "Bafia" or "Baka" or "Bakongo" or "Bakwe" or "Balunda" or "Balovale" or "Bango" or "Bassa" or "Beja" or "Bekpak" or "Bemba" or "Bembe" or "Benet" or "Berber" or "Berbers" or "Bira" or "Bowe" or "Bubi" or "Budja" or "Bulu" or "Bunrun" or "Chaga" or "Chopi" or "Damara" or "Dinka" or "Djerba" or "Duala" or "Dzing" or "Efe" or "Elmolo" or "Fang" or "Foora" or "Fula" or "Fur" or "Ghomara" or "Ghadames" or "Gllana" or "Glu" or "Gogo" or "Gongo" or "Haya" or "Havu" or "Hema" or "Hima" or "Hunde" or "Hutu" or "Huva" or "Iboko" or "Igbo" or "Ijo" or "Jieng" or "Kadu" or "Kande" or "Kango" or "Katla" or "Kgaga" or "Khoe" or "Kola" or "Komo" or "Kota" or "Kua" or "Kuba" or "Kwango" or "Kx'z" or "Kxoe" or "Lala" or "Lozi" or "Luo" or "Luba" or "Lupu" or "Masmuda" or "Matmata" or "Mbala" or "Mbam" or "Mbo" or "Mbolo" or "Mbuza" or "Mongo" or "Mpondo" or "Myene" or "Naadh" or "Nama" or "Nande" or "Naro" or "Ngoni" or "Ndau" or "Ndebele" or "Ngoli" or "Ngondi" or "Ngoni" or "Nguni" or "Nkoya" or "Nkumu" or "Nuba" or "Nubian" or "Nuer" or "Nzebi" or "Ogoni" or "Omoro" or "Oroko" or "Pygmy" or "Popoi" or "Poto" or "Puru" or "Rashad" or (San not ("San Francisco" or "San Diego" or "San Antonio")) or "Sango" or "Sanhaja" or "Sena" or "Shilha" or "Shira" or "Shona" or "Shua" or "Sokna" or "Somali*" or "Sotho" or "Sua" or "Subu" or "Swazi" or "Taitaa" or "Tchokwe" or "Teke" or "Tembo" or "Tetela" or (Tonga and Africa*) or "Tshwa" or "Tsoa" or "Twa" or "Turkana" or "Tuu" or "Venda" or "Vira" or "Watta" or "Wakuti" or "Yaaku" or "Yaka" or "Yakoma" or "Yanzi" or "Yao" or "Yeke" or "Yela" or "Yeyi" or "Zulu") adj3 (population* or people or peoples or person or persons or elder or elders or man or men or woman or women or child* or youth* or clan or clans or tribe or tribes or tribal or family or families or parent* or grandparent* or elder or elders or grandmother* or grandfather* or baby or babies or infant or infants or patient or patients or speakers or speaking or village* or communit*))) )** | **2,472** |
| **S72** | **S55 OR S56 OR S57 OR S58 OR S59 OR S60 OR S61 OR S62 OR S63 OR S64 OR S65 OR S66 OR S67 OR S68 OR S69 OR S70 OR S71** | **100,724** |
| **S74** | **(S55 OR S56 OR S57 OR S58 OR S59 OR S60 OR S61 OR S62 OR S63 OR S64 OR S65 OR S66 OR S67 OR S68 OR S69 OR S70 OR S71) AND (S54 AND S72)** | **666** |
| **S75** | **(S55 OR S56 OR S57 OR S58 OR S59 OR S60 OR S61 OR S62 OR S63 OR S64 OR S65 OR S66 OR S67 OR S68 OR S69 OR S70 OR S71) AND (S54 AND S72) and Limit t 07-2023 to 02 - 2024** | **21** |

**ProQuest Dissertations and Theses Global Searched June 20, 2023 Result =373 and updated February 14, 2024 on the Web of Science Platform Result =9**

TS=(((Athapaskan OR Saulteaux OR Wakashan OR Cree OR Dene OR Inuit OR Inuk OR Inuvialuit* OR Haida OR Ktunaxa OR Tsimshian OR Gitxsan OR Gitksan OR "Nisga'a" OR Haisla OR Heiltsuk OR Oweenkeno OR "Kwakwaka'wakw" OR "Nuu chah nulth" OR "Tsilhqot'in" OR Dakelh OR "Wet'suwet'en" OR Sekani OR Dunne-za OR Dene OR Tahltan OR Kaska OR Tagish OR Tutchone OR Nuxalk OR Salish OR St'at'imc)) OR TS=((Stl'atl'imx OR Stl'atl'imc OR Nlaka'pamux OR Okanagan OR "Sec wepmc" OR Secwepemc OR Tlingit OR Anishinaabe OR Blackfoot OR Nakoda OR Tasttine OR "Tsuu T'ina" OR "Tsuut'ina" OR "Gwich'in")) OR TS=((Algonquin OR Nipissing OR Ojibwa OR Potawatomi OR Innu OR Maliseet OR "Mi'kmaq" OR Micmac OR Passamaquoddy OR Haudenosaunee OR Cayuga OR Mohawk OR Oneida OR Onondaga OR Seneca OR Tuscarora OR Wyandot OR Aboriginal* OR Indigenous* OR Metis)) OR TS=(("First Nation" OR "First Nations" OR Amerindian OR "torres strait islander" OR aborigine* OR "indigenous people" OR maori OR Saami OR Sami OR sampi OR Southernsami* OR Umesami* OR Pitesami* OR Lulesami* OR Northernsami* OR Enaresami* OR Kolasami* OR Lapp OR Lapps OR Lappish OR Lappland OR (Lapland* NOT longspur) OR Lappalainen* OR Saamelainen* OR reindeer herd* OR reindeer culture* OR reindeer pastoral* OR Lappbys OR Samebys OR reinbeitesdistrikt OR paliskunta OR siida)) OR TS=((Abakuria OR Abaluhya OR Abagusii OR Abakuria OR Aembu OR Agikuyu OR Akamba OR Anuak OR Anywaa OR Amazigh OR Ambala OR Ambeere OR Ambundu OR Ambuun OR Amharan OR Angba OR Baaka OR Baamba OR Babindi OR Babini OR Baboma OR Bachokwe OR Bacwa OR Bafumbira OR Baganda OR Bagyele OR Bagwere OR Bagyeli OR Bakiga OR Bakola OR Baholo OR Bakalanga OR Bakiga OR Bakolo OR Bakongo OR Bakonjo OR Baluba OR Balunda OR Balovale OR Bamasaba OR Bambuti OR Bangala OR Bangoli OR Bangungu OR Bantu OR Banyankole OR Banyarwanda OR Banyole OR Banyoro OR Bapende OR Bapedi OR Barabaig OR Barombi OR Barundi OR Baruuli OR Basamia OR Basoga OR Batswana OR Batooro OR Batsamba OR Batswana OR Batwa OR Bayaka OR Bedzan OR Bazombe OR Bebayaka OR Bedzan OR Bhaca OR Biaka OR Borana OR Chewa OR Copts OR Cormorian OR Cushitic OR Dahalo OR Datooga OR Dikidiki OR Dogon OR Ewondo OR Fulani OR Fuliru OR Ganguela OR Gciriku OR Gyele OR Hadza OR Hadzabe OR Haillom OR Haratin OR Herero OR Himba OR Hlubi OR Iriryen OR Iqvayliyen)) OR TS=((Kabyle* OR Kalenjin OR Kanioka OR Kanioka OR Kaonde OR Karamojong OR Kavango OR Kereuyu OR Khoikhoi OR KhoiSan OR Kikuyu Kwangali OR Lokele OR Lowme OR Lotuko OR Lwalwa OR Maasai OR Makonde OR Makua OR Mande OR Masalit OR Matumbi OR Mayeuyi OR Mayeyi OR Mbenga OR Mbukushu OR Mbochi OR Mboro OR Mbuti OR Medzan OR Mijikenda OR Mozabite* OR Nafusa OR Ndebele OR Ngombe OR Namaqua OR Nyanga OR Nyamwezi OR Ogiek OR Ovambo OR Ovimbundu OR Phuthi OR Pokomo OR Rendille OR Riffian OR Riffians OR Sakuma OR Samburu OR Sandawe OR Sangha OR Sango OR Sengwer OR Serer OR Sesotho OR Shangaan OR Shawiya OR Shenwa OR Shi OR Shilluk OR Sukua OR Sukus OR Swahili OR Tabwa OR Tambuka OR Taveta OR Thembu OR Tigrayan OR Topoke OR Tsonga OR Toubou OR Tuareg OR Tumbuka OR Ugana OR Wochua OR Xhosa OR Xindonga OR Yoruba OR Zenati OR Zuwara))) OR TS=(((Indigenous OR Afar OR Afars OR "Aka People" OR Akie OR Ameru OR Asua OR Ateker OR Atwot OR Awjila OR Bafia OR Baka OR Bakongo OR Bakwe OR Balunda OR Balovale OR Bango OR Bassa OR Beja OR Bekpak OR Bemba OR Bembe OR Benet OR Berber OR Berbers OR Bira OR Bowe OR Bubi OR Budja OR Bulu OR Bunrun OR Chaga OR Chopi OR Damara OR Dinka OR Djerba OR Duala OR Dzing OR Efe OR Elmolo OR Fang OR Foora OR Fula OR Fur OR Ghomara OR Ghadames OR Gllana OR Glu OR Gogo OR Gongo OR Haya OR Havu OR Hema OR Hima OR Hunde OR Hutu OR Huva OR Iboko OR Igbo OR Ijo OR Jieng OR Kadu OR Kande OR Kango OR Katla OR Kgaga OR Khoe OR Kola OR Komo OR Kota OR Kua OR Kuba OR Kwango OR Kx'z OR Kxoe OR Lala OR Lozi OR Luo OR Luba OR Lupu OR Masmuda OR Matmata OR Mbala OR Mbam OR "Mbo People" OR Mbolo OR Mbuza OR Mongo OR Mpondo OR Myene OR Naadh OR Nama OR Nande OR Naro OR Ngoni OR Ndau OR Ndebele OR Ngoli OR Ngondi OR Ngoni OR Nguni OR Nkoya OR Nkumu OR Nuba OR Nubian OR Nuer OR Nzebi OR Ogoni OR Omoro OR Oroko OR Pygmy OR Popoi OR Poto OR Puru OR Rashad OR Sango OR Sanhaja OR Sena OR Shilha OR Shira OR Shona OR Shua OR Sokna OR Somali* OR Sotho OR Sua OR Subu OR Swazi OR Taitaa OR Tchokwe OR Teke OR Tembo OR Tetela OR Tshwa OR Tsoa OR Twa OR Turkana OR Tuu OR Venda OR Vira OR Watta OR Wakuti OR Yaaku OR Yaka OR Yakoma OR Yanzi OR Yao OR Yeke OR Yela OR Yeyi OR Zulu) NEAR/3 (population* OR people* OR person* OR elder* OR man OR men OR woman OR women OR child* OR youth* OR clan OR clans OR tribe OR tribes OR tribal OR family OR families OR parent* OR grandparent* OR elder OR elders OR grandmother* OR grandfather* OR baby OR babies OR infant OR infants OR patient OR patients OR speakers OR speaking OR village* OR communit*))) OR TS=((Acatec OR Aguacateco OR Amuzgo OR Bokota OR Boruca OR Bribri OR "Bri Bri" OR Buglere OR Cabecar OR Cakchiquel OR Changuena OR Chatino OR Chiapanec OR Chicomuceltec OR Chinantee OR Chocho OR Cholti OR "Ch'olti'" OR "Ch'olti'anor Chontal" OR Chorotega OR Chorti OR Chuj OR Chumbia OR Corobici OR (Cueva NOT Spain) OR Cuicatec OR Cuitlatee OR Cuytec OR Dorasque OR Embera OR Garifuna OR Guatuso OR Guaymi OR Guaymis OR Guetar OR Huastec OR Huave OR Huetar OR Itzaj OR Ixil OR Jacalteco OR Jonaz OR Kanjobal OR Kekchi OR Kuna OR Maleku OR Mangue OR Matambu OR Matlatzinca OR Mazahua OR Motozintlec OR Mayan OR Mayangna OR Miskito OR Mixtec OR Mopan OR Nahua OR Nahuatl OR Ngabe OR Otomi OR Pantec OR Paya OR Popoloca OR Popoloc OR Poqomam OR Poqomchi OR "Q'eqchi'" OR Quiche OR Quitirrisi OR Sacapulteco OR Sipacapense OR Subtiaba OR Tacaneco OR Tarasco OR Tamaulipec OR Tamazultec OR Tecoxquin OR Tectiteco OR Tecual OR Tecuexe OR Tepehura OR Tepuztecor OR Teribe OR Terraba OR Totonac OR Trique OR Tzeltal OR Tzotzil OR Tzutujil OR Ulwa OR Uspantec* OR Uspanteko OR Voto OR Xinca OR Waunana OR Wounaan OR Yucatec OR Zapotec OR Zoque)) OR TS=(("Adivasis" OR "Adnamanese" OR "Andaman" OR "Baluch" OR "Baluchis" OR "Bodo" OR "Boro" OR "Boros" OR "Bote" OR "Brahuis" OR "Chakmas" OR "Chepang" OR "Chhantyal" OR "Damai" OR "Dewan" OR "Ghale" OR "Gurkha" OR "Gurung" OR "Hayu" OR "Hyolmo" OR "Jarawa" OR "Jirel" OR "Jumma" OR "Kalash" OR "Khas" OR "Kirati" OR "Koinch" OR "Kulung" OR "Kusunda" OR "Limbu" OR "Lohorung" OR "Magar" OR "Makrani" OR "Mangar" OR "Marma" OR "Miji" OR "Mongar" OR "Mro" OR "Naga" OR "Nepami" OR "Newar" OR "Nicobar" OR "Onge" OR "Rang" OR "Raute" OR "Sajolang" OR "Santhal" OR "Sentinelese" OR "Sindhis" OR "Sulemani" OR "Sunuwar" OR "Tamang" OR "Thakali" OR "Thangmi" OR "Tharu" OR "Tripura" OR "Tumbahangphe" OR "Wanniyala-Aetto" OR "Yakkha" OR "Yolmopa")) OR (TS=(Aguacatec) or TS=(Akwa'ala) or TS=(Abxubal) or TS=(Ayuukja'ay) or TS=("Batzil k'op") or TS=(Binizaa) or TS=("Chichimeca Jonaz") or TS=(Chinantec) or TS=(Chocho) or TS=("Ch ol") or TS=(Chontal) or TS=("Chuj") or TS=(Cochimi) or TS=(Comcaac) or TS=(Hamasipini) or TS=(Harijio) or TS=("Ha shuta enima") or TS=("Hach t'an") or TS=(Huastecor Hnahnu) or TS=(Hnatho) or TS=(Ixcatec) or TS=(Ixil) or TS=(Jacaltec) or TS=(K'akchikel) or TS=(K'anjobal) or TS=(Kanjobal) or TS=(Kaqchikel) or TS=(Kechi) or TS=(K'iche) or TS=(Kikapooa) or TS=(Kikapu) or TS=(Kiliwa) or TS=("Ko'lew") or TS=("K'op o winik atel") or TS=(Kumiai) or TS=(Lacandon) or TS=(Laymon) or TS=(Makurawe) or TS=(Maya) or TS=(Maya'wiinik) or TS=(Mazahua) or TS=(Mazatec) or TS=(Me'phaa) or TS=(Mexicanero) or TS=(Mexikatlajtolli) or TS=(Mixe) or TS=(Mixtec) or TS=(Motocintleco) or TS=(mti'pa) or TS=(Nahuas) or TS=(Ocuiltec) or TS=(Otomi) or (TS=(Oaxaca) not TS=(Oaxaca-Blinder)) or TS=("Pame") or TS=(Papago) or TS=(Tlahuica) or TS=(Paipai) or TS=("Pima Bajo") or TS=(Purepecha) or TS=(P'urhepecha) or TS=(Qatok) or (TS=(Quiche) not TS=(Guatemala)) or TS=(Q'iche) or TS=(Raramuri) or TS=("Runixa ngiigua") or (TS=("Seri") not TS=("Seri 82")) or TS=("Slijuala sihanuk") or TS=(Tacuate) or TS=(Tarahumara) or TS=(Teenek) or TS=(Tepehua) or TS=(Ti'pai) or TS=(Tlapanec) or TS=("Tohono O'odham") or TS=(Totonac) or TS=(Tachiwin) or TS=("Tsa jujmi") or TS=(Tzotzil) or TS=("Tu'un savi") or TS=(Tzeltal) or TS=("Uza") or TS=(Winik) or TS=(Xigue) or TS=(Yucatec) or TS=(Zapotec)) OR TI=(("A' ani" OR Absaroka OR Haaninin OR Atsina OR "Gros Ventre" OR Acopsel OR Tlacopsel OR Lacopsel OR Ahtna OR Ahtena OR Akenitsi OR Occaneechi OR Akokisa OR Horcoquisa OR Orcoquizas OR Aleut OR Unangax OR Unangan OR Alibamu OR "Alabama Alsea" OR Alutiiq OR Sugpiag OR Amahami OR Awaxawi OR Androscoggin OR Arosaguntacook OR Ameriscoggin OR Anishinaabeg OR Chippewa OR Anihsinape OR Saulteaux OR Apalachee OR Aranama OR "Texan Coahuilteca" OR Tamique OR Arikara OR Sahnish OR Arickaree OR Adakadaho OR Assiniboine OR Hohe OR Nakota OR Nakoda OR Nakona OR "Atsa' Kudok-wa" OR Awatixa OR Bannock OR "Snake Indian*" OR Bidai OR Quasmigdo OR Biloxi OR Blackfoot OR Niitsitapi OR Sikasikaitsitapi OR Cahto OR Kaipomo OR Cahuilla OR Ivilyuqaletem OR Ivilyuat OR Catawba OR Inna OR Iswa OR Chemehuevi OR Chickasaw OR "Chilula Chimakum" OR Aqokulo OR Chimariko OR Chiricahua OR Tsokanende OR Chitimacha OR Chetimachan OR Sitimacha OR Chowanoke OR Roanoke OR Chumash OR Ciboney OR "Taino Ciwat" OR Clatsop OR Coos OR Coosa OR Uchis OR Chiaha OR Coste OR Talisi OR Coquille OR Kokwell OR Coso OR Cowlitz OR Taitnapam OR "Crow Nation" OR "Cui Ui Ticutta" OR Cupeno OR Kuupangaxwichem OR Cupa OR "Cup' ig" OR Nunivak OR "Dakota Oyate" OR Lakota OR Nakota OR Santee OR Teton OR Sioux OR Deadose OR "Deg Xina" OR "Deg Xit' an" OR Kaiyuhkhotana OR "Deg Hit' an" OR "Dena' ina" OR Tanaina OR "Dichinanek' Hwt' ana" OR "Upper Kuskokwim Athabascan*" OR Kolchan OR Goltsan OR "Tundra Kolosh" OR "Do lkabya" OR Duwamish OR Esselen OR Eyak OR "Gidi' tikadi" OR Guwevkabaya OR "Gwich' in" OR Kutchin OR Haida OR Xaadas OR Xaat OR Halchidhoma OR Havasupai OR "Green Water People" OR Hiratsa OR Hiraaca OR "Ho-chaaqa" OR Winnebago OR Holikachuk OR Innoko OR "Tlegon-khotana" OR Hopi OR "Houma-Louisiana" OR Huaco OR Waco OR Hualapai OR Hupa OR Natinixwe OR "Natinook-wa" OR "Hwech' in" OR Hankutchin OR "Iroquois Confederacy" OR "Hodinoso ni" OR "Illinois Confedera*" OR Ilinoweg OR Illini OR Inupiat OR Inuit OR Ioway OR Baxoje OR Jicarilla OR Juaneno OR Acjachemen OR Jumano OR Kalapuya OR Clackama OR Kalispel OR "Pend d' Oreilles" OR Qlispe OR Karuk OR Karok OR "Chum-ne" OR Katkoc OR Kansa OR Kanza OR Kawaiisu OR Nuwa OR Kennebec OR "Kinipekw Kittitas" OR Klickitat OR "Qwu' lh-hwai-pum" OR "Awi-adshi" OR Mahane OR Wahnookt OR "Koa' aga' itoka" OR Keresan OR Kichai OR Kitsai OR Keechi OR "K' itaish" OR Kiowa OR Gaigwu OR Cauigu OR Kutjau OR "Kwu-da" OR "Tep-da" OR Kitanemuk OR Kittitas OR Klickitat OR "Qwu' lh-hwai-pum" OR "Awi-adshi" OR Mahane OR Wahnookt OR "Koa' aga' itoka" OR Konkow OR "Koop Ticutta" OR Koyukon OR Ktunaxa OR Kootenai OR Flathead OR Kucadikadi OR "Kotsa' va" OR Kumeyaay OR "Tipai-Ipai" OR Kamia OR Diegueno OR Kwapa OR Cocopah OR Cucapa OR "Xawitt kwnchawaay" OR Lassik OR Lenape OR "Leni-Lenape" OR Lipan OR Luiseno OR Payomkawichum OR Madqwadabaya OR "Desert Yavapai" OR Mahican OR Mohicans OR Makah OR Makuhadokado OR Maliseet OR Wolistoqiag OR Manahoac OR Mahock OR Meipontsky OR Mandan OR Mattole OR "Bear River" OR "Tul' bush" OR "Ni' ekeni" OR Meherrin OR Menominee OR Mackinac OR Mescalero OR Myaamiaki OR Kickapoo OR Twigtwee OR Missouria OR Miwok OR Miwuk OR Moadokado OR Modoc OR Mohave OR "Aha Makhav" OR Mohawk OR "Kaneng' hega" OR Molala OR Molale OR Molele OR Nyyhmy OR Moosonee OR "Moose Cree" OR Monsonis OR Multnomah OR Chinook OR Nabedache OR Nabaydacu OR Wawadishe OR Nabiltse OR Dakubetede OR "Nacho Nyak Dun" OR Tutchone OR Nacono OR "Na' isha" OR Nanticoke OR Navajo OR Ndee OR Nial OR Niimiipu OR "Nez Perce" OR Watapala OR Watapahlute OR Nisenan OR Nisqually OR Nomlaki OR Noamlakee OR "Central Wintun" OR Nongatl OR Nottoway OR Cheroenhaka OR "Northern Cheyenne" OR Ohlone OR Costanoan OR Omaha OR "O' odham" OR Pima OR Papago OR Osage OR Otoe OR Otse OR "Ozav Dika" OR Palus OR Passamaquoddy OR Pestomuhkati OR Patiri OR Petaros OR Pastia OR Patwin OR "Southern Wintun" OR Panis OR Skidi OR Pedee OR Penobscot OR "Petun Piipaash" OR "Kokmalik' op" OR Piscatawa OR Doeg OR Conoy OR "Pit River" OR Pomo OR Kashaya OR Ponca OR Ponka OR Pottawatomi OR Bodewadmik OR Powhatan OR Puyallup OR Spuyalepabs OR Quapaw OR Ugahxpa OR Quechan OR Yuma OR Kwtsaan OR Quileute OR Salinan OR Saponi OR Monacan OR Sapon OR "Eastern Blackfoot" OR Christanna OR Sawawatodo OR Serrano OR Taaqtam OR "Maarenga' yam" OR Yuhaviatam OR Shasta OR Chasta OR Sasti OR Shoshone OR Siletz OR Sinkine OR Sinkyone OR "Siuslaw Umpqua" OR Skitswish OR "Schitsu' umash" OR Snohomish OR Snuqualmi OR Sokoki OR Missiquoi OR Stillaguamish OR Stoluckwamish OR Suquamish OR Sutaio OR Swinomish OR Skagit OR Syilx OR Okanagan OR Sotaae OR "Taga Ticutta" OR Takelma OR Dagelma OR Taltushtuntede OR Galice OR "Tanan Gwich' in" OR Taos OR Taovaya OR Tataviam OR Alliklik OR Tawakoni OR Tahuacano OR Tenino OR Thawikila OR Hathawekela OR "Fort Ancient" OR Tigua OR Tillamook OR Nehalem OR Timbisha OR Panamint OR Timpanogos OR Tlingit OR "Toi Ticutta" OR Tolowa OR "Talawa Dini' " OR Tongva OR Gabrieleno OR Fernandeno OR Tobikhar OR Tonkawa OR Ticanwatic OR Tsikip OR Appalousa OR Opelousa OR Tsitsistas OR Tubatulabal OR Tukabatchee OR Tuscarora OR Tomahittan OR Kuskarawock OR Tutelo OR Tutero OR Totteroy OR Tutera OR Yusan OR Tututni OR Umatilla OR Umpqua OR Waccamaw OR Waxmaw OR Wadatika OR "Harney Valley Paiute" OR Wailiki OR Waluulapam OR "Walla Walla" OR Walpapi OR Huipui OR Wampanoag OR Massasoit OR Wanapum OR Wappo OR Washoe OR Wichita OR Willapa OR Kwalhioqua OR "Wi pukba" OR "Verde Valley Yavapai" OR Wintu OR "Northern Wintun" OR Wiyot OR "Wee' at" OR Weyet OR Yakama OR "Yamosopo Tuviwarai" OR Yaqui OR Yoeme OR Yatasi OR Yattasih OR "Yavbe" OR "Yavapai" OR "Ysleta del Sur" OR Yojuane OR Yokuts OR Mariposa OR Yuki OR Yupighyt OR "Yup'ik" OR Yupik OR Yurok OR "Olekwo'l" OR Zuni)) OR TI=((Abipon OR Achuar OR Achuagua OR Akawaio OR Amarizana OR Andoque OR Akawaio OR Akuriyo OR Anauya OR Araona OR Arawak OR Ayamn OR Aguaruna OR Amahuaca OR Amarakaeri OR Andoa OR Arabela OR Arawak OR Arhuaco OR Ashaninca OR Asheninca OR Atsahuaca OR Aymara OR Ayoreo OR Bakairi OR "Baniva" OR Barasana OR Baniwa OR "Baure" OR Bororo OR Cabiyari OR Cacataibo OR Caquinte OR Cacua OR Cahuarano OR "Caiua" OR "Camara Indians" OR Camaracoto OR Camsa OR Canamari OR Candoshi OR Canela OR Canichana OR Capanahua OR Carapana OR Cariay OR "Carib" OR Carijona OR Carutana OR Cashibo OR Cashinahua OR Cawishana OR Cavinena OR Caxuiana OR Cayuvava OR Chontaquiro OR Cocama OR "Cubeo" OR Curipaco OR Chacobo OR Chaima OR (Chana NOT striatus) OR Chapacura OR Charrua OR Chimila OR Chitonahua OR Chorote OR Chipaya OR Chiquitano OR Chulupi OR Carare OR Coconuco OR Cofan OR Coreguaje OR Coyaima OR Chamacoco OR Chamicuro OR Chayahuita OR Cocama OR "Culina" OR Culino OR Cubeo OR Cuiba OR "Cuiva" OR Cumanagoto OR Curripaco OR "Deni" OR Desano OR Embera OR Guarani OR Guajajara OR "Guana" OR Guanano OR Guarayo OR Guarayu OR Guahibo OR Guajiro OR Guambiano OR Guanano OR Guayabero OR Guarequena OR Guinao OR "Guana" OR Gayon OR Guahibo OR Hixkaryana OR Huachipairi OR Huambisa OR Huarayo OR Lauanaua OR Ikpeng OR Ingariko OR Irantxe OR Itonama OR Inapari OR Iquito OR Isconahua OR Jumana OR Japreria OR Jirajara OR Juruti OR Jaqaru OR Jebero OR Kadiweu OR Kaingang OR Kamayura OR Karaja OR Karipuna OR "Kariri" OR Katukina OR Kaxarari OR Kayabi OR Kayapo OR "Kuikuro alapalo" OR Kulina OR "Kaiwa" OR Kallawaya OR "Kogui" OR "Kuna" OR Kaweskar OR "Lule" OR Macuna OR Maipure OR Mapuche OR Mataco OR Mocovi OR Machinere OR Machinerev OR Machiguenga OR Macushi OR Macuna OR "Madi" OR Malayo OR Mamainde OR Manao OR Mandauaca OR Mandawaka OR Mapidian OR Mapuche OR Mapidian OR Maquiritare OR Maquiritari OR Maragua OR Marawan OR Mariate OR Marubo OR Mastanahua OR Mataco OR Matipuhy OR "Matis" OR "Matses" OR Mawakua OR Mawakwa OR Maxakali OR Mehinaku OR Miranha OR Moronawa OR Munduruku OR Movima OR Muellama OR Muinane OR Mapoyo OR "Mashco Piro" OR Muniche OR Nambikwara OR Nocaman OR Nuquini OR Nomatsiguenga OR Nanti OR Ocaina OR Omagua OR Orejon OR "Opon" OR Pacahuara OR "Paez" OR Paicone OR Palicur OR Panare OR "Pano" OR "Paresi" OR Paumari OR "Pemon" OR Pilaga OR Puelche OR Pauna OR Pauserna OR Piapoco OR Piraha OR Piratapuyo OR Pisabo OR Piaroa OR "Pijao" OR Piratapuyo OR Paraujano OR Pemon OR Pemono OR Piapoco OR Puinave OR Patamona OR Poyanawa OR Puinave OR Puquina OR Quechua OR Quichua OR Retuara OR Resigaro OR Reyesano OR Sabanes OR "Saliba" OR Saluma OR Sarave OR Secoya OR Selknam OR "Sensi" OR Shaninawa OR Shapra OR Sharanahua OR Shebayo OR Shiwiar OR Shikiana OR Sikiana OR Siriono OR Sinsiga OR "Siona" OR Suruwaha OR Tacano OR Tamanaco OR Tiahuanaco OR Tariano OR Tehuelche OR Tariano OR Tatuyo OR "Tembe" OR "Terena" OR Telembi OR Ticuna OR Ticuna OR Tiriyo OR Tiwanaku OR Tiwanaku OR "Torom" OR "Totoro" OR Tsimane OR Tuberao OR "Tucano" OR Tunebo OR Tuxinawa OR Tuyuca OR Uainuma OR Urarina OR Vilela OR Waimaha OR Waiampi OR Waiwai OR Wapishana OR Waraiku OR Warekena OR Waura OR Wayampi OR Wayana OR Wirina OR Waimaha OR Waunana OR "Wiwa" OR "Warao" OR Wayuu OR Witoto OR Xavante OR Xipaya OR Xiriana OR Xokleng OR Yabaana OR Yaminawa OR Yaminahua OR Yaruma OR Yawalapiti OR Yuracare OR Yabarana OR Yavitero OR "Yine" OR Yamana OR Yaghan OR Yucuna OR Yurumangui OR Yukpa OR Yanesha OR Yoranahua OR Yagua OR Yaminahua OR Zaparo OR Zamuco OR "Trio Indians" OR "More Indians" OR "Bare Indians")) OR TS=(((("Inga" OR "Maca" OR "Leco" OR "Mojo" OR "Uro" OR "Maco" OR Lengua OR "Toba" OR "Zoe" OR "Ona" OR "Catio" OR "Passe" OR "Bari" OR "Awa" OR "Bora" OR "Bara" OR "Remo" OR "Pano" OR "Sape") NEAR/3 (Indians OR Indian OR Indigenous OR Amerindian* OR Aborigin* OR people OR peoples OR elder OR elders OR grandmother* OR grandfather* OR parent* OR women OR men OR woman OR man OR child* OR youth OR youths OR baby OR babies OR tribe OR tribes OR tribal OR shaman* OR native OR patient OR patients)))) OR TI=((Ainus OR Ainu OR Aleuts OR Alyutors OR Chukchis OR Chuvans OR Dolgans OR Enets OR Entsy OR Yupik OR Yup'ik OR Yuit OR Yupigyt OR Chaplino OR Naukan OR Itelmens OR Kamchadals OR Kereks OR "Komi" OR Koryaks OR Nenets OR Nentsy OR Nganasans OR Tavgi OR Sami OR Veps OR Yukaghirs OR Chulyms OR Evenks OR Tungus OR Evens OR "Kets" OR Khantys OR Mansi OR Vguls OR Selkups OR Teleuts OR Nanais OR Nanaitsy OR Negidal OR Nivikh OR Oroch OR orok OR Taz OR udege OR ulch OR Kumadins OR Chelkans OR Shorians OR Soyots OR Telengits OR Tofalars OR Tugalars OR "Tufans Todzhins" OR Laks OR Tabasarans OR Turuls OR Aguls OR Tsakhurs OR Kumyks OR Nogais OR "Andis" OR Akhvakh OR Archins OR Bagvalals OR Bezhta OR Botlikhs OR Chamalals OR Godoberi OR Hinukh OR Hunzibs OR Khwarshi OR Karata OR Tindis OR Tsez OR Abazin OR Besermyan OR Izhorians OR Karelians OR Nagaybaks OR Setos OR Shapsugs OR Quratay)) OR TS=((Greenlander OR "Kalaallit Nunaat" OR Nuuk OR Sisimiut OR Ilulissat OR "Qaqortoq Aasiaat" OR Maniitsoq OR Tasiilaq OR Uummannaq OR Narsaq OR Paamiut OR Nanortalik OR Upernavik OR Qasigiannguit)) OR TI=(("Abui" OR "Aeta" OR "Alfur" OR "Ati People" OR "Bahau" OR "Bali Aga" OR "Bali Mula" OR "Baliaga" OR "Balinese" OR "Basap" OR "Bataq" OR "Batek" OR "Bateq" OR "Batin" OR "Blaan" OR "Brao" OR "Bru" OR "Bru-Van Kieu" OR "Bugkalot" OR "Bumiputera" OR "Bunong" OR "Chams" OR "Champa" OR "Cheq Wong" OR "Dasun" OR "Dayak" OR "Deli Malay" OR "Filipina*" OR "Filiopino*" OR "Gaddang" OR "Ibaloi" OR "Iban" OR "Idan" OR "Igorot" OR "Jahai" OR "Jehai" OR "Jakun" OR "Javanese Kshatriya" OR "Kangeanese" OR " Kagayanen" OR "Kanayatn" OR "Katu" OR "Kayan" OR "Kavet" OR "Kenyah" OR "Khmer Krom" OR "Kimaragang" OR "Klemantan" OR "Kreung" OR "Kuijau" OR "Kuy" OR "Kwijau" OR "Lampung*" OR "Lanoh" OR "Lawangan" OR "Luangan" OR "Lumad" OR "Dayak" OR "Loloan Malay*" OR "Mah Meri" OR "Malay Singaporean*" OR "Malbog" OR "Maluku" OR "Mangka'ak" OR "Mangyan" OR "Maniq" OR "Maragang" OR "Ma'anyan" OR "Minokok" OR "Moluccas" OR Montagnard OR "Murut" OR "Naga" OR "Negrito" OR "New Guinea" OR "Ngaju" OR "Orang Hulu" OR "Orang Kanaq" OR "Orang Kuala" OR "Orang Seletar" OR "Orang Ulu" OR "Ot Danum" OR "Palawano" OR "Palembang*" OR " Panay-Bukidnon" OR "Pan-ayanon" OR "Papua" OR "Phnong" OR "Punong" OR "Pear People" OR "Por People" OR "Punan" OR "Rumanau" OR "Sasak" OR "Semai" OR "Semang" OR "Semaq Beri" OR "Semelai" OR "Senoi" OR "Simeulue" OR "Suludnon" OR "Sunda Island*" OR "Tabanwa" OR "Takua" OR "Tambanuo" OR " Tombonuo" OR "Taron" OR "Temiar*" OR "Temoq" OR "Temuan" OR "Tidung" OR "Trone" OR "Taaw't Bato") ) OR TI=(("pacific islander" OR Banabans OR "Native Hawaiian*" OR Guamanian OR Chamarro* OR Chuukese OR I-Kiribati OR Kosraean* OR Maohi OR Marshallese OR Moriori OR Mortlockese OR Mwokilese OR Namonuito OR Nauruans OR Ngatikese OR Niuean OR Paafang OR Paluans OR Pingelapese OR Pohnpeian* OR Pollapese OR Puluwat OR Rapanui OR Rotuman OR Satwalese OR Samoan*or Sonsorolese OR Tahitian* OR Tobian OR Tongan OR "Torres Strait Islander*" OR Tuvaluan OR Ulithian OR Woleian OR Yapese OR Polynesian* OR Micronesian* OR Melanesian*)) OR TI=(("Bedouin*" OR "Jahalin" OR "al-Kaabneh" OR "al-Azazmeh" OR "al-Ramadin" OR "al-Rshaida"))) AND ((TI=(((gyn?ecologic* OR rectal OR rectum OR genital OR prostat* OR unterine OR uterus OR ovarian OR endometrial) NEAR/3 (surgery OR surgical)) AND ("postoperative complication*" OR "post operative complication" OR "post surg* complication*")) OR AB=(((gyn?ecologic* OR rectal OR rectum OR genital OR prostat* OR unterine OR uterus OR ovarian OR endometrial) NEAR/3 (surgery OR surgical)) AND ("postoperative complication*" OR "post operative complication" OR "post surg* complication*"))) OR (TI=((prolaps* N/3 ("pelvic organ" OR uterine OR uterus OR bladder OR rectal OR rectum OR urethra* OR apical))) OR AB=((prolaps* N/3 ("pelvic organ" OR uterine OR uterus OR bladder OR rectal OR rectum OR urethra* OR apical)))) OR (TI=(((uterocele OR cystocele OR rectocele OR enterocele OR urethrocele OR sigmoidocele OR incontinence OR constipation OR encopresis OR "vaginal wind" OR "pelvic girdle pain" OR "neurogenic bladder*" OR "sexual dysfunction" OR "erectile dysfunction" OR "persistent genital arousal" OR impotence OR "bed wett*" OR bedwett* OR encopresis OR "hirshprung* disease" OR "gender confirmation surg*" OR "sex reassignment surg*"))) OR AB=(((uterocele OR cystocele OR rectocele OR enterocele OR urethrocele OR sigmoidocele OR incontinence OR constipation OR encopresis OR "vaginal wind" OR "pelvic girdle pain" OR "neurogenic bladder*" OR "sexual dysfunction" OR "erectile dysfunction" OR "persistent genital arousal" OR impotence OR "bed wett*" OR bedwett* OR encopresis OR "hirshprung* disease" OR "gender confirmation surg*" OR "sex reassignment surg*")))) OR (TI=((dyssynergia NEAR/3 (bowel* OR bladder*))) OR AB=((dyssynergia NEAR/3 (bowel* OR bladder*)))) OR (TI=(("lichen sclerosis" OR nocturia OR "lower urinary tract symptom*" OR dysuria OR "LUTS" OR "voiding dysfunction")) OR AB=(("lichen sclerosis" OR nocturia OR "lower urinary tract symptom*" OR dysuria OR "LUTS" OR "voiding dysfunction"))) OR (TI=(((fistula* OR fistulae) NEAR/3 (bladder* OR urinary OR vagina* OR rectovaginal* OR vesicovaginal*))) OR AB=(((fistula* OR fistulae) NEAR/3 (bladder* OR urinary OR vagina* OR rectovaginal* OR vesicovaginal*)))))

**SCOPUS June 20, 2023 Result =100**

( ( ( TITLE-ABS-KEY ( ( ( indige* OR aborig* OR nomadic OR seminomad* OR semi-nomad* ) W/3 ( people OR peoples OR elder OR elders OR grandmother* OR grandfather* OR parent* OR women* OR men OR woman OR man OR child* OR youth OR youths OR baby OR babies OR tribe OR tribes OR tribal OR shaman* OR native OR patient* ) ) AND ( middle-east OR afghanistan OR bahrain OR iran OR iraq OR israel OR ( jordan AND NOT jordan's-principle ) OR kuwait OR lebanon OR oman OR palestine OR qatar OR saudi-arabia ) ) ) OR ( TITLE-ABS-KEY ( ( saami OR sampi OR ( sami AND NOT ulus ) OR samis OR southernsami* OR umesami* OR pitesami* OR lulesami* OR northernsami* OR enaresami* OR kolasami* OR lapp OR lapps OR lappish OR lappland OR ( lapland* AND NOT longspur ) OR lappalainen* OR saamelainen* OR reindeer AND herd* OR reindeer AND culture* OR reindeer AND pastoral* OR lappbys OR samebys OR reinbeitesdistrikt OR paliskunta OR siida ) OR ( fennoscandia OR finnmark OR scandinavia OR nordic OR sweden OR norway OR finland OR swedish OR finnish OR norwegian OR norge OR svensk* OR suomi OR "Barents Region" OR ( kola AND NOT ( garcinia OR gotu ) ) OR "Arctic Europe*" OR "Polar Europe*" OR "North* Europ*" ) AND ( indigenous* OR ( traditional W/3 ( food* OR heal* OR medicine* OR shaman* ) ) ) OR ( indigenous* W/3 ( people* OR person* OR mother* OR father* OR parent* OR grandparent* OR grandmother* OR grandfather* OR elder OR elders OR child* OR boy OR boys OR girl* OR youth* OR healer* OR patient* OR famil* OR herder* OR village* OR communit* ) ) ) ) OR ( TITLE-ABS-KEY ( ( ( ( indigen* OR aborig* ) W/3 ( population* OR people OR peoples OR person OR persons OR elder OR elders OR man OR men OR woman OR women OR child* OR youth* OR clan OR clans OR tribe OR tribes OR tribal OR family OR families OR parent* OR grandparent* OR elder OR elders OR grandmother* OR grandfather* OR baby OR babies OR infant OR infants OR patient OR patients OR speakers OR speaking OR village* OR communit* ) ) ) AND ( ( ( micronesia OR "Caroline Island" OR guam OR kiribati OR "Mariana Island*" OR nauru OR palau OR melanesia OR "East Nusa Tenggara" OR fiji OR fijian OR "Loyalty Island*" OR "Maluku Islands" OR moluccas OR "New Caledonia*" OR "New Guinea" OR papua OR rotuma OR "Solomon Island*" OR vanuatu OR polynesia OR "Austral Island*" OR "Cook Island*" OR "Chatham Islands" OR "Easter Island*" OR hawaii* OR "Marquesas Island" OR "Norfolk Island" OR samoa OR tahiti OR tokelau OR tonga OR tuvalu OR tuamotu OR kermadec OR banabans OR "Native Hawaiian*" OR guamanian OR chamarro* OR chuukese OR i-kiribati OR kosraean* OR maohi OR marshallese OR moriori OR mortlockese OR mwokilese OR namonuito OR nauruans OR ngatikese OR niuean OR paafang OR paluans OR pingelapese OR pohnpeian* OR pollapese OR puluwat OR rapanui OR rotuman OR satwalese OR samoan* OR sonsorolese OR tahitian* OR tobian OR tongan OR "Torres Strait Islander*" OR tuvaluan OR ulithian OR woleian OR yapese OR polynesian* OR micronesian* OR melanesian* ) ) ) ) ) OR ( TITLE-ABS-KEY ( ( ( ainus OR ainu OR aleuts OR alyutors OR chukchis OR chuvans OR dolgans OR enets OR entsy OR yupik OR yup'ik OR yuit OR yupigyt OR chaplino OR naukan OR itelmens OR kamchadals OR kereks OR "Komi" OR koryaks OR nenets OR nentsy OR nganasans OR tavgi OR sami OR veps OR yukaghirs OR chulyms OR evenks OR tungus OR evens OR "Kets" OR khantys OR mansi OR vguls OR selkups OR teleuts OR nanais OR nanaitsy OR negidal OR nivikh OR oroch OR orok OR taz OR udege OR ulch OR kumadins OR chelkans OR shorians OR soyots OR telengits OR tofalars OR tugalars OR "Tufans" OR "Todzhins" OR laks OR tabasarans OR turuls OR aguls OR tsakhurs OR kumyks OR nogais OR "Andis" OR akhvakh OR archins OR bagvalals OR bezhta OR botlikhs OR chamalals OR godoberi OR hinukh OR hunzibs OR khwarshi OR karata OR tindis OR tsez OR abazin OR besermyan OR izhorians OR karelians OR nagaybaks OR setos OR shapsugs OR quratay ) OR ( ( ( indigenous OR aborig* OR tribe OR tribal OR tribes ) W/3 ( population* OR people* OR person* OR elder* OR man OR men OR woman OR women OR child* OR youth* OR clan OR clans OR tribe OR tribes OR tribal OR family OR families OR parent* OR grandparent* OR elder OR elders OR grandmother* OR grandfather* OR baby OR babies OR infant OR infants OR patient OR patients OR speakers OR speaking OR village* OR communit* ) ) ) ) AND ( russia* ) ) ) OR ( TITLE-ABS-KEY ( ( abipon OR achuar OR achuagua OR akawaio OR amarizana OR andoque OR akawaio OR akuriyo OR anauya OR araona OR arawak OR ayamn OR aguaruna OR amahuaca OR amarakaeri OR andoa OR arabela OR arawak OR arhuaco OR ashaninca OR asheninca OR atsahuaca OR aymara OR ayoreo OR bakairi OR "Baniva" OR barasana OR baniwa OR "Baure" OR bororo OR cabiyari OR cacataibo OR caquinte OR cacua OR cahuarano OR "Caiua" OR "Camara Indians" OR camaracoto OR camsa OR canamari OR candoshi OR canela OR canichana OR capanahua OR carapana OR cariay OR "Carib" OR carijona OR carutana OR cashibo OR cashinahua OR cawishana OR cavinena OR caxuiana OR cayuvava OR chontaquiro OR cocama OR "Cubeo" OR curipaco OR chacobo OR chaima OR ( chana AND NOT striatus ) OR chapacura OR charrua OR chimila OR chitonahua OR chorote OR chipaya OR chiquitano OR chulupi OR carare OR coconuco OR cofan OR coreguaje OR coyaima OR chamacoco OR chamicuro OR chayahuita OR cocama OR "Culina" OR culino OR cubeo OR cuiba OR "Cuiva" OR cumanagoto OR curripaco OR "Deni" OR desano OR embera OR guarani OR guajajara OR "Guana" OR guanano OR guarayo OR guarayu OR guahibo OR guajiro OR guambiano OR guanano OR guayabero OR guarequena OR guinao OR "Guana" OR gayon OR guahibo OR hixkaryana OR huachipairi OR huambisa OR huarayo OR lauanaua OR ikpeng OR ingariko OR irantxe OR itonama OR inapari OR iquito OR isconahua OR jumana OR japreria OR jirajara OR juruti OR jaqaru OR jebero OR kadiweu OR kaingang OR kamayura OR karaja OR karipuna OR "Kariri" OR katukina OR kaxarari OR kayabi OR kayapo OR "Kuikuro alapalo" OR kulina OR "Kaiwa" OR kallawaya OR "Kogui" OR "Kuna" OR kaweskar OR "Lule" OR macuna OR maipure OR mapuche OR mataco OR mocovi OR machinere OR machinerev OR machiguenga OR macushi OR macuna OR "Madi" OR malayo OR mamainde OR manao OR mandauaca OR mandawaka OR mapidian OR mapuche OR mapidian OR maquiritare OR maquiritari OR maragua OR marawan OR mariate OR marubo OR mastanahua OR mataco OR matipuhy OR "Matis" OR "Matses" OR mawakua OR mawakwa OR maxakali OR mehinaku OR miranha OR moronawa OR munduruku OR movima OR muellama OR muinane OR mapoyo OR "Mashco Piro" OR muniche OR nambikwara OR nocaman OR nuquini OR nomatsiguenga OR nanti OR ocaina OR omagua OR orejon OR "Opon" OR pacahuara OR "Paez" OR paicone OR palicur OR panare OR "Pano" OR "Paresi" OR paumari OR "Pemon" OR pilaga OR puelche OR pauna OR pauserna OR piapoco OR piraha OR piratapuyo OR pisabo OR piaroa OR "Pijao" OR piratapuyo OR paraujano OR pemon OR pemono OR piapoco OR puinave OR patamona OR poyanawa OR puinave OR puquina OR quechua OR quichua OR retuara OR resigaro OR reyesano OR sabanes OR "Saliba" OR saluma OR sarave OR secoya OR selknam OR "Sensi" OR shaninawa OR shapra OR sharanahua OR shebayo OR shiwiar OR shikiana OR sikiana OR siriono OR sinsiga OR "Siona" OR suruwaha OR tacano OR tamanaco OR tiahuanaco OR tariano OR tehuelche OR tariano OR tatuyo OR "Tembe" OR "Terena" OR telembi OR ticuna OR ticuna OR tiriyo OR tiwanaku OR tiwanaku OR "Torom" OR "Totoro" OR tsimane OR tuberao OR "Tucano" OR tunebo OR tuxinawa OR tuyuca OR uainuma OR urarina OR vilela OR waimaha OR waiampi OR waiwai OR wapishana OR waraiku OR warekena OR waura OR wayampi OR wayana OR wirina OR waimaha OR waunana OR "Wiwa" OR "Warao" OR wayuu OR witoto OR xavante OR xipaya OR xiriana OR xokleng OR yabaana OR yaminawa OR yaminahua OR yaruma OR yawalapiti OR yuracare OR yabarana OR yavitero OR "Yine" OR yamana OR yaghan OR yucuna OR yurumangui OR yukpa OR yanesha OR yoranahua OR yagua OR yaminahua OR zaparo OR zamuco OR "Trio Indians" OR "More Indians" OR "Bare Indians" ) OR ( ( "Inga" OR "Maca" OR "Leco" OR "Mojo" OR "Uro" OR "Maco" OR lengua OR "Toba" OR "Zoe" OR "Ona" OR "Catio" OR "Passe" OR "Bari" OR "Awa" OR "Bora" OR "Bara" OR "Remo" OR "Pano" OR "Sape" ) W/3 ( indians OR indian OR indigenous OR amerindian* OR aborigin* OR people OR peoples OR elder OR elders OR grandmother* OR grandfather* OR parent* OR women OR men OR woman OR man OR child* OR youth OR youths OR baby OR babies OR tribe OR tribes OR tribal OR shaman* OR native OR patient* ) ) OR ( ( indian* OR amerindian, OR aboriginal* OR indigenas OR indigenous OR tribe OR tribes OR tribal ) AND ( "South America*" OR argentin* OR bolivia* OR brazil* OR chile* OR colombia* OR ecuador* OR "French Guiana" OR guyana* OR paraguay* OR peru OR peruvian OR suriname OR uruguay* OR venezuela* OR "Amazon Region" OR amazonia OR andes OR andean ) ) ) ) OR ( ( TITLE-ABS-KEY ( ( ( "Bali" OR "Borneo" OR "Brunei" OR cambodia OR {Flores Island} OR "Indonesia" OR java OR komodo OR "Laos" OR "Lombok" OR "Malasia" OR {Malay Peninsula} OR "Myanmar" OR {Mekong Valley} OR {New Guinea} OR "Philippeans" OR "Sabah" OR "Singapore" OR "Sumatra" OR "Sumba" OR "Sumbawa" OR "Thailand" OR "Timor" OR "Vietnam" ) ) AND ( ( indigen* OR aborig* OR tribe OR tribes OR tribal ) W/3 ( population* OR people OR peoples OR person OR persons OR elder OR elders OR man OR men OR woman OR women OR child* OR youth* OR clan OR clans OR family OR families OR parent* OR grandparent* OR elder OR elders OR grandmother* OR grandfather* OR baby OR babies OR infant OR infants OR patient OR patients OR speakers OR speaking OR village* OR communit* ) ) ) ) OR ( TITLE-ABS-KEY ( ( "Abui" OR "Aeta" OR "Alfur" OR "Ati People" OR "Bahau" OR "Bali Aga" OR "Bali Mula" OR "Baliaga" OR "Balinese" OR "Basap" OR "Bataq" OR "Batek" OR "Bateq" OR "Batin" OR "Blaan" OR "Brao" OR "Bru" OR {Bru-Van Kieu} OR "Bugkalot" OR "Bumiputera" OR "Bunong" OR "Chams" OR "Champa" OR {Cheq Wong} OR "Dasun" OR "Dayak" OR "Deli Malay" OR "Filipina*" OR "Filiopino*" OR "Gaddang" OR "Ibaloi" OR "Iban" OR "Idan" OR "Igorot" OR "Jahai" OR "Jehai" OR "Jakun" OR {Javanese Kshatriya} OR "Kangeanese" OR " Kagayanen" OR "Kanayatn" OR "Katu" OR "Kayan" OR "Kavet" OR "Kenyah" OR {Khmer Krom} OR "Kimaragang" OR "Klemantan" OR "Kreung" OR "Kuijau" OR "Kuy" OR "Kwijau" OR "Lampung*" OR "Lanoh" OR "Lawangan" OR "Luangan" OR "Lumad" OR "Dayak" OR {Loloan Malay*} OR {Mah Meri} OR {Malay Singaporean*} OR "Malbog" OR "Maluku" OR "Mangka'ak" OR "Mangyan" OR "Maniq" OR "Maragang" OR "Ma'anyan" OR "Minokok" OR "Moluccas" OR montagnard OR "Murut" OR "Naga" OR "Negrito" OR {New Guinea} OR "Ngaju" OR {Orang Hulu} OR {Orang Kanaq} OR {Orang Kuala} OR {Orang Seletar} OR {Orang Ulu} OR {Ot Danum} OR "Palawano" OR "Palembang*" OR {Panay-Bukidnon} OR {Pan-ayanon} OR "Papua" OR "Phnong" OR "Punong" OR {Pear People} OR {Por People} OR "Punan" OR "Rumanau" OR "Sasak" OR "Semai" OR "Semang" OR {Semaq Beri} OR "Semelai" OR "Senoi" OR "Simeulue" OR "Suludnon" OR {Sunda Island*} OR "Tabanwa" OR "Takua" OR "Tambanuo" OR " Tombonuo" OR "Taron" OR "Temiar*" OR "Temoq" OR "Temuan" OR "Tidung" OR "Trone" OR {Taaw't Bato} ) ) ) ) OR ( TITLE-ABS-KEY ( ( "A' ani" OR absaroka OR haaninin OR atsina OR "Gros Ventre" OR acopsel OR tlacopsel OR lacopsel OR ahtna OR ahtena OR akenitsi OR occaneechi OR akokisa OR horcoquisa OR orcoquizas OR aleut OR unangax OR unangan OR alibamu OR "Alabama Alsea" OR alutiiq OR sugpiag OR amahami OR awaxawi OR androscoggin OR arosaguntacook OR ameriscoggin OR anishinaabeg OR chippewa OR anihsinape OR saulteaux OR apalachee OR aranama OR "Texan Coahuilteca" OR tamique OR arikara OR sahnish OR arickaree OR adakadaho OR assiniboine OR hohe OR nakota OR nakoda OR nakona OR "Atsa' Kudok-wa" OR awatixa OR bannock OR "Snake Indian*" OR bidai OR quasmigdo OR biloxi OR blackfoot OR niitsitapi OR sikasikaitsitapi OR cahto OR kaipomo OR cahuilla OR ivilyuqaletem OR ivilyuat OR catawba OR inna OR iswa OR chemehuevi OR chickasaw OR "Chilula Chimakum" OR aqokulo OR chimariko OR chiricahua OR tsokanende OR chitimacha OR chetimachan OR sitimacha OR chowanoke OR roanoke OR chumash OR ciboney OR "Taino Ciwat" OR clatsop OR coos OR coosa OR uchis OR chiaha OR coste OR talisi OR coquille OR kokwell OR coso OR cowlitz OR taitnapam OR "Crow Nation" OR "Cui Ui Ticutta" OR cupeno OR kuupangaxwichem OR cupa OR "Cup' ig" OR nunivak OR "Dakota Oyate" OR lakota OR nakota OR santee OR teton OR sioux OR deadose OR "Deg Xina" OR "Deg Xit' an" OR kaiyuhkhotana OR "Deg Hit' an" OR "Dena' ina" OR tanaina OR "Dichinanek' Hwt' ana" OR "Upper Kuskokwim Athabascan*" OR kolchan OR goltsan OR "Tundra Kolosh" OR "Do lkabya" OR duwamish OR esselen OR eyak OR "Gidi' tikadi" OR guwevkabaya OR "Gwich' in" OR kutchin OR haida OR xaadas OR xaat OR halchidhoma OR havasupai OR "Green Water People" OR hiratsa OR hiraaca OR "Ho-chaaqa" OR winnebago OR holikachuk OR innoko OR "Tlegon-khotana" OR hopi OR "Houma-Louisiana" OR huaco OR waco OR hualapai OR hupa OR natinixwe OR "Natinook-wa" OR "Hwech' in" OR hankutchin OR "Iroquois Confederacy" OR "Hodinoso ni" OR "Illinois Confedera*" OR ilinoweg OR illini OR inupiat OR inuit OR ioway OR baxoje OR jicarilla OR juaneno OR acjachemen OR jumano OR kalapuya OR clackama OR kalispel OR "Pend d' Oreilles" OR qlispe OR karuk OR karok OR "Chum-ne" OR katkoc OR kansa OR kanza OR kawaiisu OR nuwa OR kennebec OR "Kinipekw Kittitas" OR klickitat OR "Qwu' lh-hwai-pum" OR "Awi-adshi" OR mahane OR wahnookt OR "Koa' aga' itoka" OR keresan OR kichai OR kitsai OR keechi OR "K' itaish" OR kiowa OR gaigwu OR cauigu OR kutjau OR "Kwu-da" OR "Tep-da" OR kitanemuk OR kittitas OR klickitat OR "Qwu' lh-hwai-pum" OR "Awi-adshi" OR mahane OR wahnookt OR "Koa' aga' itoka" OR konkow OR "Koop Ticutta" OR koyukon OR ktunaxa OR kootenai OR flathead OR kucadikadi OR "Kotsa' va" OR kumeyaay OR "Tipai-Ipai" OR kamia OR diegueno OR kwapa OR cocopah OR cucapa OR "Xawitt kwnchawaay" OR lassik OR lenape OR "Leni-Lenape" OR lipan OR luiseno OR payomkawichum OR madqwadabaya OR "Desert Yavapai" OR mahican OR mohicans OR makah OR makuhadokado OR maliseet OR wolistoqiag OR manahoac OR mahock OR meipontsky OR mandan OR mattole OR "Bear River" OR "Tul' bush" OR "Ni' ekeni" OR meherrin OR menominee OR mackinac OR mescalero OR myaamiaki OR kickapoo OR twigtwee OR missouria OR miwok OR miwuk OR moadokado OR modoc OR mohave OR "Aha Makhav" OR mohawk OR "Kaneng' hega" OR molala OR molale OR molele OR nyyhmy OR moosonee OR "Moose Cree" OR monsonis OR multnomah OR chinook OR nabedache OR nabaydacu OR wawadishe OR nabiltse OR dakubetede OR "Nacho Nyak Dun" OR tutchone OR nacono OR "Na' isha" OR nanticoke OR navajo OR ndee OR nial OR niimiipu OR "Nez Perce" OR watapala OR watapahlute OR nisenan OR nisqually OR nomlaki OR noamlakee OR "Central Wintun" OR nongatl OR nottoway OR cheroenhaka OR "Northern Cheyenne" OR ohlone OR costanoan OR omaha OR "O' odham" OR pima OR papago OR osage OR otoe OR otse OR "Ozav Dika" OR palus OR passamaquoddy OR pestomuhkati OR patiri OR petaros OR pastia OR patwin OR "Southern Wintun" OR panis OR skidi OR pedee OR penobscot OR "Petun Piipaash" OR "Kokmalik' op" OR piscatawa OR doeg OR conoy OR "Pit River" OR pomo OR kashaya OR ponca OR ponka OR pottawatomi OR bodewadmik OR powhatan OR puyallup OR spuyalepabs OR quapaw OR ugahxpa OR quechan OR yuma OR kwtsaan OR quileute OR salinan OR saponi OR monacan OR sapon OR "Eastern Blackfoot" OR christanna OR sawawatodo OR serrano OR taaqtam OR "Maarenga' yam" OR yuhaviatam OR shasta OR chasta OR sasti OR shoshone OR siletz OR sinkine OR sinkyone OR "Siuslaw Umpqua" OR skitswish OR "Schitsu' umash" OR snohomish OR snuqualmi OR sokoki OR missiquoi OR stillaguamish OR stoluckwamish OR suquamish OR sutaio OR swinomish OR skagit OR syilx OR okanagan OR sotaae OR "Taga Ticutta" OR takelma OR dagelma OR taltushtuntede OR galice OR taos OR taovaya OR tataviam OR alliklik OR tawakoni OR tahuacano OR tenino OR thawikila OR hathawekela OR "Fort Ancient" OR tigua OR tillamook OR nehalem OR timbisha OR panamint OR timpanogos OR tlingit OR "Toi Ticutta" OR tolowa OR "Talawa Dini'" OR tongva OR gabrieleno OR fernandeno OR tobikhar OR tonkawa OR ticanwatic OR tsikip OR appalousa OR opelousa OR tsitsistas OR tubatulabal OR tukabatchee OR tuscarora OR tomahittan OR kuskarawock OR tutelo OR tutero OR totteroy OR tutera OR yusan OR tututni OR umatilla OR umpqua OR waccamaw OR waxmaw OR wadatika OR "Harney Valley Paiute" OR wailiki OR waluulapam OR walpapi OR huipui OR wampanoag OR massasoit OR wanapum OR wappo OR washoe OR wichita OR willapa OR kwalhioqua OR "Wi pukba" OR "Verde Valley Yavapai" OR wintu OR "Northern Wintun" OR wiyot OR "Wee' at" OR weyet OR yakama OR "Yamosopo Tuviwarai" OR yaqui OR yoeme OR yatasi OR yattasih OR "Yavbe'" OR "Yavapai" OR "Ysleta del Sur" OR yojuane OR yokuts OR mariposa OR yuki OR yupighyt OR "Yup'ik" OR yupik OR yurok OR "Olekwo'l" OR zuni ) ) OR ( ( applegate OR delaware OR iowa OR ishak OR kaw OR kato OR spokane OR miami OR arkansas OR tali OR tunica OR han OR pawnee OR "Coeur D' Alene" OR piscataway OR ree OR tula OR "Walla Walla" ) W/3 ( reservation* OR nation OR people OR peoples OR population OR man OR men OR woman OR women OR child* OR youth* OR elder OR elders OR communit* OR tribe OR tribes OR tribal OR indian* ) ) ) OR ( TITLE-ABS-KEY ( ( ( ( acatec OR aguacateco OR amuzgo OR bokota OR boruca OR bribri OR "Bri Bri" OR buglere OR cabecar OR cakchiquel OR changuena OR chatino OR chiapanec OR chicomuceltec OR chinantee OR chocho OR cholti OR "Ch'olti'" OR "Ch'olti'an" OR chontal OR chorotega OR chorti OR chuj OR chumbia OR corobici OR ( cueva AND NOT spain ) OR cuicatec OR cuitlatee OR cuytec OR dorasque OR embera OR garifuna OR guatuso OR guaymi OR guaymis OR guetar OR huastec OR huave OR huetar OR itzaj OR ixil OR jacalteco OR jonaz OR kanjobal OR kekchi OR kuna OR maleku OR mangue OR matambu OR matlatzinca OR mazahua OR motozintlec OR mayan OR mayangna OR miskito OR mixtec OR mopan OR nahua OR nahuatl OR ngabe OR otomi OR pantec OR paya OR popoloca OR popoloc OR poqomam OR poqomchi OR "Q'eqchi'" OR quiche OR quitirrisi OR sacapulteco OR sipacapense OR subtiaba OR tacaneco OR tarasco OR tamaulipec OR tamazultec OR tecoxquin OR tectiteco OR tecual OR tecuexe OR tepehura OR tepuztecor OR teribe OR terraba OR totonac OR trique OR tzeltal OR tzotzil OR tzutujil OR ulwa OR uspantec* OR uspanteko OR voto OR xinca OR waunana OR wounaan OR yucatec OR zapotec OR zoque OR ( ( "Costa Rica*" OR hondura* OR nicaragua* OR panama* OR guatemala* OR achi OR belize OR belizean* OR maya* OR mixe OR pame OR pipil OR pech OR chol OR "Ch'olan" OR cora OR cuna OR mam OR rama ) W/5 ( indian OR indians OR amerindian* OR amerindio* OR aboriginal* OR indigenous OR indigena* OR aborigen* OR mestizo OR tribe OR tribes OR tribal OR "traditional medicine*" OR shaman* ) ) ) OR ( belize OR "Costa Rica" OR "El Salvador" OR guatemala OR honduras OR nicaragua OR panama ) ) AND ( ( indigenous W/3 ( population* OR people* OR person* OR elder* OR man OR men OR woman OR women OR child* OR youth* OR clan OR clans OR tribe OR tribes OR tribal OR family OR families OR parent* OR grandparent* OR elder OR elders OR grandmother* OR grandfather* OR baby OR babies OR infant OR infants OR patient OR patients OR speakers OR speaking OR village* OR communit* ) ) ) ) AND NOT ( ( mexico OR mexican ) ) ) ) ) OR ( ( ( TITLE-ABS-KEY ( ( greenland* OR "Kalaallit Nunaat" OR nuuk OR sisimiut OR ilulissat OR qaqortoq AND aasiaat OR maniitsoq OR tasiilaq OR uummannaq OR narsaq OR paamiut OR nanortalik OR upernavik OR qasigiannguit ) AND ( inuit OR inuk OR ( ( indigen* OR aborigin* ) W/3 ( population* OR people OR peoples OR person OR persons OR elder OR elders OR man OR men OR woman OR women OR child* OR youth* OR clan OR clans OR tribe OR tribes OR tribal OR family OR families OR parent* OR grandparent* OR elder OR elders OR grandmother* OR grandfather* OR baby OR babies OR infant OR infants OR patient OR patients OR speakers OR speaking OR village* OR communit* ) ) ) OR ( greenlandic OR kalaallit OR kalaallisut OR tunumiit OR inughuit OR avanersuarmiut ) ) ) OR ( TITLE-ABS-KEY ( ( ( india OR bangladesh* OR ( bhutan* AND NOT bhutanensis ) OR ( nepal* AND NOT nepalensis ) OR pakistan* OR "Sri Lanka*" ) ) AND ( ( ( indigenous* OR tribe OR tribal OR tribes ) W/3 ( population* OR people* OR person* OR elder* OR man OR men OR woman OR women OR child* OR youth* OR clan OR clans OR tribe OR tribes OR tribal OR family OR families OR parent* OR grandparent* OR elder OR elders OR grandmother* OR grandfather* OR baby OR babies OR infant OR infants OR patient OR patients OR speakers OR speaking OR village* OR communit* ) ) ) OR "Adivasis" OR "Adnamanese" OR "Andaman" OR "Baluch" OR "Baluchis" OR "Bodo" OR "Boro" OR "Boros" OR "Bote" OR "Brahuis" OR "Chakmas" OR "Chepang" OR "Chhantyal" OR "Damai" OR "Dewan" OR "Ghale" OR "Gurkha" OR "Gurung" OR "Hayu" OR "Hyolmo" OR "Jarawa" OR "Jirel" OR "Jumma" OR "Kalash" OR "Khas" OR "Kirati" OR "Koinch" OR "Kulung" OR "Kusunda" OR "Limbu" OR "Lohorung" OR "Magar" OR "Makrani" OR "Mangar" OR "Marma" OR "Miji" OR "Mongar" OR "Mro" OR "Naga" OR "Nepami" OR "Newar" OR "Nicobar" OR "Onge" OR "Rai" OR "Rang" OR "Raute" OR "Sajolang" OR "Santhal" OR "Sentinelese" OR "Sindhis" OR "Sulemani" OR "Sunuwar" OR "Tamang" OR "Thakali" OR "Thangmi" OR "Tharu" OR "Tripura" OR "Tumbahangphe" OR "Wanniyala-Aetto" OR "Yakkha" OR "Yolmopa" ) ) OR ( TITLE-ABS-KEY ( ( ( ( ( mexico AND NOT {New Mexico} ) OR mexican OR aguascalientes OR {Baja California} OR campeche OR chiapas OR ( chihuahua AND NOT ( dog OR dogs OR pet OR pets ) ) OR coahuila OR colima OR durango OR guanajuato OR guerrero OR hidalgo OR jalisco OR michoacan OR morelos OR nayarit OR {Nuevo Leon} OR oaxaca OR puebla OR queretaro OR {Quintana Roo} OR {San Luis Potosi} OR sinaloa OR sonora OR ( tabasco AND NOT ( sauce* OR flavo* ) ) OR tamaulipas OR tlaxcala OR veracruz OR yucatan OR zatecas ) ) AND ( ( mesoamerindian* OR indigen* OR aborig* OR {first people*} OR {indos mexicano} OR {original people*} OR {pueblos indigenas} ) ) ) OR ( aguacatec OR akwa'ala OR abxubal OR ayuukja'ay OR {Batzil k'op} OR binizaa OR {Chichimeca Jonaz} OR chinantec OR chocho OR {Ch ol} OR chontal OR "Chuj" OR cochimi OR comcaac OR hamasipini OR harijio OR {Ha shuta enima} OR {Hach t'an} OR huastecor AND hnahnu OR hnatho OR ixcatec OR ixil OR jacaltec OR {K'akchikel} OR {K'anjobal} OR kanjobal OR kaqchikel OR kechi OR {K'iche} OR kikapooa OR kikapu OR kiliwa OR {Ko'lew} OR {K'op o winik atel} OR kumiai OR lacandon OR laymon OR makurawe OR maya OR {Maya'wiinik} OR mazahua OR mazatec OR me'phaa OR mexicanero OR mexikatlajtolli OR "Mixe" OR mixtec OR motocintleco OR {Mti'pa} OR nahuas OR ocuiltec OR otomi OR ( oaxaca AND NOT oaxaca-blinder ) OR "Pame" OR papago OR tlahuica OR paipai OR {Pima Bajo} OR purepecha OR {P'urhepecha} OR qatok OR ( quiche AND NOT guatemala ) OR {Q'iche} OR raramuri OR {Runixa ngiigua} OR ( "Seri" AND NOT {Seri 82} ) OR {Slijuala sihanuk} OR tacuate OR tarahumara OR teenek OR tepehua OR ti'pai OR tlapanec OR {Tohono O'odham} OR totonac OR tachiwin OR {Tsa jujmi} OR tzotzil OR {Tu'un savi} OR tzeltal OR "Uza" OR "Winik" OR xigue OR yucatec OR zapotec ) ) ) ) OR ( ( ( TITLE-ABS-KEY ( ( "Indigenous" OR "Afar" OR "Afars" OR "Aka" OR "Akie" OR "Ameru" OR "Asua" OR "Ateker" OR "Atwot" OR "Awjila" OR "Bafia" OR "Baka" OR "Bakongo" OR "Bakwe" OR "Balunda" OR "Balovale" OR "Bango" OR "Bassa" OR "Beja" OR "Bekpak" OR "Bemba" OR "Bembe" OR "Benet" OR "Berber" OR "Berbers" OR "Bira" OR "Bowe" OR "Bubi" OR "Budja" OR "Bulu" OR "Bunrun" OR "Chaga" OR "Chopi" OR "Damara" OR "Dinka" OR "Djerba" OR "Duala" OR "Dzing" OR "Efe" OR "Elmolo" OR "Fang" OR "Foora" OR "Fula" OR "Fur" OR "Ghomara" OR "Ghadames" OR "Gllana" OR "Glu" OR "Gogo" OR "Gongo" OR "Haya" OR "Havu" OR "Hema" OR "Hima" OR "Hunde" OR "Hutu" OR "Huva" OR "Iboko" OR "Igbo" OR "Ijo" OR "Jieng" OR "Kadu" OR "Kande" OR "Kango" OR "Katla" OR "Kgaga" OR "Khoe" OR "Kola" OR "Komo" OR "Kota" OR "Kua" OR "Kuba" OR "Kwango" OR "Kx'z" OR "Kxoe" OR "Lala" OR "Lozi" OR "Luo" OR "Luba" OR "Lupu" OR "Masmuda" OR "Matmata" OR "Mbala" OR "Mbam" OR "Mbo" OR "Mbolo" OR "Mbuza" OR "Mongo" OR "Mpondo" OR "Myene" OR "Naadh" OR "Nama" OR "Nande" OR "Naro" OR "Ngoni" OR "Ndau" OR "Ndebele" OR "Ngoli" OR "Ngondi" OR "Ngoni" OR "Nguni" OR "Nkoya" OR "Nkumu" OR "Nuba" OR "Nubian" OR "Nuer" OR "Nzebi" OR "Ogoni" OR "Omoro" OR "Oroko" OR "Pygmy" OR "Popoi" OR "Poto" OR "Puru" OR "Rashad" OR ( san AND NOT ( "San Francisco" OR "San Diego" OR "San Antonio" ) ) OR "Sango" OR "Sanhaja" OR "Sena" OR "Shilha" OR "Shira" OR "Shona" OR "Shua" OR "Sokna" OR "Somali*" OR "Sotho" OR "Sua" OR "Subu" OR "Swazi" OR "Taitaa" OR "Tchokwe" OR "Teke" OR "Tembo" OR "Tetela" OR ( tonga AND africa* ) OR "Tshwa" OR "Tsoa" OR "Twa" OR "Turkana" OR "Tuu" OR "Venda" OR "Vira" OR "Watta" OR "Wakuti" OR "Yaaku" OR "Yaka" OR "Yakoma" OR "Yanzi" OR "Yao" OR "Yeke" OR "Yela" OR "Yeyi" OR "Zulu" ) W/3 ( population* OR people OR peoples OR person OR persons OR elder OR elders OR man OR men OR woman OR women OR child* OR youth* OR clan OR clans OR tribe OR tribes OR tribal OR family OR families OR parent* OR grandparent* OR elder OR elders OR grandmother* OR grandfather* OR baby OR babies OR infant OR infants OR patient OR patients OR speakers OR speaking OR village* OR communit* ) ) ) OR ( TITLE-ABS-KEY ( "Abakuria" OR "Abaluhya" OR "Abagusii" OR "Abakuria" OR "Aembu" OR "Agikuyu" OR "Akamba" OR "Anuak" OR "Anywaa" OR "Amazigh" OR "Ambala" OR "Ambeere" OR "Ambundu" OR "Ambuun" OR "Amharan" OR "Angba" OR "Baaka" OR "Baamba" OR "Babindi" OR "Babini" OR "Baboma" OR "Bachokwe" OR "Bacwa" OR "Bafumbira" OR "Baganda" OR "Bagyele" OR "Bagwere" OR "Bagyeli" OR "Bakiga" OR "Bakola" OR "Baholo" OR "Bakalanga" OR "Bakiga" OR "Bakolo" OR "Bakongo" OR "Bakonjo" OR "Baluba" OR "Balunda" OR "Balovale" OR "Bamasaba" OR "Bambuti" OR "Bangala" OR "Bangoli" OR "Bangungu" OR "Bantu" OR "Banyankole" OR "Banyarwanda" OR "Banyole" OR "Banyoro" OR "Bapende" OR "Bapedi" OR "Barabaig" OR "Barombi" OR "Barundi" OR "Baruuli" OR "Basamia" OR "Basoga" OR "Batswana" OR "Batooro" OR "Batsamba" OR "Batswana" OR "Batwa" OR "Bayaka" OR "Bedzan" OR "Bazombe" OR "Bebayaka" OR "Bedzan" OR "Bhaca" OR "Biaka" OR "Borana" OR "Chewa" OR "Copts" OR "Cormorian" OR "Cushitic" OR "Dahalo" OR "Datooga" OR "Dikidiki" OR "Dogon" OR "Ewondo" OR "Fulani" OR "Fuliru" OR "Ganguela" OR "Gciriku" OR "Gyele" OR "Hadza" OR "Hadzabe" OR "Haillom" OR "Haratin" OR "Herero" OR "Himba" OR "Hlubi" OR "Iriryen" OR "Iqvayliyen" OR "Kabyle*" OR "Kalenjin" OR "Kanioka" OR "Kanioka" OR "Kaonde" OR "Karamojong" OR "Kavango" OR "Kereuyu" OR "Khoikhoi" OR "KhoiSan" OR "Kikuyu Kwangali" OR "Lokele" OR "Lowme" OR "Lotuko" OR "Lwalwa" OR "Maasai" OR "Makonde" OR "Makua" OR "Mande" OR "Masalit" OR "Matumbi" OR "Mayeuyi" OR "Mayeyi" OR "Mbenga" OR "Mbukushu" OR "Mbochi" OR "Mboro" OR "Mbuti" OR "Medzan" OR "Mijikenda" OR "Mozabite*" OR "Nafusa" OR "Ndebele" OR "Ngombe" OR "Namaqua" OR "Nyanga" OR "Nyamwezi" OR "Ogiek" OR "Ovambo" OR "Ovimbundu" OR "Phuthi" OR "Pokomo" OR "Rendille" OR "Riffian" OR "Riffians" OR "Sakuma" OR "Samburu" OR "Sandawe" OR "Sangha" OR "Sango" OR "Sengwer" OR "Serer" OR "Sesotho" OR "Shangaan" OR "Shawiya" OR "Shenwa" OR "Shi" OR "Shilluk" OR "Sukua" OR "Sukus" OR "Swahili" OR "Tabwa" OR "Tambuka" OR "Taveta" OR "Thembu" OR "Tigrayan" OR "Topoke" OR "Tsonga" OR "Toubou" OR "Tuareg" OR "Tumbuka" OR "Ugana" OR "Wochua" OR "Xhosa" OR "Xindonga" OR "Yoruba" OR "Zenati" OR "Zuwara" ) ) ) OR ( TITLE-ABS-KEY ( aborigine OR "torres strait islander*" ) OR TITLE-ABS-KEY ( ( australia* OR northern AND territory OR tasmania OR new AND south AND wales OR victoria OR queensland ) AND ( ( tribe OR tribes OR tribal OR indig* ) W/3 ( population* OR people OR peoples OR person OR persons OR elder OR elders OR man OR men OR woman OR women OR child* OR youth* OR clan OR clans OR family OR families OR parent* OR grandparent* OR elder OR elders OR grandmother* OR grandfather* OR baby OR babies OR infant OR infants OR patient OR patients OR speakers OR speaking OR village* OR communit* ) ) ) ) OR ( TITLE-ABS-KEY ( ( maori OR "tangata whenua" ) OR ( ( ( new AND zealand OR aukland OR aotearoa ) ) AND ( ( aborig* OR indig* ) ) ) ) ) OR ( TITLE-ABS-KEY ( athapaskan OR saulteaux OR wakashan OR cree OR dene OR inuit OR inuk OR inuvialuit* OR haida OR ktunaxa OR tsimshian OR gitsxan OR {Nisga'a} OR haisla OR heiltsuk OR oweenkeno OR {Kwakwaka'wakw} OR {Nuu chah nulth} OR {Tsilhqot'in} OR dakelh OR {Wet'suwet'en} OR sekani OR {dunne-za} OR dene OR tahltan OR kaska OR tagish OR tutchone OR nuxalk OR salish OR {stl'atlimc} OR {nlaka'pamux} OR okanagan OR {Sec wepmc} OR tlingit OR anishinaabe OR blackfoot OR nakoda OR tasttine OR {Tsuu T'ina} OR "Tsuut'ina" OR {Gwich'in} OR ( han AND NOT chinese ) OR tagish OR tutchone OR algonquin OR nipissing OR ojibwa OR potawatomi OR innu OR maliseet OR "Mi'kmaq" OR micmac OR passamaquoddy OR haudenosaunee OR cayuga OR mohawk OR oneida OR onodaga OR seneca OR tuscarora OR wyandot OR aboriginal* OR indigenous* OR metis OR {red road} OR {on reserve} OR {off-reserve} OR {First Nation} OR {First Nations} OR "Original People*" OR indigenous* OR amerindian OR ( urban W/3 ( indian* OR native* OR aboriginal* ) ) OR {country food*} OR {residential school*} OR ( ( {traditional medicine*} OR {traditional heal*} OR ethnomedicine* ) AND NOT ( korea* OR chinese ) ) OR shaman* OR {traditional food*} OR {medicine man} OR {medicine men} OR "medicine woman" OR {medicine women} OR autochtone* OR ( native* W/1 ( man OR men OR women OR woman OR boy* OR girl* OR adolescent* OR youth OR youths OR person* OR adult OR patient OR patients OR elder OR elders OR elderly OR people* OR indian* OR nation OR tribe* OR tribal OR band OR bands ) ) ) AND TITLE-ABS-KEY ( canad* OR "British Columbia" OR {Colombie Britannique} OR alberta OR saskatchewan OR manitoba OR ontario OR quebec OR {nova scotia} OR {New Brunswick} OR newfoundland OR labrador OR {Prince Edward Island} OR "Yukon" OR nwt OR {Northwest Territories} OR nunavut OR nunavik OR nunatsiavut OR nunatukavut ) ) ) ) ) AND ( ( TITLE-ABS-KEY ( ( gynecologic* OR rectal OR rectum OR genital OR prostat* OR uterin* OR uterus OR ovarian OR endometrial ) W/3 ( surgery OR surgical ) ) AND ( "post surg*" OR "postsurgic*" OR "post operative*" OR "postoperative" ) ) OR ( TITLE-ABS-KEY ( ( prolaps* W/3 ( "pelvic organ" OR uterine OR uterus OR bladder OR rectal OR rectum OR urethra* OR apical ) ) ) ) OR ( TITLE-ABS-KEY ( incontinen* OR uterocele OR cystocele OR rectocele OR enterocele OR urethrocele OR sigmoidocele ) )	OR ( TITLE-ABS-KEY ( bowel W/3 ( frequency OR urgency ) ) ) OR ( TITLE-ABS-KEY ( bladder* W/3 ( overactive OR urgency OR urgent OR frequency OR heistency OR retention ) ) ) OR ( TITLE-ABS-KEY ( constipat* OR encopresis OR dyspareunia OR vulvodynia OR vaginismus OR vestibulodynia OR "interstitial cystitis" OR "painful bladder syndrome" OR "proctalgia fugax" OR prostatitis OR endometriosis OR adenomyosis OR "pudendal neuralgia" OR anismus OR "vaginal wind" OR "pelvic girdle pain" OR "neurogenic bladder" ) ) OR ( TITLE-ABS-KEY ( ( dyssynergia W/3 ( bowel* OR bladder* ) ) ) ) OR ( TITLE-ABS-KEY ( "sexual dysfunction" OR "erectile dysfunction" OR "persistent genital arousal" OR impotence ) ) OR ( TITLE-ABS-KEY ( "bed wett*" OR bedwett* OR encopresis OR "hirshprung* disease" ) ) OR ( TITLE-ABS-KEY ( "gender confirmation surg*" OR "sex reassignment surg*" ) ) OR ( TITLE-ABS-KEY ( "lichen sclerosis" ) ) OR ( TITLE-ABS-KEY ( ( ( fistula* OR fistulae ) W/3 ( bladder* OR urinary OR vagina* OR rectovaginal* OR vesicovaginal* ) ) ) ) OR ( TITLE-ABS-KEY ( "levator ani avulsion" OR noctura OR "voiding dysfunction" OR "LUTS" OR "lower urinary tract symptom*" ) ) )	AND ORIG-LOAD-DATE > 20230620 AND NOT INDEX ( medline OR embase )

**Cochrane Library Searched August 18, 2022**

ID	Search	Hits

#1	MeSH descriptor: [Urinary Incontinence] explode all trees	3498

#2	MeSH descriptor: [Urogenital Diseases] explode all trees	108779

#3	MeSH descriptor: [Constipation] explode all trees	2359

#4	MeSH descriptor: [Colorectal Neoplasms] explode all trees	12560

#5	MeSH descriptor: [Gynecologic Surgical Procedures] explode all trees	5969

#6	MeSH descriptor: [Urogenital Neoplasms] explode all trees	22347

#7	(((gyneologic* or rectal or rectum or genital or prostat* or unterine or
uterus or ovarian or endometrial) Near 3 (surgery or surgical))):ti,ab,kw	1668

#8	#4 or #5 or #6 or #7	40481

#9	MeSH descriptor: [Postoperative Complications] explode all trees	54552

#10	postoperative or "post operative" or "post surgical"	162329

#11	#9 or #10	167895

#12	#8 and #11	7853

#13	MeSH descriptor: [Pelvic Organ Prolapse] explode all trees	976

#14	MeSH descriptor: [Female Urogenital Diseases] explode all trees	58988

#15	(uterocele or cystocele or rectocele or enterocele or urethrocele or

sigmoidocele or incontinence or constipation or encopresis or "vaginal wind"

or "pelvic girdle pain"):ti,ab,kw	27280

#16	((bladder* near 3 (overactive or urgency or urgent or frequency or hesitency or retention))):ti,ab,kw	585

#17	((dyspareunia or vulvodynia or vaginismus or vestibulodynia or
 "interstitial cystitis" or "painful bladder syndrome" or "proctalgia fugax"

or prostatitis or endometriosis or adenomyosis or "pudendal neuralgia" or

anismus)):ti,ab,kw	6451

#18	MeSH descriptor: [Sex Reassignment Surgery] 3 tree(s) exploded	7

#19	MeSH descriptor: [Lichen Sclerosus et Atrophicus] explode all trees	54

#20	MeSH descriptor: [Erectile Dysfunction] explode all trees	1920

#21	MeSH descriptor: [Urinary Fistula] explode all trees	42

#22	#1 or #2 or #3 or #12 or #13 or #14 or #15 or #16 or #17 or #18
or #19 or #20 or #21	138903

#23	MeSH descriptor: [Indigenous Peoples] explode all trees	779

#24	(Athapaskan or Saulteaux or Wakashan or Cree or Dene or Inuit or Inuk
or Inuvialuit* or Haida or Ktunaxa or Tsimshian or Gitxsan or Gitksan or "Nisga'a"
 or Haisla or Heiltsuk or Oweenkeno or "Kwakwaka'wakw" or "Nuu chah nulth" or
 "Tsilhqot'in" or Dakelh or "Wet'suwet'en" or Sekani or Dunne-za or Dene or Tahltan
or Kaska or Tagish or Tutchone or Nuxalk or Salish or St'at'imc):ti	47

#25	(Stl'atl'imx or Stl'atl'imc or Nlaka'pamux or Okanagan or "Sec wepmc"
 or Secwepemc or Tlingit or Anishinaabe or Blackfoot or Nakoda or Tasttine or
 "Tsuu T'ina" or "Tsuut'ina" or "Gwich'in"):ti	1

#26	(Algonquin or Nipissing or Ojibwa or Potawatomi or Innu or Maliseet or

"Mi'kmaq" or Micmac or Passamaquoddy or Haudenosaunee or Cayuga or Mohawk

or Oneida or Onondaga or Seneca or Tuscarora or Wyandot or Aboriginal* or

Indigenous* or Metis):ti	665

#27	("First Nation" or "First Nations" or Amerindian or "torres strait islander"
 or aborigine* or "indigenous people" or maori or Saami or Sami or sampi or
 Southernsami* or Umesami* or Pitesami* or Lulesami* or Northernsami* or
Enaresami* or Kolasami* or Lapp or Lapps or Lappish or Lappland or
 (Lapland* not longspur) or Lappalainen* or Saamelainen* or reindeer herd*
or reindeer culture* or reindeer pastoral* or Lappbys or Samebys or
reinbeitesdistrikt or paliskunta or siida):ti	190

#28	(Abakuria or Abaluhya or Abagusii or Abakuria or Aembu or Agikuyu

or Akamba or Anuak or Anywaa or Amazigh or Ambala or Ambeere or Ambundu

or Ambuun or Amharan or Angba or Baaka or Baamba or Babindi or Babini or

Baboma or Bachokwe or Bacwa or Bafumbira or Baganda or Bagyele or Bagwere

or Bagyeli or Bakiga or Bakola or Baholo or Bakalanga or Bakiga or Bakolo or

Bakongo or Bakonjo or Baluba or Balunda or Balovale or Bamasaba or Bambuti

or Bangala or Bangoli or Bangungu or Bantu or Banyankole or Banyarwanda or

Banyole or Banyoro or Bapende or Bapedi or Barabaig or Barombi or Barundi or

Baruuli or Basamia or Basoga or Batswana or Batooro or Batsamba or Batswana or

Batwa or Bayaka or Bedzan or Bazombe or Bebayaka or Bedzan or Bhaca or Biaka or

Borana or Chewa or Copts or Cormorian or Cushitic or Dahalo or Datooga or Dikidiki

or Dogon or Ewondo or Fulani or Fuliru or Ganguela or Gciriku or Gyele or Hadza

or Hadzabe or Haillom or Haratin or Herero or Himba or Hlubi or Iriryen or

Iqvayliyen):ti	26

#29	(Kabyle* or Kalenjin or Kanioka or Kanioka or Kaonde or Karamojong or

Kavango or Kereuyu or Khoikhoi or KhoiSan or Kikuyu Kwangali or Lokele or

Lowme or Lotuko or Lwalwa or Maasai or Makonde or Makua or Mande or Masalit

or Matumbi or Mayeuyi or Mayeyi or Mbenga or Mbukushu or Mbochi or Mboro

or Mbuti or Medzan or Mijikenda or Mozabite* or Nafusa or Ndebele or Ngombe or

Namaqua or Nyanga or Nyamwezi or Ogiek or Ovambo or Ovimbundu or Phuthi or

Pokomo or Rendille or Riffian or Riffians or Sakuma or Samburu or Sandawe or

Sangha or Sango or Sengwer or Serer or Sesotho or Shangaan or Shawiya or Shenwa

or Shi or Shilluk or Sukua or Sukus or Swahili or Tabwa or Tambuka or Taveta or

Thembu or Tigrayan or Topoke or Tsonga or Toubou or Tuareg or Tumbuka or Ugana

or Wochua or Xhosa or Xindonga or Yoruba or Zenati or Zuwara):ti	199

#30	((Indigenous or Afar or Afars or "Aka People" or Akie or Ameru or Asua
or Ateker or Atwot or Awjila or Bafia or Baka or Bakongo or Bakwe or Balunda or
Balovale or Bango or Bassa or Beja or Bekpak or Bemba or Bembe or Benet or
Berber or Berbers or Bira or Bowe or Bubi or Budja or Bulu or Bunrun or Chaga or
 Chopi or Damara or Dinka or Djerba or Duala or Dzing or Efe or Elmolo or Fang or
 Foora or Fula or Fur or Ghomara or Ghadames or Gllana or Glu or Gogo or Gongo or
Haya or Havu or Hema or Hima or Hunde or Hutu or Huva or Iboko or Igbo or Ijo or
Jieng or Kadu or Kande or Kango or Katla or Kgaga or Khoe or Kola or Komo or Kota
 or Kua or Kuba or Kwango or Kx'z or Kxoe or Lala or Lozi or Luo or Luba or Lupu or Masmuda or Matmata or Mbala or Mbam or "Mbo People" or Mbolo or Mbuza or
Mongo or Mpondo or Myene or Naadh or Nama or Nande or Naro or Ngoni or Ndau
or Ndebele or Ngoli or Ngondi or Ngoni or Nguni or Nkoya or Nkumu or Nuba or
Nubian or Nuer or Nzebi or Ogoni or Omoro or Oroko or Pygmy or Popoi or Poto or
Puru or Rashad or Sango or Sanhaja or Sena or Shilha or Shira or Shona or Shua or
Sokna or Somali* or Sotho or Sua or Subu or Swazi or Taitaa or Tchokwe or Teke or
Tembo or Tetela or (Tonga and Africa*) or Tshwa or Tsoa or Twa or Turkana or Tuu
or Venda or Vira or Watta or Wakuti or Yaaku or Yaka or Yakoma or Yanzi or Yao or
Yeke or Yela or Yeyi or Zulu) Near 3 (population* or people* or person* or elder*
 or man or men or woman or women or child* or youth* or clan or clans or tribe or
 tribes or tribal or family or families or parent* or grandparent* or elder or elders or grandmother* or grandfather* or baby or babies or infant or infants or patient or
 patients or speakers or speaking or village* or communit*)):ti,ab,kw	839

#31	(Acatec or Aguacateco or Amuzgo or Bokota or Boruca or Bribri or "Bri Bri"
 or Buglere or Cabecar or Cakchiquel or Changuena or Chatino or Chiapanec or
Chicomuceltec or Chinantee or Chocho or Cholti or "Ch'olti'" or "Ch'olti'anor
Chontal" or Chorotega or Chorti or Chuj or Chumbia or Corobici or
(Cueva not Spain) or Cuicatec or Cuitlatee or Cuytec or Dorasque or Embera or
Garifuna or Guatuso or Guaymi or Guaymis or Guetar or Huastec or Huave or
Huetar or Itzaj or Ixil or Jacalteco or Jonaz or Kanjobal or Kekchi or Kuna or Maleku
 or Mangue or Matambu or Matlatzinca or Mazahua or Motozintlec or Mayan or
Mayangna or Miskito or Mixtec or Mopan or Nahua or Nahuatl or Ngabe or Otomi
or Pantec or Paya or Popoloca or Popoloc or Poqomam or Poqomchi or "Q'eqchi'" or
 Quiche or Quitirrisi or Sacapulteco or Sipacapense or Subtiaba or Tacaneco or
Tarasco or Tamaulipec or Tamazultec or Tecoxquin or Tectiteco or Tecual or
Tecuexe or Tepehura or Tepuztecor or Teribe or Terraba or Totonac or Trique
or Tzeltal or Tzotzil or Tzutujil or Ulwa or Uspantec* or Uspanteko or Voto or
Xinca or Waunana or Wounaan or Yucatec or Zapotec or Zoque):ti	13

#32	("A' ani" or Absaroka or Haaninin or Atsina or "Gros Ventre" or Acopsel or
 Tlacopsel or Lacopsel or Ahtna or Ahtena or Akenitsi or Occaneechi or Akokisa
or Horcoquisa or Orcoquizas or Aleut or Unangax or Unangan or Alibamu or
"Alabama Alsea" or Alutiiq or Sugpiag or Amahami or Awaxawi or Androscoggin or Arosaguntacook or Ameriscoggin or Anishinaabeg or Chippewa or Anihsinape or
Saulteaux or Apalachee or Aranama or "Texan Coahuilteca" or Tamique or Arikara
 or Sahnish or Arickaree or Adakadaho or Assiniboine or Hohe or Nakota or Nakoda
 or Nakona or "Atsa' Kudok-wa" or Awatixa or Bannock or "Snake Indian" or Bidai or Quasmigdo or Biloxi or Blackfoot or Niitsitapi or Sikasikaitsitapi or Cahto or
 Kaipomo or Cahuilla or Ivilyuqaletem or Ivilyuat or Catawba or Inna or Iswa or
Chemehuevi or Chickasaw or "Chilula Chimakum" or Aqokulo or Chimariko or
Chiricahua or Tsokanende or Chitimacha or Chetimachan or Sitimacha or
 Chowanoke or Roanoke or Chumash or Ciboney or "Taino Ciwat" or Clatsop or
Coos or Coosa or Uchis or Chiaha or Coste or Talisi or Coquille or Kokwell or Coso
 or Cowlitz or Taitnapam or "Crow Nation" or "Cui Ui Ticutta" or Cupeno or
 Kuupangaxwichem or Cupa or "Cup' ig" or Nunivak or "Dakota Oyate" or Lakota
 or Nakota or Santee or Teton or Sioux or Deadose or "Deg Xina" or "Deg Xit' an" or Kaiyuhkhotana or "Deg Hit' an" or "Dena' ina" or Tanaina or "Dichinanek' Hwt' ana"
 or "Upper Kuskokwim Athabascan" or Kolchan or Goltsan or "Tundra Kolosh"
or "Do lkabya" or Duwamish or Esselen or Eyak or "Gidi' tikadi" or Guwevkabaya
or "Gwich' in" or Kutchin or Haida or Xaadas or Xaat or Halchidhoma or Havasupai
 or "Green Water People" or Hiratsa or Hiraaca or "Ho-chaaqa" or Winnebago or
Holikachuk or Innoko or "Tlegon-khotana" or Hopi or "Houma-Louisiana" or Huaco
 or Waco or Hualapai or Hupa or Natinixwe or "Natinook-wa" or "Hwech' in" or
Hankutchin or "Iroquois Confederacy" or "Hodinoso ni" or "Illinois Confederacy"
or "Illinois Confederation" or Ilinoweg or Illini or Inupiat or Inuit or Ioway or Baxoje
or Jicarilla or Juaneno or Acjachemen or Jumano or Kalapuya or Clackama or
 Kalispel or "Pend d' Oreilles" or Qlispe or Karuk or Karok or "Chum-ne" or Katkoc
or Kansa or Kanza or Kawaiisu or Nuwa or Kennebec or "Kinipekw Kittitas" or
 Klickitat or "Qwu' lh-hwai-pum" or "Awi-adshi" or Mahane or Wahnookt or
"Koa' aga' itoka" or Keresan or Kichai or Kitsai or Keechi or "K' itaish" or Kiowa
 or Gaigwu or Cauigu or Kutjau or "Kwu-da" or "Tep-da" or Kitanemuk or Kittitas
 or Klickitat or "Qwu' lh-hwai-pum" or "Awi-adshi" or Mahane or Wahnookt or
 "Koa' aga' itoka" or Konkow or "Koop Ticutta" or Koyukon or Ktunaxa or Kootenai
or Flathead or Kucadikadi or "Kotsa' va" or Kumeyaay or "Tipai-Ipai" or Kamia or
Diegueno or Kwapa or Cocopah or Cucapa or "Xawitt kwnchawaay" or Lassik or
Lenape or "Leni-Lenape" or Lipan or Luiseno or Payomkawichum or Madqwadabaya
 or "Desert Yavapai" or Mahican or Mohicans or Makah or Makuhadokado or
Maliseet or Wolistoqiag or Manahoac or Mahock or Meipontsky or Mandan or
Mattole or "Bear River" or "Tul' bush" or "Ni' ekeni" or Meherrin or Menominee
 or Mackinac or Mescalero or Myaamiaki or Kickapoo or Twigtwee or Missouria
or Miwok or Miwuk or Moadokado or Modoc or Mohave or "Aha Makhav" or
Mohawk or "Kaneng' hega" or Molala or Molale or Molele or Nyyhmy or Moosonee
 or "Moose Cree" or Monsonis or Multnomah or Chinook or Nabedache or
 Nabaydacu or Wawadishe or Nabiltse or Dakubetede or "Nacho Nyak Dun" or
Tutchone or Nacono or "Na' isha" or Nanticoke or Navajo or Ndee or Nial or
Niimiipu or "Nez Perce" or Watapala or Watapahlute or Nisenan or Nisqually or
 Nomlaki or Noamlakee or "Central Wintun" or Nongatl or Nottoway or
Cheroenhaka or "Northern Cheyenne" or Ohlone or Costanoan or Omaha or
 "O' odham" or Pima or Papago or Osage or Otoe or Otse or "Ozav Dika" or
Palus or Passamaquoddy or Pestomuhkati or Patiri or Petaros or Pastia or Patwin
 or "Southern Wintun" or Panis or Skidi or Pedee or Penobscot or "Petun Piipaash" or "Kokmalik' op" or Piscatawa or Doeg or Conoy or "Pit River" or Pomo or Kashaya or
 Ponca or Ponka or Pottawatomi or Bodewadmik or Powhatan or Puyallup or
Spuyalepabs or Quapaw or Ugahxpa or Quechan or Yuma or Kwtsaan or Quileute
 or Salinan or Saponi or Monacan or Sapon or "Eastern Blackfoot" or Christanna
or Sawawatodo or Serrano or Taaqtam or "Maarenga' yam" or Yuhaviatam or Shasta
or Chasta or Sasti or Shoshone or Siletz or Sinkine or Sinkyone or "Siuslaw Umpqua" or Skitswish or "Schitsu' umash" or Snohomish or Snuqualmi or Sokoki or Missiquoi or Stillaguamish or Stoluckwamish or Suquamish or Sutaio or Swinomish or Skagit or
Syilx or Okanagan or Sotaae or "Taga Ticutta" or Takelma or Dagelma or Taltushtuntede or Galice or "Tanan Gwich' in" or Taos or Taovaya or Tataviam or Alliklik or Tawakoni or Tahuacano or Tenino or Thawikila or Hathawekela or "Fort Ancient" or Tigua or
Tillamook or Nehalem or Timbisha or Panamint or Timpanogos or Tlingit or
 "Toi Ticutta" or Tolowa or "Talawa Dini' " or Tongva or Gabrieleno or Fernandeno
 or Tobikhar or Tonkawa or Ticanwatic or Tsikip or Appalousa or Opelousa or
Tsitsistas or Tubatulabal or Tukabatchee or Tuscarora or Tomahittan or Kuskarawock
 or Tutelo or Tutero or Totteroy or Tutera or Yusan or Tututni or Umatilla or Umpqua or Waccamaw or Waxmaw or Wadatika or "Harney Valley Paiute" or Wailiki or
Waluulapam or "Walla Walla" or Walpapi or Huipui or Wampanoag or Massasoit
or Wanapum or Wappo or Washoe or Wichita or Willapa or Kwalhioqua or "Wi pukba"
 or "Verde Valley Yavapai" or Wintu or "Northern Wintun" or Wiyot or "Wee' at" or
Weyet or Yakama or "Yamosopo Tuviwarai" or Yaqui or Yoeme or Yatasi or Yattasih
or "Yavbe" or "Yavapai" or "Ysleta del Sur" or Yojuane or Yokuts or Mariposa or
Yuki or Yupighyt or "Yup'ik" or Yupik or Yurok or "Olekwo'l" or Zuni):ti	303

#33	("Adivasis" or "Adnamanese" or "Andaman" or "Baluch" or "Baluchis" or
 "Bodo" or "Boro" or "Boros" or "Bote" or "Brahuis" or "Chakmas" or "Chepang" or "Chhantyal" or "Damai" or "Dewan" or "Ghale" or "Gurkha" or "Gurung" or "Hayu" or "Hyolmo" or "Jarawa" or "Jirel" or "Jumma" or "Kalash" or "Khas" or "Kirati" or
"Koinch" or "Kulung" or "Kusunda" or "Limbu" or "Lohorung" or "Magar" or
 "Makrani" or "Mangar" or "Marma" or "Miji" or "Mongar" or "Mro" or "Naga" or
"Nepami" or "Newar" or "Nicobar" or "Onge" or "Rang" or "Raute" or "Sajolang"
 or "Santhal" or "Sentinelese" or "Sindhis" or "Sulemani" or "Sunuwar" or "Tamang" or "Thakali" or "Thangmi" or "Tharu" or "Tripura" or "Tumbahangphe" or
 "Wanniyala-Aetto" or "Yakkha" or "Yolmopa"):ti	90

#34	(Aguacatec or Akwa'ala or Abxubal or Ayuukja'ay or "Batzil k'op" or Binizaa or "Chichimeca Jonaz" or Chinantec or Chocho or "Ch ol" or Chontal or "Chuj" or
 Cochimi or Comcaac or Hamasipini or Harijio or "Ha shuta enima" or "Hach t'an"
or Huastecor Hnahnu or Hnatho or Ixcatec or Ixil or Jacaltec or K'akchikel or
K'anjobal or Kanjobal or Kaqchikel or Kechi or K'iche or Kikapooa or Kikapu or
Kiliwa or "Ko'lew" or "K'op o winik atel" or Kumiai or Lacandon or Laymon or
Makurawe or Maya or Maya'wiinik or Mazahua or Mazatec or Me'phaa or
Mexicanero or Mexikatlajtolli or Mixe or Mixtec or Motocintleco or mti'pa or
 Nahuas or Ocuiltec or Otomi or (Oaxaca not Oaxaca-Blinder) or "Pame" or
Papago or Tlahuica or Paipai or "Pima Bajo" or Purepecha or P'urhepecha or
Qatok or (Quiche not Guatemala) or Q'iche or Raramuri or "Runixa ngiigua"
or ("Seri" not "Seri 82") or "Slijuala sihanuk" or Tacuate or Tarahumara or Teenek
 or Tepehua or Ti'pai or Tlapanec or "Tohono O'odham" or Totonac or Tachiwin or
 "Tsa jujmi" or Tzotzil or "Tu'un savi" or Tzeltal or "Uza" or Winik or Xigue or
Yucatec or Zapotec):ti	25

#35	(Abipon or Achuar or Achuagua or Akawaio or Amarizana or Andoque or
Akawaio or Akuriyo or Anauya or Araona or Arawak or Ayamn or Aguaruna or
Amahuaca or Amarakaeri or Andoa or Arabela or Arawak or Arhuaco or Ashaninca or Asheninca or Atsahuaca or Aymara or Ayoreo or Bakairi or "Baniva" or Barasana or
aniwa or "Baure" or Bororo or Cabiyari or Cacataibo or Caquinte or Cacua or
Cahuarano or "Caiua" or "Camara Indians" or Camaracoto or Camsa or Canamari
or Candoshi or Canela or Canichana or Capanahua or Carapana or Cariay or "Carib"
or Carijona or Carutana or Cashibo or Cashinahua or Cawishana or Cavinena or
Caxuiana or Cayuvava or Chontaquiro or Cocama or "Cubeo" or Curipaco or Chacobo or Chaima or (Chana not striatus) or Chapacura or Charrua or Chimila or Chitonahua or
Chorote or Chipaya or Chiquitano or Chulupi or Carare or Coconuco or Cofan or
 Coreguaje or Coyaima or Chamacoco or Chamicuro or Chayahuita or Cocama or
"Culina" or Culino or Cubeo or Cuiba or "Cuiva" or Cumanagoto or Curripaco or
"Deni" or Desano or Embera or Guarani or Guajajara or "Guana" or Guanano or
Guarayo or Guarayu or Guahibo or Guajiro or Guambiano or Guanano or Guayabero or Guarequena or Guinao or "Guana" or Gayon or Guahibo or Hixkaryana or
Huachipairi or Huambisa or Huarayo or Lauanaua or Ikpeng or Ingariko or Irantxe
or Itonama or Inapari or Iquito or Isconahua or Jumana or Japreria or Jirajara or
 Juruti or Jaqaru or Jebero or Kadiweu or Kaingang or Kamayura or Karaja or
Karipuna or "Kariri" or Katukina or Kaxarari or Kayabi or Kayapo or
"Kuikuro alapalo" or Kulina or "Kaiwa" or Kallawaya or "Kogui" or "Kuna"
or Kaweskar or "Lule" or Macuna or Maipure or Mapuche or Mataco or Mocovi
 or Machinere or Machinerev or Machiguenga or Macushi or Macuna or "Madi"
 or Malayo or Mamainde or Manao or Mandauaca or Mandawaka or Mapidian or
 Mapuche or Mapidian or Maquiritare or Maquiritari or Maragua or Marawan or
Mariate or Marubo or Mastanahua or Mataco or Matipuhy or "Matis" or "Matses"
 or Mawakua or Mawakwa or Maxakali or Mehinaku or Miranha or Moronawa or
Munduruku or Movima or Muellama or Muinane or Mapoyo or "Mashco Piro" or
Muniche or Nambikwara or Nocaman or Nuquini or Nomatsiguenga or Nanti or
Ocaina or Omagua or Orejon or "Opon" or Pacahuara or "Paez" or Paicone or Palicur
 or Panare or "Pano" or "Paresi" or Paumari or "Pemon" or Pilaga or Puelche or
 Pauna or Pauserna or Piapoco or Piraha or Piratapuyo or Pisabo or Piaroa or "Pijao"
 or Piratapuyo or Paraujano or Pemon or Pemono or Piapoco or Puinave or Patamona
 or Poyanawa or Puinave or Puquina or Quechua or Quichua or Retuara or Resigaro
 or Reyesano or Sabanes or "Saliba" or Saluma or Sarave or Secoya or Selknam or
 "Sensi" or Shaninawa or Shapra or Sharanahua or Shebayo or Shiwiar or Shikiana
 or Sikiana or Siriono or Sinsiga or "Siona" or Suruwaha or Tacano or Tamanaco
or Tiahuanaco or Tariano or Tehuelche or Tariano or Tatuyo or "Tembe" or "Terena"
 or Telembi or Ticuna or Ticuna or Tiriyo or Tiwanaku or Tiwanaku or "Torom" or
 "Totoro" or Tsimane or Tuberao or "Tucano" or Tunebo or Tuxinawa or Tuyuca or
Uainuma or Urarina or Vilela or Waimaha or Waiampi or Waiwai or Wapishana or
 Waraiku or Warekena or Waura or Wayampi or Wayana or Wirina or Waimaha or
Waunana or "Wiwa" or "Warao" or Wayuu or Witoto or Xavante or Xipaya or
Xiriana or Xokleng or Yabaana or Yaminawa or Yaminahua or Yaruma or Yawalapiti
 or Yuracare or Yabarana or Yavitero or "Yine" or Yamana or Yaghan or Yucuna or
 Yurumangui or Yukpa or Yanesha or Yoranahua or Yagua or Yaminahua or Zaparo
or Zamuco or "Trio Indians" or "More Indians" or "Bare Indians"):ti	24

#36	((("Inga" or "Maca" or "Leco" or "Mojo" or "Uro" or "Maco" or Lengua or
 "Toba" or "Zoe" or "Ona" or "Catio" or "Passe" or "Bari" or "Awa" or "Bora" or
 "Bara" or "Remo" or "Pano" or "Sape") NEAR 3 (Indians or Indian or Indigenous or Amerindian* or Aborigin* or people or peoples or elder or elders or grandmother* or grandfather* or parent* or women or men or woman or man or child* or youth or
youths or baby or babies or tribe or tribes or tribal or shaman* or native or patient
 or patients))):ti	2

#37	(Ainus or Ainu or Aleuts or Alyutors or Chukchis or Chuvans or Dolgans
 or Enets or Entsy or Yupik or Yup'ik or Yuit or Yupigyt or Chaplino or Naukan
or Itelmens or Kamchadals or Kereks or "Komi" or Koryaks or Nenets or Nentsy
 or Nganasans or Tavgi or Sami or Veps or Yukaghirs or Chulyms or Evenks or
Tungus or Evens or "Kets" or Khantys or Mansi or Vguls or Selkups or Teleuts
or Nanais or Nanaitsy or Negidal or Nivikh or Oroch or orok or Taz or udege or
 ulch or Kumadins or Chelkans or Shorians or Soyots or Telengits or Tofalars or
Tugalars or "Tufans Todzhins" or Laks or Tabasarans or Turuls or Aguls or Tsakhurs
 or Kumyks or Nogais or "Andis" or Akhvakh or Archins or Bagvalals or Bezhta or
 Botlikhs or Chamalals or Godoberi or Hinukh or Hunzibs or Khwarshi or Karata or
 Tindis or Tsez or Abazin or Besermyan or Izhorians or Karelians or Nagaybaks or
Setos or Shapsugs or Quratay):ti	45

#38	(Greenlander or "Kalaallit Nunaat" or Nuuk or Sisimiut or Ilulissat or
 "Qaqortoq Aasiaat" or Maniitsoq or Tasiilaq or Uummannaq or Narsaq or Paamiut
 or Nanortalik or Upernavik or Qasigiannguit):ti,ab,kw	2

#39	("Abui" or "Aeta" or "Alfur" or "Ati People" or "Bahau" or "Bali Aga" or
"Bali Mula" or "Baliaga" or "Balinese" or "Basap" or "Bataq" or "Batek" or "Bateq"
or "Batin" or "Blaan" or "Brao" or "Bru" or "Bru-Van Kieu" or "Bugkalot" or
 "Bumiputera" or "Bunong" or "Chams" or "Champa" or "Cheq Wong" or "Dasun" or
"Dayak" or "Deli Malay" or "Filipina" or "Filiopino" or "Gaddang" or "Ibaloi" or
 "Iban" or "Idan" or "Igorot" or "Jahai" or "Jehai" or "Jakun" or "Javanese Kshatriya"
 or "Kangeanese" or " Kagayanen" or "Kanayatn" or "Katu" or "Kayan" or "Kavet"
or "Kenyah" or "Khmer Krom" or "Kimaragang" or "Klemantan" or "Kreung" or
 "Kuijau" or "Kuy" or "Kwijau" or "Lampung" or "Lanoh" or "Lawangan" or
 "Luangan" or "Lumad" or "Dayak" or "Loloan Malay" or "Mah Meri" or
"Malay Singaporean" or "Malbog" or "Maluku" or "Mangka'ak" or "Mangyan" or
"Maniq" or "Maragang" or "Ma'anyan" or "Minokok" or "Moluccas" or Montagnard
 or "Murut" or "Naga" or "Negrito" or "New Guinea" or "Ngaju" or "Orang Hulu" or
 "Orang Kanaq" or "Orang Kuala" or "Orang Seletar" or "Orang Ulu" or "Ot Danum" or "Palawano" or "Palembang" or " Panay-Bukidnon" or "Pan-ayanon" or "Papua" or
 "Phnong" or "Punong" or "Pear People" or "Por People" or "Punan" or "Rumanau"
or "Sasak" or "Semai" or "Semang" or "Semaq Beri" or "Semelai" or "Senoi" or
 "Simeulue" or "Suludnon" or "Sunda Island" or "Tabanwa" or "Takua" or "Tambanuo"
or "Tombonuo" or "Taron" or "Temiar" or "Temoq" or "Temuan" or "Tidung" or "Trone"
 or "Taaw't Bato"):ti	185

#40	("pacific islander" or Banabans or "Native Hawaiian" or Guamanian or
Chamarro* or Chuukese or I-Kiribati or Kosraean* or Maohi or Marshallese or
 Moriori or Mortlockese or Mwokilese or Namonuito or Nauruans or Ngatikese or
Niuean or Paafang or Paluans or Pingelapese or Pohnpeian* or Pollapese or Puluwat
 or Rapanui or Rotuman or Satwalese or Samoan*or Sonsorolese or Tahitian* or
Tobian or Tongan or "Torres Strait Islander" or Tuvaluan or Ulithian or Woleian or
"Yapese or Polynesian* or Micronesian* or Melanesian*):ti	118

#41	("Bedouin" or "Jahalin" or "al-Kaabneh" or "al-Azazmeh" or "al-Ramadin" or "al-Rshaida"):ti	2

#42	#23 or #24 or #25 or #26 or #27 or #28 or #29 or #30 or #31 or #32 or #33
or #34 or #35 or #36 or #37 or #38 or #39 or #40 or #41	3079

#43	#22 and #42	271

Limit 2023-07-01 to 2024-02-14 (Trials only)	8

**PROSPERO Searched August 24, 2022**

Search updated June 21, 2022 retrieving 2 additional results

LineSearch forHits

#1 MeSH DESCRIPTOR Urinary Incontinence EXPLODE ALL TREES	205

#2 MeSH DESCRIPTOR Fecal Incontinence EXPLODE ALL TREES	45

#3 pelvic organ prolapse	268

#4 MeSH DESCRIPTOR Pelvic Organ Prolapse EXPLODE ALL TREES	56

#5 MeSH DESCRIPTOR Cystocele EXPLODE ALL TREES	0

#6 MeSH DESCRIPTOR Rectal Prolapse EXPLODE ALL TREES	9

#7 MeSH DESCRIPTOR Uterine Prolapse EXPLODE ALL TREES	18

#8 MeSH DESCRIPTOR Visceral Prolapse EXPLODE ALL TREES	0

#9 MeSH DESCRIPTOR Urinary Bladder, Overactive EXPLODE ALL TREES	50

#10 MeSH DESCRIPTOR Urinary Retention EXPLODE ALL TREES	25

#11 MeSH DESCRIPTOR Constipation EXPLODE ALL TREES	87

#12 MeSH DESCRIPTOR Encopresis EXPLODE ALL TREES	0

#13 MeSH DESCRIPTOR Pelvic Floor Disorders EXPLODE ALL TREES	25

#14 MeSH DESCRIPTOR Pelvic Girdle Pain EXPLODE ALL TREES	7

#15 MeSH DESCRIPTOR Dyspareunia EXPLODE ALL TREES	10

#16 MeSH DESCRIPTOR Vulvodynia EXPLODE ALL TREES	10

#17 MeSH DESCRIPTOR Vaginismus EXPLODE ALL TREES	3

#18 MeSH DESCRIPTOR Adenomyosis EXPLODE ALL TREES	26

#19 MeSH DESCRIPTOR Pudendal Neuralgia EXPLODE ALL TREES	1

#20 MeSH DESCRIPTOR Gynecologic Surgical Procedures EXPLODE ALL
TREES	236

#21 MeSH DESCRIPTOR Colectomy EXPLODE ALL TREES	47

#22 MeSH DESCRIPTOR Colorectal Neoplasms EXPLODE ALL TREES	711

#23 MeSH DESCRIPTOR Endometrial Neoplasms EXPLODE ALL TREES	79

#24 MeSH DESCRIPTOR Uterine Neoplasms EXPLODE ALL TREES	342

#25 MeSH DESCRIPTOR Urogenital Neoplasms EXPLODE ALL TREES	1350

#26 MeSH DESCRIPTOR Genital Neoplasms, Female EXPLODE ALL TREES	539

#27 MeSH DESCRIPTOR Genital Neoplasms, Male EXPLODE ALL TREES	455

#28 MeSH DESCRIPTOR Ovarian Neoplasms EXPLODE ALL TREES	163

#29 MeSH DESCRIPTOR Prostatectomy EXPLODE ALL TREES	116

#30 MeSH DESCRIPTOR Prostatic Neoplasms EXPLODE ALL TREES	415

#31 ((gynecologic* or rectal or rectum or genital or prostat* or unterine or
uterus or ovarian or endometrial) and (surgery or surgical))	2932

#32 #21 OR #22 OR #23 OR #24 OR #25 OR #26 OR #27 OR #28 OR #29
 OR #30 OR #31	4446

#33 MeSH DESCRIPTOR Postoperative Complications EXPLODE ALL TREES	1338

#34 postoperative or "post operative" or "post surg*"	12434

#35 #33 OR #34	12846

#36 #32 AND #35	1112

#37 neurogenic urinary bladder	4

#38 MeSH DESCRIPTOR Urinary Bladder, Neurogenic EXPLODE ALL TREES	17

#39 MeSH DESCRIPTOR Sexual Dysfunction, Physiological EXPLODE ALL TREES	164

#40 MeSH DESCRIPTOR Erectile Dysfunction EXPLODE ALL TREES	73

#41 MeSH DESCRIPTOR Impotence, Vasculogenic EXPLODE ALL TREES	2

#42 MeSH DESCRIPTOR Nocturnal Enuresis EXPLODE ALL TREES	12

#43 MeSH DESCRIPTOR Hirschsprung Disease EXPLODE ALL TREES	9

#44 MeSH DESCRIPTOR Sex Reassignment Surgery EXPLODE ALL TREES	5

#45 MeSH DESCRIPTOR Lichen Sclerosus et Atrophicus EXPLODE ALL TREES	7

#46 MeSH DESCRIPTOR Urinary Fistula EXPLODE ALL TREES	10

#47 MeSH DESCRIPTOR Rectovaginal Fistula EXPLODE ALL TREES	2

#48 MeSH DESCRIPTOR Rectovaginal Fistula EXPLODE ALL TREES	2

#49 MeSH DESCRIPTOR Pelvic Floor EXPLODE ALL TREES	128

#50 #1 OR #2 OR #3 OR #4 OR #5 OR #6 OR #7 OR #8 OR #9 OR #10 OR
#11 OR #12 OR #13 OR #14 OR #15 OR #16 OR #17 OR #18 OR #19 OR
 #20 OR #36 OR #37 OR #38 OR #39 OR #40 OR #41 OR #42 OR #43 OR
 #44 OR #45 OR #46 OR #47 OR #48 OR #49	2158

#51 (prolaps* and ("pelvic organ" or uterine or uterus or bladder or rectal or
rectum or urethra* or apical))	369

#52 incontinence	1283

#53 (uterocele or cystocele or rectocele or enterocele or urethrocele or sigmoidocele)	38

#54 (bladder* and (overactive or urgency or urgent or frequency or heistency or
retention))	627

#55 (bowel and (frequency or urgency))	659

#56 constipation	946

#57 encopresis	17

#58 (dyssynergia and (bowel* or bladder*))	14

#59 "vaginal wind"	1

#60 "pelvic girdle pain"	57

#61 (dyspareunia or vulvodynia or vaginismus or vestibulodynia or
"interstitial cystitis" or "painful bladder syndrome" or "proctalgia fugax"
 or prostatitis or endometriosis or adenomyosis or "pudendal neuralgia"
or anismus)	933

#62 ("sexual dysfunction" or "erectile dysfunction" or "persistent genital arousal"
 or impotence)	860

#63 ("bed wett*" or bedwett* or encopresis or "hirshprung* disease")	36

#64 ("gender confirmation surg*" or "sex reassignment surg*")	8

#65 "lichen sclerosis"	5

#66 nocturia	165

#67 ((fistula* or fistulae) and (bladder* or urinary or vagina* or rectovaginal* or vesicovaginal*))	149

#68 "levator ani avulsion"	6

#69 ("Lower urinary tract symptom*" or dysuria or "LUTS")	429

#70 "voiding dysfunction"	54

#71 #70 OR #69 OR #68 OR #67 OR #66 OR #65 OR #64 OR #63 OR #62
 OR #61 OR #60 OR #59 OR #58 OR #57 OR #56 OR #55 OR #54 OR #53
OR #52 OR #51 OR #50	5649

#72MeSH DESCRIPTOR Indigenous Peoples EXPLODE ALL TREES	10

#73 ((Indigen* or aborig* or Amerindian or inuit or metis or sami or maori or
"native american*" or "native hawaiian") or ((native* or tribe or tribal or tribes)
 and (people or peoples or elder or elders or grandmother* or grandfather* or
parent* or women or men or woman or man or child* or youth or youths or
 baby or babies or shaman* or native or patient or patients))):TI,KW	376

#74 #72 OR #73	377

#75 #71 and #74	4

LineSearch forHits

#1	MeSH DESCRIPTOR Urinary Incontinence EXPLODE ALL TREES	205

#2	MeSH DESCRIPTOR Fecal Incontinence EXPLODE ALL TREES	45

#3	pelvic organ prolapse	371

#4	MeSH DESCRIPTOR Pelvic Organ Prolapse EXPLODE ALL TREES	56

#5	MeSH DESCRIPTOR Cystocele EXPLODE ALL TREES	0

#6	MeSH DESCRIPTOR Rectal Prolapse EXPLODE ALL TREES	9

#7	MeSH DESCRIPTOR Uterine Prolapse EXPLODE ALL TREES	18

#8	MeSH DESCRIPTOR Visceral Prolapse EXPLODE ALL TREES	0

#9	MeSH DESCRIPTOR Urinary Bladder, Overactive EXPLODE ALL TREES	50

#10	MeSH DESCRIPTOR Urinary Retention EXPLODE ALL TREES	25

#11	MeSH DESCRIPTOR Constipation EXPLODE ALL TREES	87

#12	MeSH DESCRIPTOR Encopresis EXPLODE ALL TREES	0

#13	MeSH DESCRIPTOR Pelvic Floor Disorders EXPLODE ALL TREES	25

#14	MeSH DESCRIPTOR Pelvic Girdle Pain EXPLODE ALL TREES	7

#15	MeSH DESCRIPTOR Dyspareunia EXPLODE ALL TREES	10

#16	MeSH DESCRIPTOR Vulvodynia EXPLODE ALL TREES	10

#17	MeSH DESCRIPTOR Vaginismus EXPLODE ALL TREES	3

#18	MeSH DESCRIPTOR Adenomyosis EXPLODE ALL TREES	26

#19	MeSH DESCRIPTOR Pudendal Neuralgia EXPLODE ALL TREES	1

#20	MeSH DESCRIPTOR Gynecologic Surgical Procedures EXPLODE ALL TREES0

#21	MeSH DESCRIPTOR Colectomy EXPLODE ALL TREES	47

#22	MeSH DESCRIPTOR Colorectal Neoplasms EXPLODE ALL TREES	712

#23	MeSH DESCRIPTOR Endometrial Neoplasms EXPLODE ALL TREES	79

#24	MeSH DESCRIPTOR Uterine Neoplasms EXPLODE ALL TREES	343

#25	MeSH DESCRIPTOR Uterine Neoplasms EXPLODE ALL TREES	343

#26	MeSH DESCRIPTOR Urogenital Neoplasms EXPLODE ALL TREES	1352

#27	MeSH DESCRIPTOR Genital Neoplasms, Female EXPLODE ALL TREES	540

#28	MeSH DESCRIPTOR Ovarian Neoplasms EXPLODE ALL TREES	163

#29	MeSH DESCRIPTOR Prostatectomy EXPLODE ALL TREES	116

#30	MeSH DESCRIPTOR Prostatic Neoplasms EXPLODE ALL TREES	416

#31	((gynecologic* or rectal or rectum or genital or prostat* or unterine or
uterus or ovarian or endometrial) and (surgery or surgical))	4441

#32	MeSH DESCRIPTOR Postoperative Complications EXPLODE ALL TREES	1338

#33	`#21 OR #22 OR #23 OR #24 OR #25 OR #26 OR #27 OR #28 OR #29 OR
 #30 OR #31	5958

#34	MeSH DESCRIPTOR Postoperative Complications EXPLODE ALL TREES	1338

#35	MeSH DESCRIPTOR Postoperative Complications EXPLODE ALL TREES	1338

#36	postoperative or "post operative" or "post surg*"	18927

#37	postoperative or "post operative" or "post surg*"	18927

#38	#35 OR #36	19339

#39	#33 AND #38	1617

#40	neurogenic urinary bladder	6

#41	MeSH DESCRIPTOR Urinary Bladder, Neurogenic EXPLODE ALL TREES	17

#42	MeSH DESCRIPTOR Sexual Dysfunction, Physiological EXPLODE
ALL TREES	165

#43	MeSH DESCRIPTOR Erectile Dysfunction EXPLODE ALL TREES	74

#44	MeSH DESCRIPTOR Impotence, Vasculogenic EXPLODE ALL TREES	2

#45	MeSH DESCRIPTOR Nocturnal Enuresis EXPLODE ALL TREES	12

#46	MeSH DESCRIPTOR Hirschsprung Disease EXPLODE ALL TREES	9

#47	MeSH DESCRIPTOR Sex Reassignment Surgery EXPLODE ALL TREES	5

#48	MeSH DESCRIPTOR Lichen Sclerosus et Atrophicus EXPLODE ALL TREES	7

#49	MeSH DESCRIPTOR Urinary Fistula EXPLODE ALL TREES	10

#50	MeSH DESCRIPTOR Rectovaginal Fistula EXPLODE ALL TREES	2

#51	MeSH DESCRIPTOR Pelvic Floor EXPLODE ALL TREES	128

#52	#39 OR #40 OR #41 OR #42 OR #43 OR #44 OR #45 OR #46 OR #47
OR #48 OR #49 OR #50 OR #51 OR #1 OR #2 OR #3 OR #4 OR #5 OR #6 OR
 #7 OR #8 OR #9 OR #10 OR #11 OR #12 OR #13 OR #14 OR #15 OR #16 OR
#17 OR #18 OR #19 OR #20	2584

#54	(prolaps* and ("pelvic organ" or uterine or uterus or bladder or rectal
 or rectum or urethra* or apical))	497

#55	incontinence1	782

#56	(uterocele or cystocele or rectocele or enterocele or urethrocele or sigmoidocele)	51

#57	(bladder* and (overactive or urgency or urgent or frequency or heistency
or retention))	833

#58	(bowel and (frequency or urgency))	926

#59	(bowel and (frequency or urgency))	926

#60	constipation	1360

#61	encopresis	18

#62	(dyssynergia and (bowel* or bladder*))	20

#63	"vaginal wind"	1

#64	"pelvic girdle pain"	78

#65	(dyspareunia or vulvodynia or vaginismus or vestibulodynia or
 "interstitial cystitis" or "painful bladder syndrome" or "proctalgia fugax"
or prostatitis or endometriosis or adenomyosis or "pudendal neuralgia" or
anismus)	1408

#66	("sexual dysfunction" or "erectile dysfunction" or "persistent genital arousal" or impotence)	1241

#67	("bed wett*" or bedwett* or encopresis or "hirshprung* disease")	42

#68	("gender confirmation surg*" or "sex reassignment surg*")	11

#69	"lichen sclerosis"	11

#70	nocturia	214

#71	((fistula* or fistulae) and (bladder* or urinary or vagina* or rectovaginal* or vesicovaginal*))	207

#72	"levator ani avulsion"	8

#73	("Lower urinary tract symptom*" or dysuria or "LUTS")	594

#74	"voiding dysfunction"	66

#75	#56 OR #57 OR #58 OR #60 OR #61 OR #62 OR #63 OR #64 OR #65 OR
 #66 OR #67 OR #68 OR #69 OR #70 OR #71 OR #72 OR #73 OR #74	5618

#76	MeSH DESCRIPTOR Indigenous Peoples EXPLODE ALL TREES	10

#77	((Indigen* or aborig* or Amerindian or inuit or metis or sami or maori or "native american*" or "native hawaiian") or ((native* or tribe or tribal or tribes) and (people
or peoples or elder or elders or grandmother* or grandfather* or parent* or women
 or men or woman or man or child* or youth or youths or baby or babies or shaman*
 or native or patient or patients))):TI,KW	493

#78#	76 OR #77	494

#79	#75 OR #52 OR #54 OR #55	7887

#80	#78 AND #79	7
